# Subtyping of common complex diseases and disorders by integrating heterogeneous data. Identifying clusters among women with lower urinary tract symptoms in the LURN study

**DOI:** 10.1101/2021.09.17.21263124

**Authors:** Victor P. Andreev, Margaret E. Helmuth, Gang Liu, Abigail R. Smith, Robert M. Merion, Claire C. Yang, Anne P. Cameron, J. Eric Jelovsek, Cindy L. Amundsen, Brian T. Helfand, Catherine S. Bradley, John O. L. DeLancey, James W. Griffith, Alexander P. Glaser, Brenda W. Gillespie, J. Quentin Clemens, H. Henry Lai, the LURN Study Group

**Affiliations:** Arbor Research Collaborative for Health, Ann Arbor, Michigan, United States of America; Department of Computational Medicine and Bioinformatics, University of Michigan, Ann Arbor, Michigan, United States of America; Department of Urology, University of Washington, Seattle, Washington, United States of America; Department of Urology, University of Michigan, Ann Arbor, Michigan, United States of America; Department of Obstetrics and Gynecology, Duke University, Raleigh, North Carolina, United States of America; Department of Urology, North Shore University, Evanston, Illinois, United States of America; Department of Obstetrics and Gynecology, University of Iowa, Iowa City, Iowa, United States of America; Department of Medical Social Sciences, Northwestern University, Chicago, Illinois, United States of America; Department of Biostatistics, University of Michigan, Ann Arbor, Michigan, United States of America; Department of Surgery, Washington University, St Louis, Missouri, United States of America

## Abstract

We present a novel methodology for subtyping of persons with a common clinical symptom complex by integrating heterogeneous continuous and categorical data. We illustrate it by clustering women with lower urinary tract symptoms (LUTS), who represent a heterogeneous cohort with overlapping symptoms and multifactorial etiology. Identifying subtypes within this group would potentially lead to better diagnosis and treatment decision-making. Data collected in the Symptoms of Lower Urinary Tract Dysfunction Research Network (LURN), a multi-center prospective observational cohort study, included self-reported urinary and non-urinary symptoms, bladder diaries, and physical examination data for 545 women. Heterogeneity in these multidimensional data required thorough and non-trivial preprocessing, including scaling by controls and weighting to mitigate data redundancy, while the various data types (continuous and categorical) required novel methodology using a weighted Tanimoto indices approach. Data domains only available on a subset of the cohort were integrated using a semi-supervised clustering approach. Novel contrast criterion for determination of the optimal number of clusters in consensus clustering was introduced and compared with existing criteria. Distinctiveness of the clusters was confirmed by using multiple criteria for cluster quality, and by testing for significantly different variables in pairwise comparisons of the clusters. Cluster dynamics were explored by analyzing longitudinal data at 3- and 12-month follow-up. Five distinct clusters of women with LUTS were identified using the developed methodology. The clinical relevance of the identified clusters is discussed and compared with the current conventional approaches to the evaluation of LUTS patients. Rationale and thought process are described for selection of procedures for data preprocessing, clustering, and cluster evaluation. Suggestions are provided for minimum reporting requirements in publications utilizing clustering methodology with multiple heterogeneous data domains.

## INTRODUCTION

Complex diseases, such as obesity, diabetes, atherosclerosis, Alzheimer’s disease, major depressive disorder, and cancer result from multiple genetic, epigenetic, and environmental factors, and importantly, interactions between these factors [1-3]. Important differences between patients with these complex diseases and disorders may exist at multiple levels, including: (a) symptoms, subjective experiences, and adaptive behaviors; (b) characteristics of the physical state of the organism, including comorbidities; (c) characteristics of organs and systems; and (d) characteristics at the cellular or molecular level. The extent of these differences suggests that some complex diseases and disorders are better represented by subtypes, each of which can potentially have different etiologies, mechanisms, and outcomes, and require different approaches to treatment. Subtype identification is therefore a potentially important aspect to understanding and treating complex diseases and disorders. Specifically, extracting patient characteristics at each level and grouping patients based on these characteristics allows for comprehensive clinical phenotyping and provides necessary information for discovery and implementation of personalized treatments. Characteristics at each level are represented by variables of different types (continuous, categorical, and binary), scales, and different level of relevance or impact for the disease of interest. This data heterogeneity requires thoughtful preprocessing and novel approaches to data integration.

Lower urinary tract symptoms (LUTS) is a general term representing a heterogeneous group of symptoms with sometimes unclear etiology, high economic and social costs, and significant effects on patients’ quality of life. LUTS can include frequent urination during day and night (nocturia), urinary urgency (a sudden urge to urinate), stress and urgency urinary incontinence (UI), and bladder emptying symptoms such as straining, hesitancy (delay to start to urinate), weak urine stream, and post-void dribbling. None of the symptoms is pathognomonic for a particular diagnosis, and many persons have more than one symptom. The prevalence of LUTS in the United States (US) ranges between 45% and 70% and increases with age [4-5]. Given the aging population in the US, the prevalence of LUTS is expected to increase in the coming years [6]. Medical expenditures for LUTS have been reported to be as high as $65 billion per year [7], yet many therapeutic options for the treatment of LUTS do not provide long-term symptom relief. Many patients experience a *combination* of symptoms, so treatment that focuses on a single symptom may result in suboptimal care. To improve treatment outcomes for patients with LUTS, it is necessary to sort out the heterogeneity of this population, increase the understanding of different subtypes of LUTS and their underlying mechanisms.

One way to better understand a complex disease or disorder is to use an unbiased, data driven unsupervised clustering approach to identify subtypes of individuals with the disease. This method uses data to identify groups, or clusters, such that members of each cluster are as similar as possible to others within their cluster, but as different as possible from those in other clusters [8]. Subtypes identified in this manner are based on similarities and differences within the data, remaining agnostic to clinical definitions or diagnostic categories.

Clustering methodologies have generated important contributions to the analysis of health-related data and are becoming increasingly valuable tools as the field of precision medicine progresses. These methods represent a burgeoning field of research [9-15]. Many unsupervised classification or clustering methods have been developed, from commonly used k-means clustering, hierarchical clustering, and self-organizing maps (SOM) [16-18] to algorithms developed in specific areas for specific applications [19-21]. An important problem in unsupervised clustering is determining the optimal number of clusters. Currently available criteria include Calinski-Harabasz, Davies-Bouldin, Dunn, Gap, Silhouette indices, and others [22-26]; however, the optimal number of clusters may vary by the criterion applied. Therefore, another critical issue is to validate the clustering results, that is, to gain confidence about the clinical significance of the putative clusters, both in terms of cluster numbers and cluster assignments.

Disease subtyping by clustering homogeneous “-omics” data of certain types, mostly gene expression data, has been extensively published [27-30]. More recent studies have subtyped complex diseases by clustering heterogeneous data that included patient health questionnaires and other clinical data. These research studies include subtyping asthma by using questionnaires, physiological tests, and lab tests [31]; subtyping type 2 diabetes by using body mass index (BMI), age at onset of diabetes, homoeostasis model assessment estimates, and insulin resistance [32]; and subtyping sepsis using demographics, vital signs, biomarkers of inflammation, and organ dysfunction or injury [33].

Resampling based consensus clustering initially proposed for clustering gene expression data [34] is gaining popularity and has been used for subtyping complex diseases using heterogeneous data [33, 35]. Multiple random resampling of patients followed by k-means clustering generates probabilities that each pair of patients appear in the same cluster, which can be treated as a pairwise distance between patients and used for determination of cluster membership through hierarchical clustering [34]. This method ensures the clustering results are robust to sampling errors.

Previous studies aiming to identify subtypes of LUTS in an unbiased manner include Epidemiology Urinary Incontinence and Comorbidities (EPIC) and Boston Area Community Health (BACH) projects [36-37], which performed clustering of LUTS patients based on a relatively small number of self-reported symptom data in community-dwelling cohorts. Another study used only BMI and bladder diary variables for clustering community-dwelling women with LUTS [38].

The Symptoms of Lower Urinary Tract Dysfunction Research Network (LURN) Observational Cohort Study is a multi-center study that collected self-reported symptoms, 3-day bladder diaries, physical examination, neuroimaging and sensory testing data, and biological samples in over 1000 care-seeking men and women across six tertiary care centers [39-40]. In our previous study [41], we performed clustering of 545 women from the LURN Observational Cohort Study using baseline self-reported urinary symptom data, captured with the LUTS Tool [42-43] and the American Urological Association Symptom Index (AUA-SI) [44]. Four distinct clusters were identified. Women in cluster F1 (n=138) were continent, but reported post-void dribbling, frequency, and voiding symptoms. Cluster F2 (n=80) reported urgency urinary incontinence, as well as urinary urgency and frequency, and minimal voiding symptoms. Cluster F3 (n=244) included women reporting all types of urinary incontinence, urgency, frequency, and mild voiding symptoms. Women in cluster F4 (n=83) reported all LUTS at uniformly high levels. These subtypes of LUTS were based solely on the above two questionnaires and require further refinement, followed by clinical verification.

The current report describes the methodology and results of refining the female LUTS symptom-based clusters by integrating multiple data domains collected in LURN: demographics, non-urinary symptoms, history, and physical examination data, as well as intake and voiding patterns captured in 3-day bladder diaries. We discuss preprocessing of heterogeneous data and combination of continuous and categorical data using our novel weighted Tanimoto indices approach. We use resampling-based consensus clustering [34] combined with a modified semi-supervised clustering approach [45-46] to make use of data available on only a subset of participants. Then we determine the optimal number of clusters using our novel contrast criterion (CC) developed for consensus clustering, and compare it with other consensus clustering criteria: proportion of ambiguous clustering (PAC) [47], and consensus score (CS) [35], as well as with the established quality of clustering criteria, such as Calinski-Harabasz, Davies-Bouldin, Dunn, and Silhouette [22-26]. We identify distinct clusters of women with LUTS and show superiority of these clusters to our published symptom-based clusters [41], in terms of the percentage of significantly different variables in pairwise comparisons of the clusters and the confidence level in the determined cluster membership. Dynamics of the clusters in 12-month follow-up, as well as clinical relevance of the clusters, are discussed.

In the *Methods* section, we describe the analytical pipeline we developed and used for subtyping women with LUTS. We provide the rationale for our choices of methods for data preprocessing, integration, clustering, and cluster evaluation, as well as review of other available options. We demonstrate that the developed pipeline allowed for identification of distinct and robust refined clusters, with a higher percentage of significantly different variables across the clusters than those previously published, and validate cluster distinctiveness by analyzing biomarker data. Finally, we review the methodological information needed to assess subtyping via clustering and propose a set of reporting requirements that should ideally be included in all clustering reports. We finish by calling for the clustering community effort to develop minimum requirements for clustering publications. We believe this paper would be of interest for clinicians and researchers involved in subtyping of common complex disease and disorders using heterogeneous and multidomain data.

## MATERIALS AND METHODS

### LURN data used for subtyping of LUTS

#### Data on women with LUTS

Data for LUTS subtyping were obtained from the LURN Observational Cohort Study [39,40], which included 545 women seeking care for LUTS at six tertiary care centers. Baseline data collection included demographic information, medical history, physical examination findings, 3-day bladder diaries [48], and self-report questionnaires of urologic and non-urologic symptoms. Urologic symptoms were collected using the LUTS Tool [42-43] and the AUA-SI [44]. The LUTS Tool contains 44 items, including questions on the frequency of occurrence and degree of bother for each urinary symptom. Possible answers to the LUTS Tool questions were ranked from zero to four, zero indicating absence of the symptom, and four indicating the most severe level of the symptom. The AUA-SI has eight items, including a single overall bother question. Responses to the first seven questions of the AUA-SI range from zero to five, zero indicating “none” or “not at all”, and five indicating “almost always”. The final question in the AUA-SI ranges from zero (delighted) to six (terrible). Participants also completed patient-reported outcome (PRO) questionnaires from the Patient-Reported Outcomes Measurement Information System (PROMIS). We used questionnaires related to bowel function (PROMIS gastrointestinal constipation, diarrhea, and bowel incontinence subsets) [49], psychological health (PROMIS Depression and Anxiety Short Forms [50], Perceived Stress Scale [51], PROMIS Sleep Disturbance Short Form [52]), urologic pain (Genitourinary Pain Index [GUPI]) [53], and pelvic floor function (Pelvic Floor Distress Inventory [PFDI]) [54]. Demographics included: age, race, ethnicity, employment status, education level, and marital status. Physical examination data included: weight, waist circumference, BMI, post-void residual volume, pelvic organ prolapse (using the Pelvic Organ Prolapse Quantification [POP-Q] system that measures the location of selected landmarks on the vagina and cervix [55]), and presence of pathology findings at the introitus, urethra, vagina, uterus, or rectum. Medical history data included Functional Comorbidity Index (FCI) [56], additional individual comorbidities, as well as information on history of urinary tract infections (UTI), pregnancy, vaginal deliveries, alcohol, smoking, recreational drug use, and medication use. Bladder diaries included data on timing and volume of each beverage intake and urinary void during a 72-hour period. Completeness and accuracy of bladder diaries collected in LURN are described in [57]. Only 193 women (35%) returned bladder diaries deemed complete. For clustering purposes, we used the following five bladder diary variables from those 193 women: number of intakes and voids, total volumes of intakes and voids, and maximum voided volume (serving as a proxy for bladder capacity).

In total, 185 variables were used for subtyping women with LUTS; 27 demographic variables, 55 medical history variables, 33 physical exam variables, 52 urinary symptoms variables (LUTS Tool and AUA-SI), 13 non-urinary PRO variables, and 5 bladder diary variables. Variables were continuous (n=83) or categorical (n=102). Supplemental Table S1 presents an overview of these variables for 545 women with LUTS used in the analysis. Although all variables were deemed important or possibly important in subtyping persons with LUTS, not all the variables are of equal importance, relevance, and non-redundancy; therefore, scaling and weighting of the variables was implemented as described below.

#### Data on non-LUTS controls

Our preprocessing procedure, described in more detail in the next section, includes scaling of variables by standardizing their values using means and standard deviations (SDs) in non-LUTS controls. The LURN study included 55 control women, who were not necessarily healthy but did not report LUTS. Unfortunately, not all the variables of interest were collected for these non-LUTS controls (e.g., physical examination, bladder diary). As a source of bladder diary data for non-LUTS controls, we used bladder diaries of 32 non-LUTS controls from the Establishing Prevalence of Incontinence (EPI) community study of women in Southeastern Michigan [58]. For other variables of interest not collected for non-LUTS controls, we used population data from literature sources indicated in Supplemental Table S1.

### Biological samples collected and analyzed in LUTS cases and non-LUTS controls

The LURN study collected numerous biological samples, including whole blood, serum, plasma, and urine at baseline and at 3- and 12-month follow-up visits [39-40]. Of these samples, 230 baseline serum samples of women with LUTS and 30 serum samples of non-LUTS controls were analyzed using the targeted proteomics approach – Proximity Extended Assay (PEA) by Olink Proteomics (Uppsala, Sweden). Three Olink panels (cardio metabolic, inflammation, neurology) were used to quantify abundances of 276 proteins. These data were not used for clustering in the current report; however, they served as an additional orthogonal approach for evaluation of the quality of the identified clusters. We compared the abundances of 276 proteins in women with LUTS and in controls and tested for significantly differentially abundant proteins in each of the identified clusters versus controls adjusted for multiple comparison using the false discovery rate (FDR) correction (FDR<0.05) [59]. Note that assays were performed in a subset (n =230, 42%) of women.

### Ethical guidelines and consent

The authors confirm all relevant ethical guidelines have been followed, and all research has been conducted according to the principles expressed in the Declaration of Helsinki. Informed consent has been obtained from participants. Institutional Review Board (IRB) approval has been obtained from: Ethical and Independent Review Services (E&I) IRB, an Association for the Accreditation of Human Research Protection Programs (AAHRPP) Accredited Board, Registration #IRB 00007807.

### Overview of the clustering pipeline

The clustering pipeline implemented in this paper contains multiple steps of data preprocessing, integration, clustering, and cluster evaluation. In this subsection, we present an overview of the sequence of steps in the pipeline shown in Figure 1. Details and rationale for each of the steps are provided in the rest of the subsections of the “Materials and Methods” section below. The pipeline is implemented using MATLAB (MathWorks, Natick, MA) and is publicly available through the Dryad repository (https://datadryad.org).

**Figure 1.**
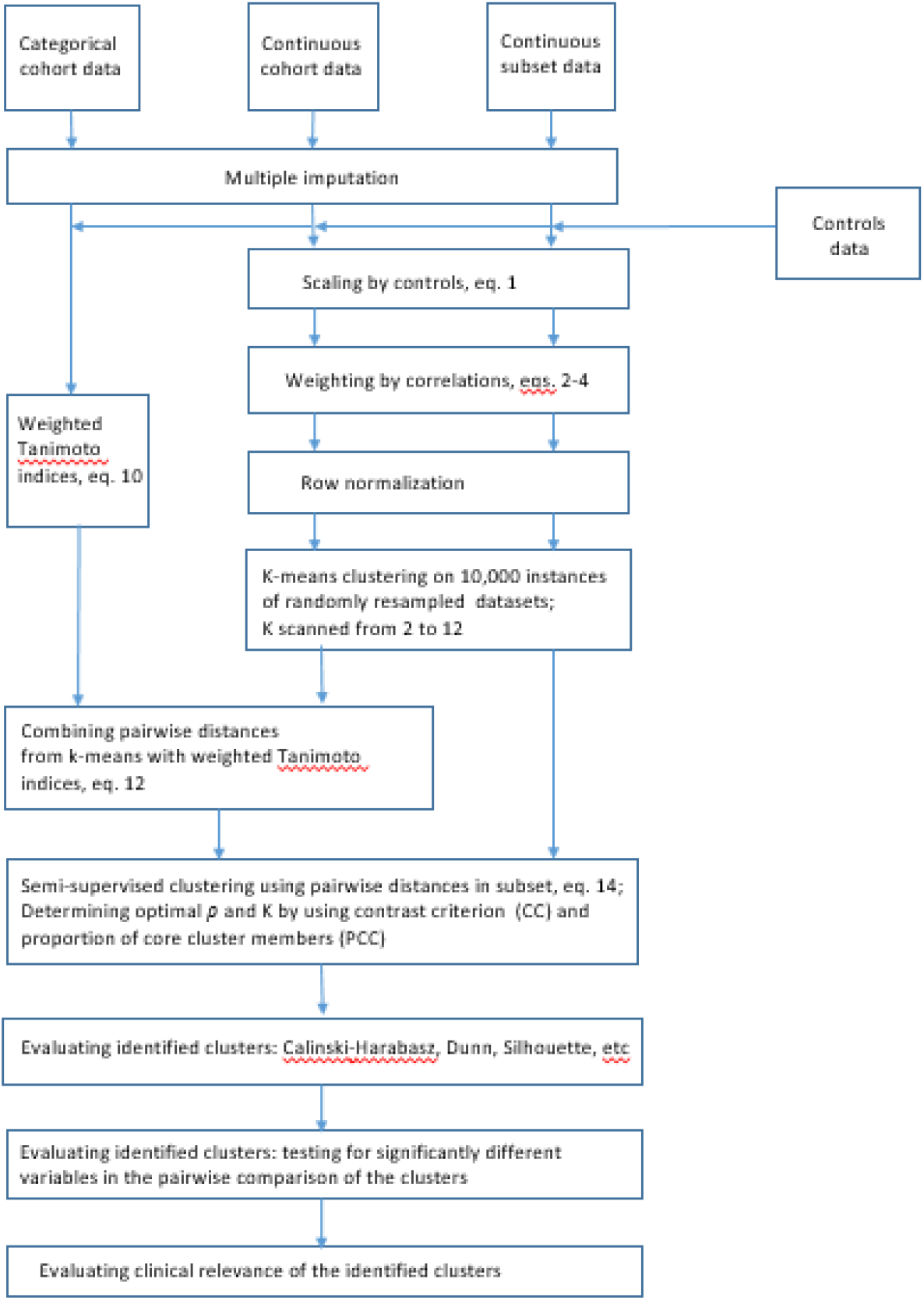
Flowchart of the pipeline for subtyping of a common complex disease or disorder by integrating heterogeneous data as used for subtyping of LUTS.

### Data preprocessing

#### Multiple imputation

Due to missing data (up to 10% in self-report questionnaires of urologic and non-urologic symptoms), multiple imputation was performed. The imputation used a sequential regression technique and was implemented using IVEware version 2.0 [60-61]. Ten imputed data sets were constructed, and each was preprocessed as described below. The k-means step of the resampling-based consensus clustering was performed on each of the data sets separately, resulting in ten pairwise probabilities of being in the same cluster for each pair of participants. Finally, hierarchical clustering on the mean of ten probabilities (pairwise distances) was performed to determine cluster membership.

#### Scaling of continuous variables

Scaling variables prior to clustering is an important and often overlooked step. Clustering algorithms group objects in a way that minimizes the sum of the pairwise distances between participants within the cluster, where the distance is composed of the distances between all variables calculated using Euclidian, Manhattan, or other suitable metrics [9]. Since each variable is measured using its own scale, the distances and optimal partition of the objects depend on these scales. As stated in [62], the problem with unscaled, unstandardized data is the inconsistency between cluster solutions when the scale of some of the variables is changed, which is a strong argument in favor of standardization. It is especially important in the case of heterogeneous data, where scales of variables in the raw data can be very different and completely unrelated. A common form of conversion of variables to standard scores (or z-scores) entails subtracting the mean and dividing by the SD for each variable. However, subtracting the cohort mean and dividing by the cohort SD would mask the subtype differences, since it ignores whether the within-cohort variance is caused by the natural biological variability of the subjects or by differences in disease subtypes, which increase the within-cohort variance due to multimodal distributions along certain variables. Using z-scores along such variables will unduly reduce their weight and will mask the presence of the subtypes. Therefore, standardization using z-scores is not suitable for our task of identifying disease subtypes. The solution to this problem is standardization by the mean and SD of a reference population that does not have the disease of interest, in this case, controls without LUTS. Following this approach, we define standardized variables *S*_*in*_ [63]:

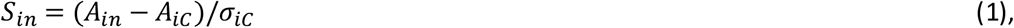

where *A*_*in*_ –*i*^th^ unstandardized variable for participant *n, A*_*iC*_-mean value of *i*^th^ variable across control subjects without disease of interest, *σ*_*iC*_ – SD of *i*^th^ variable in control subjects without disease of interest. A simulated example illustrating the benefits (substantially lower misclassification error) of clustering using variables standardized according to equation (eq.) 1, versus unstandardized variables and z-scores, is presented in Supplemental Material text.

#### Weighting variables to mitigate the redundancy

Clustering results can be skewed by including variables reflecting redundant information. An obvious and extreme case example will be including into the data set the same or highly correlated variables multiple times, which will result in the dominating role of these variables in the overall sum of squared distances, and therefore in the clustering decision. To mitigate this, we used weighting, so that the highest weight was attributed to the least correlated variable (i.e., the variable with the smallest average correlation with all other variables) and the lowest weight to the most correlated variable [41, 64]. The weights *w*_*i*_ were defined by equations (2-4):

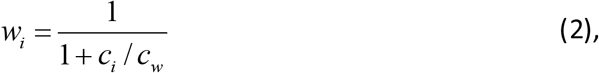

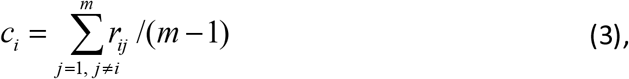

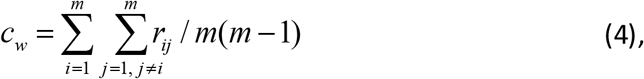

where m -number of variables and *r*_*ij*_ Pearson correlation coefficients of variables *i* and *j*.

#### Row normalization for continuous variables

Initial attempts to cluster un-normalized urinary symptoms data led to identification of two clusters that differed by overall severity of LUTS, not subtypes of symptoms. To avoid clustering predominantly by the overall severity of LUTS, in [41, 64] and here, we normalized the data by the participant’s overall severity of disease. For each participant, the weighted Euclidean length of the vector composed of all 78 continuous variables used for clustering was calculated as 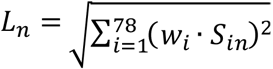, where *S*_*in*_ is the scaled *i*^th^ variable for the *n*^th^ participant and *w*_*i*_ is the weight of *i*^th^ variable defined by eq. (2-4). Each continuous variable was then normalized by *L*_*n*_, the Euclidean length of the participant’s vector, resulting in normalized continuous variables 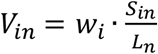. This normalization strategy allowed for clustering based on the direction rather than the length of the vector representing each subject.

### Consensus clustering using continuous variables

Clustering was performed using a resampling-based consensus clustering method introduced by Monti et al [34]. We performed 1000 instances of random resampling, each selecting a subset including 80% of participants. The same procedure was repeated for each of the ten multiply imputed data sets, resulting in 10,000 subsets. We then partitioned each of the subsets into clusters using a k-means clustering algorithm implemented as *k-means* MATLAB function (with option ‘number of replicates’ =8, see [63] for the explanation on the need of this option); with number of clusters K scanned from 2 to 12. Let *Q*_*nq*_ denote the number of times participants *n* and *q* were assigned by k-means into the same cluster. Let *I*_*nq*_ denote the number of times participants *n* and *q* were both selected in the random sampling. The probability of participants *n* and *q* belonging to the same cluster could be calculated as *Q*_*nq*_ /*I*_*nq*_. We could thus obtain ten probabilities for a pair of participants from the ten imputed data sets. The average of these probabilities represented the final consensus index *M*_*nq*_for participants *n* and *q*. A 545 by 545 consensus matrix *M* (Figure 2) was constructed to visualize these average probabilities as a heat map. Probability is color-coded: bright yellow represents probability close to one and dark blue probability close to zero. The indices of participants were reordered so that the participants belonging to the same clusters were grouped together. To reorder the indices of participants in the consensus matrix, we employed hierarchical clustering (using *clustergram* MATLAB function) with 1 − *M* as distance matrix so that participants belonging to the same clusters were grouped together, depicted as bright yellow blocks along the diagonal of consensus matrix.

**Figure 2.**
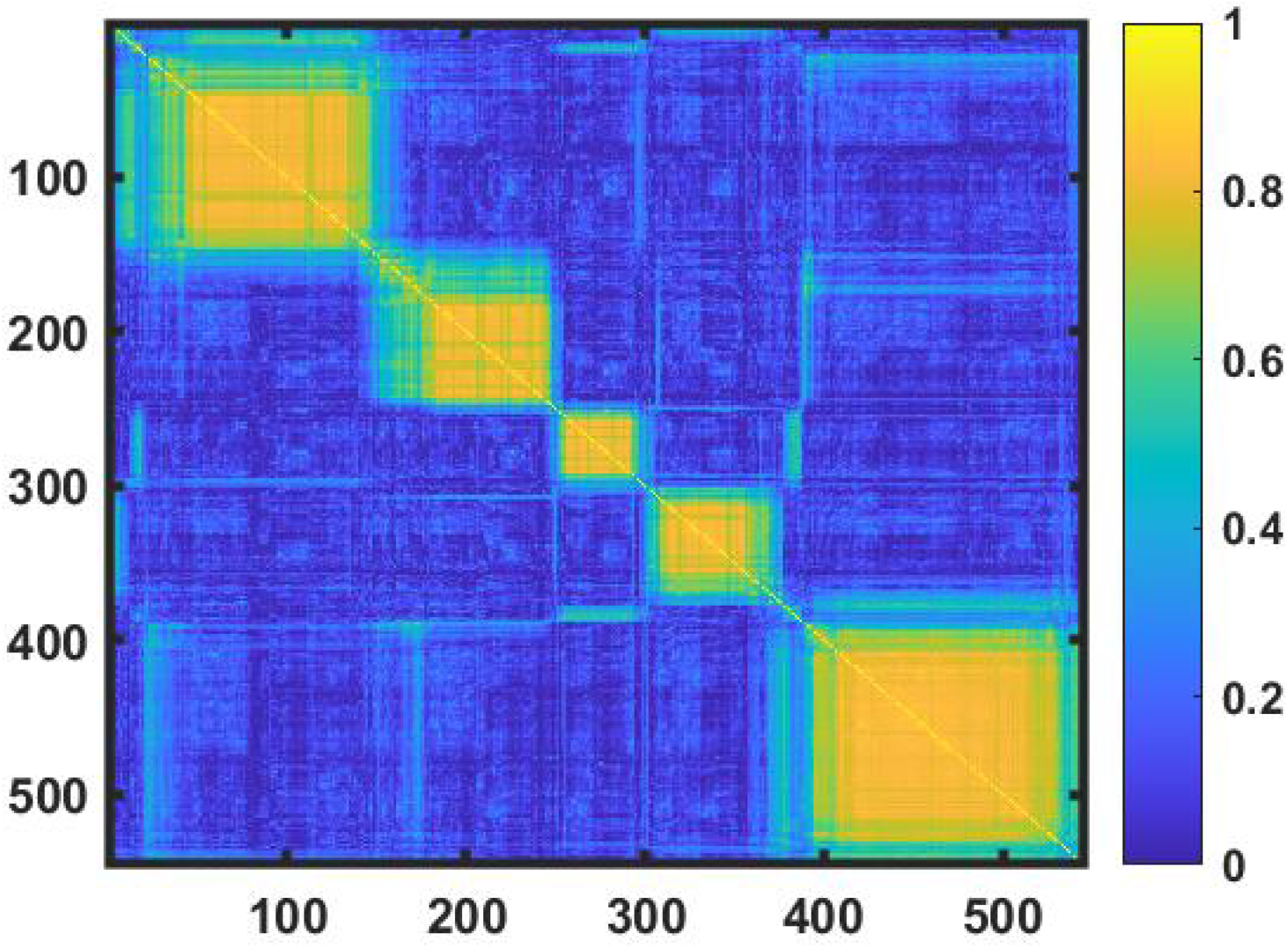
Consensus matrix. Consensus matrix is presented as a heat map, where the probabilities *M*_*nq*_for each pair of participants to be in the same cluster are shown by color-coded elements; bright yellow represents probability close to one and dark blue probability close to zero.

### Clustering using categorical variables

Of 185 variables used for clustering women with LUTS, 102 (55%) are categorical. K-means is not an appropriate method for categorical variables, so resampling-based consensus clustering with multiple runs of k-means algorithm, which we used for continuous variables, cannot be directly used for categorical variables.

#### K-prototype approach

One way to combine continuous and categorical variables is to use the k-prototype algorithm introduced by Z. Huang [65]. According to [65], the distance between two objects *X,Y*, described by *p* continuous variables and *m-p* binary variables, is represented as:

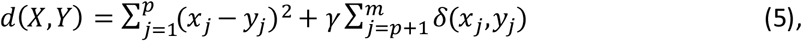

where δ is Kronnecker symbol describing simple matching and γ is the weight introduced in [65] to “avoid favoring either type of attribute” (continuous vs. categorical). The goal of the k-prototype algorithm is to minimize the sum of the distances (defined by eq. 5) between objects within the cluster. Limitations of the k-prototype algorithm include lack of scaling of categorical variables and use of the same weight γ for all categorical variables, regardless of their relevance to the disease of interest or their redundancy. A later version of the algorithm [66] attempted to overcome some of these limitations by defining the distance between an object *X*_*n*_and the centroid *Z*_*k*_ of the *k*^th^ cluster as:

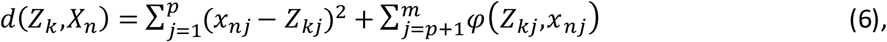

where 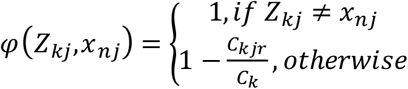, and *C*_*k*_ is the number of objects in cluster k, while *C*_*kjr*_ is the number of objects in this cluster with the categorical value 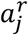 of the *j*^th^ attribute, e.g., participants with blue eyes in cluster k. Such definition of distance makes sense; for instance, if the number of participants in cluster k is 100, and 51 of them have blue eyes, then for a person with brown eyes, distance from the centroid along this dimension is 1, but for a person with blue eyes, it is 1-0.51=0.49. Now, if 99 participants have blue eyes, then for them, the distance from the centroid is 1-0.99=0.01, while for the only one with brown eyes, it is still 1. Thus, such a definition of distance makes certain attributes more important (defining) for the cluster if the majority of objects have the same value of this attribute. An algorithm using such a definition of distance between objects would strive to make clusters as homogeneous as possible with regard to both its continuous and categorical variables. This approach, however, does not distinguish between categorical variables relevant and irrelevant to the disease of interest.

To distinguish between relevant and irrelevant variables, we suggest using the same approach as for continuous variables, i.e., compare them with controls without the disease or symptom complex of interest. For the categorical variables transformed into binary variables, we suggest scaling by comparison of the frequencies of these binary variables in LUTS *F*_*j*_and in controls *F*_*jC*_ by using the following function:

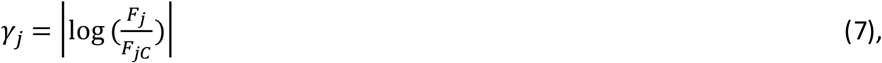

where |*x*| is absolute value of *x*.

If, for a certain binary variable, frequencies in LUTS and non-LUTS controls are equal, e.g., prevalence of blue eyes is the same in LUTS and non-LUTS, then this variable will get weight *γ*_*j*_ = 0 and would not affect clustering decision, essentially excluding the variable from clustering. However, if the frequency of this variable in LUTS is higher or lower than in controls, then *γ*_*j*_ > 0, and this, relevant to disease variable, will affect clustering decisions. To accommodate this scaling together with weighting of the variables based on their correlation with other variables described by eq. (2-4), one needs to modify eq. (6):

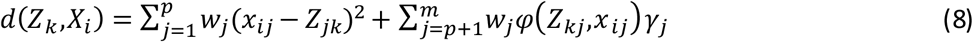

Unfortunately, none of the available implementations of k-prototype algorithm in standard software (R and SAS [67-68]) easily allows for such modification, and therefore, an alternative simpler approach was used.

#### Weighted Tanimoto indices approach

A simpler approach to clustering categorical variables is based on Tanimoto indices or Tanimoto similarity measure [69]. For two objects *a* and *b* described by *m* binary variables, Tanimoto similarity is defined as:

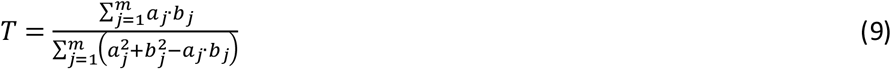

For instance, if *a* and *b* are 5-dimensional binary vectors *a* = [1, 1, 0, 0, 0] and *b* = [1, 0, 1, 0, 0], then *T*=1/(2+2-1)=1/3. Note that common “ones” but not common “zeros” are counted in this definition of similarity, which is especially useful in case of multiple binary variables formed from one categorical variable. Think, for example, of the categorical variable ‘eye color’ transformed into several binary variables: ‘blue eyes’ (yes, no), ‘brown eyes’ (yes, no), ‘green eyes’ (yes, no), etc. Tanimoto similarity between two persons with blue eyes will not depend on whether you add ‘hazel eyes’ to the list of options or not.

Not all of the categorical variables are equally relevant to the disease of interest, so we want to be able to assign weights reflecting the level of relevance for each variable by comparing its frequency in LUTS with its frequency in non-LUTS controls. We also want to compensate for redundancy in the variables by using weights defined by equations (2-4). Importantly, we want to make sure that no categorical variable, even if it is much more frequent in disease than in controls, has overwhelmingly high weight and makes the role of differences in other variables negligible. To attain this goal, we introduce a weighed Tanimoto similarity measure as:

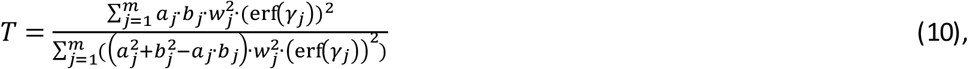

where *w*_*j*_ is the weight defined by eq. (2-4), using the appropriate correlation coefficients of the variables. Coefficient *γ*_*j*_ is defined by eq. (7) and minimizes the role of binary variables equally prevalent in disease and controls. Function 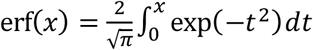 ensures that the weight of each binary variable is smaller or equal to one, even for the high values of *γ*_*j*_, since |erf (*x*)| ≤ 1. Note that maximum value of *T* is equal to one when all the categorical variables in *a* and *b* are the same, and minimum value is zero when all the categorical variables are different. Now we can use eq. (10) to define distance between any pair of participants described by *J* binary variables as:

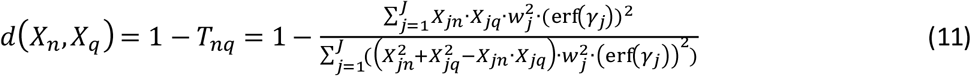

### Combining continuous and categorical variables

Note the similarity of the pairwise distance 1 – *T*_*nq*_ between two participants based on the categorical variables describing them (eq.11), and pairwise distance 1 – *M*_*nq*_ between these participants based on their continuous variables. The former is equal to zero when all the categorical variables describing these two participants are the same, while the latter is equal to zero when the two participants always were assigned to the same cluster by the 10,000 instances of k-means in the resampling-based consensus clustering. Similarly, the former is equal to one when all the categorical variables describing the two participants are different, while latter is equal to one when these participants always were assigned to the different clusters in resampling-based consensus clustering. This similarity allows combining continuous and categorical pairwise distances into a single distance measure *D*_*nq*_, with a minimum value of zero and maximum value of one using the weighted Euclidean length approach:

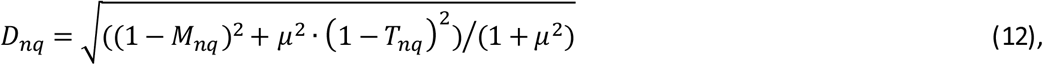

where *μ* is the weight representing the relative role of the distances based on categorical variables and on continuous variables. We set it equal to the ratio of the number of non-redundant categorical and continuous variables: 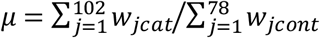, where *w*_*j*_ are determined by eq. (2-4) using correlation coefficients appropriate to the distributions of the variables.

### Semi-supervised clustering using bladder diary data

Of 545 women in the Observational Cohort of LURN, 193 (35%) returned complete bladder diaries without missing volumes of voids and intakes. These data were deemed clinically important to the subtyping of women with LUTS. One way to integrate data domains only available on a subset is by using semi-supervised clustering methods [70-71]. If class membership for the members of the subset is known, then the objective function in the clustering of the whole cohort should be modified as follows:

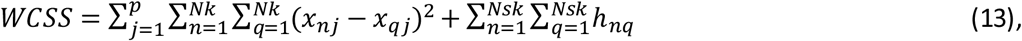

where *WCSS* - within cluster sum of squared distances, *p* – number of variables, *n* ≠ *q, Nk* - number of cohort participants in the given cluster k, *Nsk* - number of the participants in cluster k that were present in the subset. The values of *h*_*nq*_ = −*h* are negative (reward, decreasing the within-cluster-sum-of-squares [*WCSS]*) if participants *n* and m belong to the same cluster according to subset classification, and is positive *h*_*nq*_ = *h* (punishment, increasing *WCSS*) if participants *n* and *q* belong to the different classes of the subset. This approach known as “must-link, cannot-link” allows for using labels known from classification of the subset to influence clustering of the cohort. The limitation of this approach, however, is that it does not allow for different level of confidence in subset cluster membership, i.e., participants *n* and *q* are either in the same subset cluster or not. In our case, additional data available for the subset of participants do not provide 100% confidence in cluster membership for this subset; furthermore, the number of participants in the subset is lower than in the whole cohort, making the subset clusters less robust. There is a measure of similarity, however, that is quantitative and reflects the confidence in cluster membership; it is the pairwise distance between members of the subset. We suggest using this measure to modify the pairwise distance defined by eq. 12 by taking into account the similarity between members of the bladder diary (BD) subset:

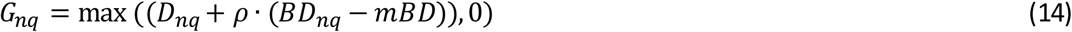

where *D*_*nq*_ is defined by eq. 12, *BD*_*nq*_is the pairwise distance between members of the subset, *mBD* is the mean pairwise distance between members of the subset, *ρ* is a parameter determined as described in the below sections. If either participant *n* or *q*, or both of them are not members of the subset, their pairwise distance is not known and is assumed equal to the mean pairwise distance within the subset *mBD*. For these participants, the second term of eq. 14 is equal to zero; while, for members of the subset, it is either negative or positive depending on whether *BD*_*nq*_ is smaller or larger than *mBD*. Note that, since the second term might be negative, for some large values of *ρ*, the sum of the two terms is negative as well. However, the pairwise distance between the objects cannot be negative, and is therefore set to zero for these cases.

We used five variables (number of intakes and voids, total volumes of intakes and voids, and maximum voided volume) from the bladder diaries of the 193 participants to refine the values of their pairwise distances. These five variables were scaled using bladder diary data for controls [58] and then added to the 78 other continuous variables to calculate pairwise distances *BD*_*nq*_ refined with bladder diary variables, as described in the subsection on consensus clustering using continuous variables. Then it was introduced into eq. 14 to get the refined matrix of the pairwise distances *G*_*nq*_. The value of the coefficient *ρ* was determined by optimizing the quality of clusters, as defined in the following subsections.

### Determining the number of clusters

Determining the number of clusters is an important step in any clustering process. In partitioning algorithms like k-means, it is necessary to decide on the number of clusters *K* prior to running the algorithm. In agglomerative algorithms like hierarchical clustering, it is possible to decide on the number of clusters when the dendrogram based on the distances between objects is already created. It is common to try several values of *K* and then to compare the resultant clusters by using various quality of clustering criteria, including Calinski-Harabasz, Davies-Bouldin, Dunn, Gap, and Silhouette indices [22-26]. Quite often, these criteria disagree on the value of *K* that optimizes the quality of the clusters.

Resampling-based consensus clustering, introduced in [34] and subsequently applied to our data set, is a combination of multiple instances of k-means clustering followed by hierarchical clustering on the pairwise distances between objects derived at the first stage. The value of *K* in k-means is typically scanned (in our case from 2 to 12), and then the optimal value of *K* is determined using criteria developed specifically for consensus clustering algorithm [34-35, 47]. In both [34] and [47], the determination of the number of clusters is based on analysis of the cumulative distribution function (CDF) of consensus index values *M*_*nq*_ (defined in the “Consensus clustering using continuous variables” subsection of this paper). The main idea of this analysis is that, in case of ideal clustering, there are only two possible values of consensus index *M*_*nq*_ = 1, when a pair of objects *n* and *q* are in the same cluster, and *M*_*nq*_ = 0, when they are in the different clusters. Therefore, the ideal CDF should consist of two vertical lines and a horizontal (flat) line between them. The length of the first vertical line will be equal to the number of pairs with *M*_*nq*_ = 0 and the length of the second vertical line to the number of pairs with *M*_*nq*_ = 1. However, if the value of *K* used in clustering is different from the true value of *K*, then the shape of the CDF curve differs from the idealized curve described above. In [34], the optimal *K* is defined as the one for which the change in the area under the CDF (relative to area at *K-1* and *K+1*) is the largest (“elbow” of the AUC vs. K curve). Analysis in [47] demonstrates several examples when the criterion of [34] does not work and suggests an alternative criterion named proportion of ambiguously clustered pairs (PAC) equal to the number of pairs with 0 < *M*_*nq*_ < 1 over the total number of pairs. Obviously, in the real-world situation of biological variability and noisy measurements, almost all of the pairs will fall into this category, so some more liberal lower and upper boundaries for *M*_*nq*_ need to be introduced, e.g., 0.1 and 0.9, as in [47]. It raises a question, however, whether these boundaries should be different for different values of *K*, since potential ambiguity increases with the increased number of clusters.

A more straightforward approach to determine the number of clusters is used in [35] by introducing mean consensus score (CS) calculated as the mean value of consensus indices *M*_*nq*_ within the clusters. The value of *K* that results in the highest CS is considered optimal. The problem with CS is that it favors the high number of clusters, e.g., CS could reach its maximum when K=N/2 and each cluster contains only a pair of most similar objects with highest values of *M*_*nq*_.

Below, we introduce a pair of complementary criteria, i.e., contrast criterion (CC) and proportion of the core clusters (PCCs), which we used in our clustering pipeline. We believe they combine the advantages of PAC and CS and are free of some of their limitations, especially when used as a complementary pair. The below sections introduce the CC and PCCs based on the analysis of consensus matrix derived by k-means clustering of the multiple resampling instances of the data set; however, these criteria can be used with any matrix of pairwise distances between the objects.

#### Contrast criterion

The idea of the CC is derived from visual representation of the consensus matrix as a heat map presented in Figure 2. Each pixel of the heat map represents the value of *M*_*nq*_ probability of two objects to be together in the same cluster. Each bright yellow square along the diagonal of the matrix represents a cluster. Next, we compared the “bright yellowness” of this diagonal square with the “color” of the rest of the row in which the diagonal square is located; therefore, the term “contrast criterion” (CC). The larger the difference between these two measures, the further the situation from the case where all *M*_*nq*_ except for *n=q* are equal, and the heat map is uniformly yellowish (single uniform cluster case), in which case the contrast is zero. We consider number of clusters K and cluster membership optimal when CC is maximized.

When analyzing the properties and behavior of clustering criteria, it is necessary to compare the clusters identified using certain clustering algorithms and clustering criteria of interest with the “true” clusters. Unfortunately, “true” clusters are not known in real life, and therefore, one needs to simulate them and then evaluate misclassification error resulting from the clustering algorithm and criteria of interest. Such an approach was used in [63] to compare several popular clustering algorithms, and was applied for the case of resampling-based consensus clustering with CC and PAC criteria (see Supplemental Material text and Supplemental Figures S2-S10). Here, we concentrate on the general analysis of CC and its properties. In this analysis, we need to introduce the term “alleged clusters”, which are different from “true clusters” and “identified clusters”. “Alleged clusters” are determined for each value of K tried by clustering algorithm, while “identified clusters” are those maximizing the value of clustering criteria of interest, and “true clusters” are specified by the simulation.

To define CC explicitly, let us first look at the most typical case where the number of alleged clusters is not equal to the number of objects, is not equal to one, and none of the clusters includes just one object. Note that we are not making any assumptions or imposing any restrictions on the properties of the “true” clusters.

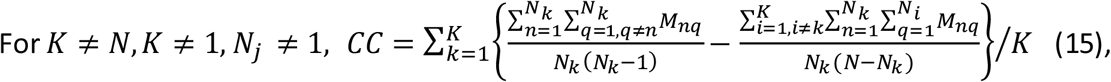

where *K* is the number of alleged clusters, *N* is the number of the clustered objects, *N*_*k*_ and *N*_*i*_ are the numbers of objects in *k*^th^ and *i*^th^ clusters. As intended, the first term in the brackets of eq. (15) represents the average “bright yellowness” of the diagonal square, while the second term in the brackets represents the average “yellowness” of the rest of the row in which the diagonal square is located.

Note that we defined CC as an averaged contrast across alleged *K* clusters independently of the size of the clusters. Other approaches are possible, e.g., a weighted average based on the sizes of the clusters, or minimax approach, where the contrast for the “worst” (least contrast cluster) is maximized. Clearly, the best choice of combining contrasts of each of the clusters into one overall depends on many factors, including the goal of the clustering, expected sizes of the clusters, and the number and distribution of the variables. It is an interesting topic; however, it is outside of the scope of this paper.

Another choice made in defining CC by eq. (15) is omitting of the diagonal terms *M*_*nn*_, which are always equal to one. The percentage of diagonal terms in each square representing an identified cluster is 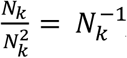, or 100%, 50%, 33%, 25% for *N*_*k*_ = 1, 2, 3, 4. Since *M*_*nn*_ = 1, *M*_*nq*_ < 1, inclusion of diagonal terms in the definition of CC would favor smaller clusters over larger clusters by assigning higher CC to the smaller clusters, irrespective of similarities of objects within the clusters. Therefore, we do not include diagonal terms in the definition of CC, as shown in eq. 15. Note that eq. (15) does not work if *K=1, K=N*, or *N*_*j*_ = 1. For these special cases, CC is defined and discussed in Supplemental Material text.

There are certain similarities between our contrast criterion *CC* and consensus score (CS) of [35]. Although the exact definition of CS is not provided, it appears that CS is similar to the first term of eq. (15). However, it is unclear if the diagonal terms *M*_*nn*_ are omitted or included and how CS for *K* clusters are combined to derive the overall CS. Nevertheless, it is of interest to compare the behavior of CS and CC in some simple idealized cases. In case of ideal clustering, both CS and CC reach their maximum possible value of 1. The worst-case scenario for both criteria is the case where *K>1* clusters are alleged, when in reality, there are no “true” clusters, and all objects are described by a unimodal random distribution of their attributes (variables). Now, if the number of alleged clusters *K* >1, there is an equal probability for the object to end up in any of *K* clusters, so the mean value of *M*_*nq*_ is *1/K*, which makes the minimal possible value for CC equal to zero and minimal possible value for CS equal to *1/K*. The rather limited range of values from *1/K* to 1, together with the dependence of the minimal value on the number of alleged clusters, are the limitations of the CS criterion, which are absent in case of CC, where the range is from 0 to 1 irrespectively of *K*. Therefore, we consider the use of CC advantageous for determining the optimal number of clusters.

#### Core clusters

When analyzing the quality of alleged clusters, it is important to know the confidence with which cluster membership is determined. Clearly, one would prefer clusters where the probability of objects to belong to a particular cluster is 0.9 rather than 0.3. The knowledge of consensus matrix allows for calculating the probability for each object *n* belonging to a particular cluster *k*:

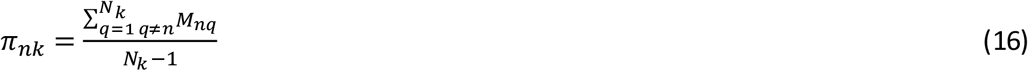

Note that it is different from the probability averaged across the cluster that was used to calculate CS and CC in the previous subsection. Importantly, within the same cluster, some objects might have probability (confidence) as high as *π*_*nk*_=0.9999…, or as low as *π*_*nk*_ = 1/*K*+0.001, assuming that it is lower for any other cluster *i* ≠ *k*. We will call the *n*^*th*^ object the core member of cluster *k* if *π*_*nk*_ > 0.5, which means that, for this object, the probability to be in cluster *k* is higher than probability to be in all other clusters combined. The rest of the members of the cluster do not belong to the core; for them, the probability to belong to the *k*^*th*^ cluster is just higher than to be in any other given cluster. The number of core members divided by the total number of objects in the cluster provides a useful measure that we named proportion of the core cluster (PCC). As with the contrast criterion CC, one can use several approaches to derive overall PCC from the PCCs for each cluster, i.e., take the average across all clusters, weighted average based on the size of the clusters, or minimax by looking at the PCC in the worst cluster. PCC provides a measure of overall confidence in the alleged cluster membership and of the uniformity of the alleged clusters. Unlike contrast CC, PCC favors smaller size of the clusters and reaches its maximum when K=N/2 and each cluster contains just a pair of objects. PCC reveals information similar to the proportion of ambiguously clustered pairs (PAC) [47], for which 0.1 < *M*_*nq*_ < 0.9; however, PCC is easier to interpret and does not include unjustified upper and lower boundaries 0.1 and 0.9.

#### Using CC and PCC to determine optimal number of clusters K

We used a combination of CC and PCC to determine the optimal number of clusters K. Note that eq. 14 contains undefined coefficient *ρ*. Therefore, we have two parameters *ρ* and K to determine and two criteria to meet. We determined *ρ* and K as the values maximizing CC and corresponding to an elbow (point of diminishing returns) for PCC. A clustering procedure was run for 24 values of *ρ* from 0.05 to 1.2 using the single-program multiple data sets (spmd) function of MATLAB Parallel Computing Toolbox; K values were scanned from K=2 to K=12. The determined optimal values were *ρ* =0.3 and K=5. The resultant consensus matrix together with the values of CC and PCC for K scanned from 2 to 12 are presented in Figure 3. As seen in Figure 3, contrast criterion CC and percent of core cluster members PCC both have maxima for number of clusters K=5. As shown in Table 1, other quality of clustering criteria also confirm K=5 as an optimal number of clusters for our cohort of women with LUTS.

**Table 1.** Other quality of clustering criteria, confirming K=5 as an optimal number of clusters.

|  | K=2 | K=3 | K=4 | K=5 | K=6 | K=7 | K=8 | K=9 | K=10 |
| --- | --- | --- | --- | --- | --- | --- | --- | --- | --- |
| Calinski-Harabasz (↑) | **1382.26** | 346.13 | 676.29 | **1126.25** | 567.83 | 398.02 | 309.93 | 234.52 | 180.57 |
| Davies-Bouldin (↓) | **0.4328** | 0.8175 | 0.6045 | **0.4720** | 0.5692 | 0.9178 | 1.0809 | 1.1498 | 1.1678 |
| Dunn (↑) | 0.0167 | 0.0072 | 0.0290 | **0.0485** | **0.0496** | 0.0455 | 0.0273 | **0.0754** | **0.0590** |
| Point-Biserial (↓) | **-4.801** | -2.728 | -3.25 | **-3.849** | -2.515 | -2.081 | -1.855 | -1.621 | -1.409 |
| Silhouette (↑) | **0.6896** | 0.4565 | 0.5731 | **0.6857** | 0.5955 | 0.4821 | 0.3723 | 0.3143 | NaN |

**Figure 3.**
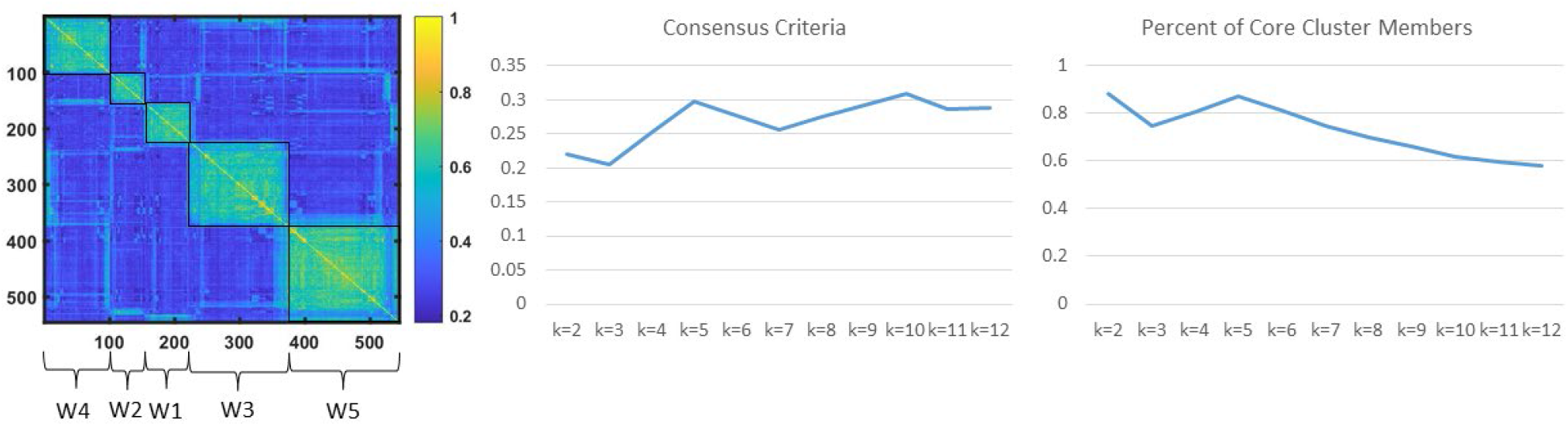
Determination of the optimal number of refined clusters. (A) Consensus matrix heat map demonstrates five clusters of participants (named W1-W5) grouped together based on the pairwise distances *G*_*nq*_ (eq 14). (B) CC eq. 15 for K=2,…,12. (C) PCC eq. 16. Both CC and PCC have maxima at K=5.

### Evaluation of the quality of the identified clusters

We used multistep procedure to evaluate the quality of identified clusters and compare with the potential alternative clusters. The first step was examination of CC and PCC for each of K clusters (Figure 3). Next, we calculated other established quality of clustering criteria, including Calinski, Davies-Bouldin, Dunn, Point-Biserial, Silhouette [22-26], etc. (Table 1). Then, we performed pairwise comparison of the clusters using Wilcoxon rank sum tests and chi square tests, where appropriate, to determine which variables used for clustering were significantly different in the pairwise comparison. All pairwise comparisons were adjusted using an FDR correction for multi-testing [59].

### Visualization of the results

We used several tools to visualize the results of our analyses. Heat maps representing the consensus matrices were generated using the clustergram MATLAB function. Properties of the identified clusters were illustrated by radar plots, built-in using SAS statistical graphics panel (sgpanel) scatter, polygon, and vector procedures. Comparison of cluster membership in the refined clusters versus previously identified symptom-based clusters was performed using Sankey diagrams, built with the googleVis package.

## RESULTS AND DISCUSSION

### Description of the clusters

Five distinct clusters of women with LUTS were identified by clustering 545 participants of the LURN Observational Cohort Study using 185 variables. We call these clusters W1-W5, in order to distinguish them from clusters F1-F4, described previously [41]. Demographic data for each cluster are presented in Table 2. Some demographic characteristics were different across the clusters, including age, ethnicity, menopausal status, obesity, prevalence of hysterectomy, percentage of participants with at least one vaginal birth, education level, and employment status. No significant differences across the clusters were observed for race, smoking, and alcohol use.

**Table 2.**
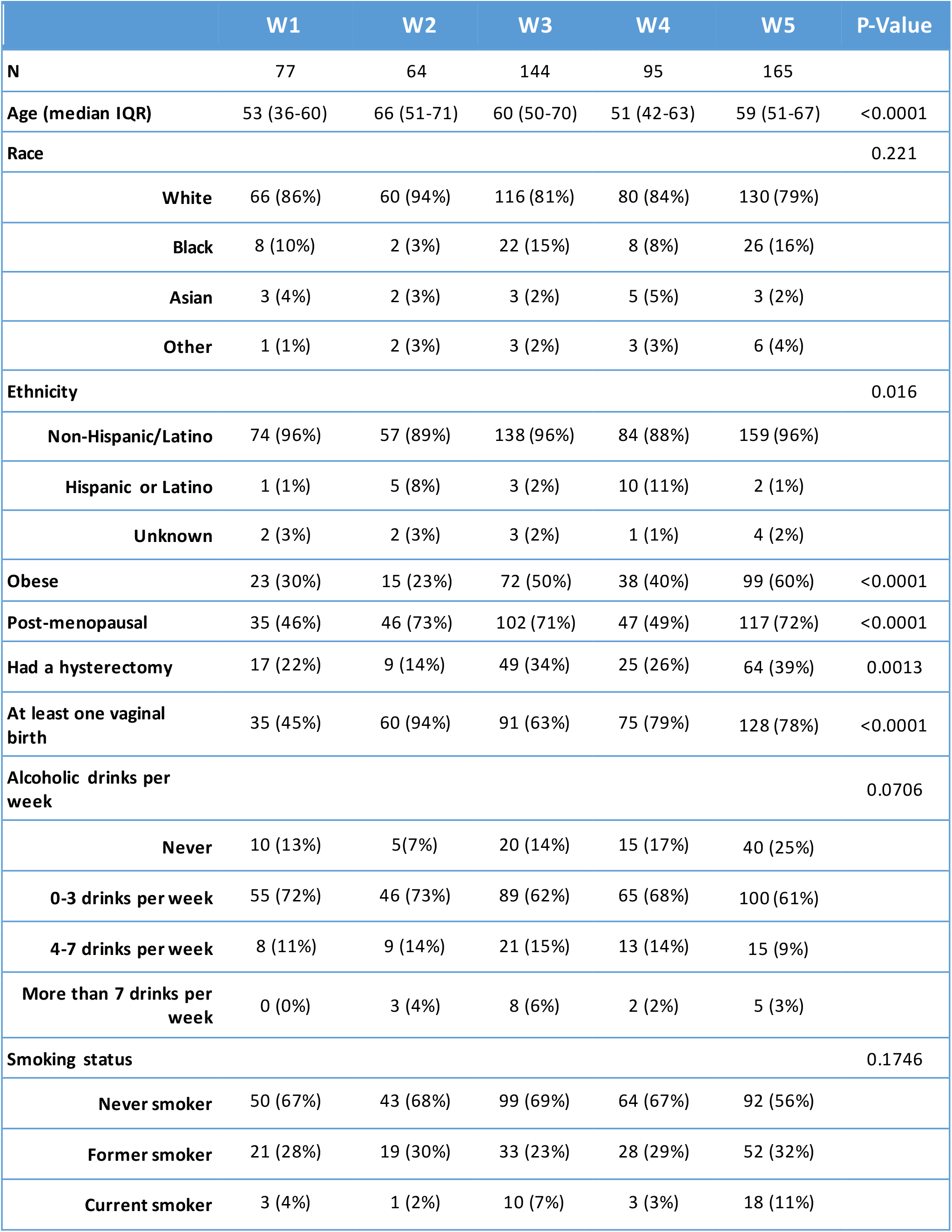
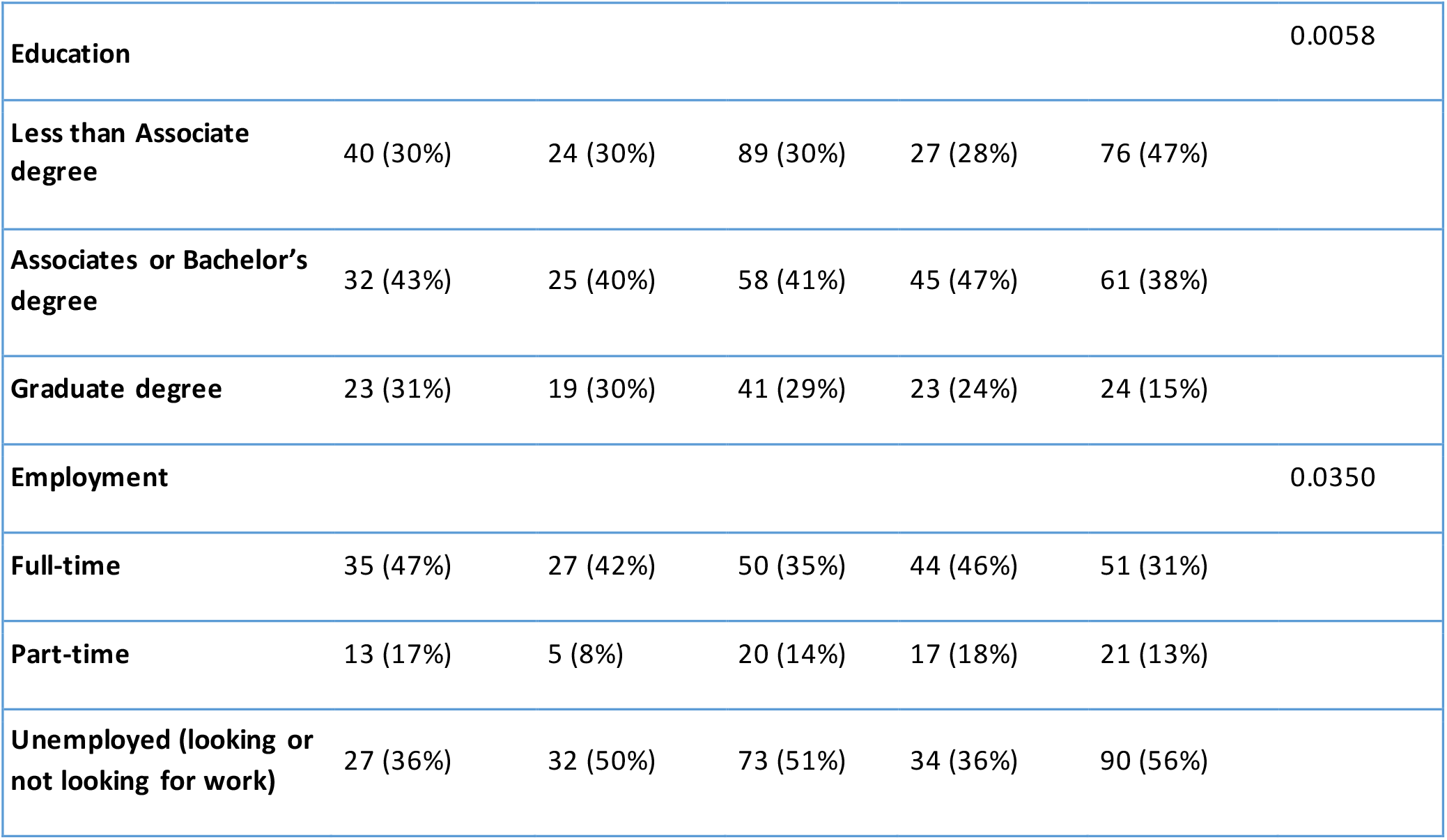
Demographic Data for clusters W1-W5.

Importantly, all urinary symptoms and many other clinical variables were significantly different across the clusters. Table 3 presents the comparison of urinary symptoms (collected with LUTS Tool and AUA-SI), bladder diary variables, and other 37 significantly different clinical variables across clusters W1-W5. For clarity, we describe these significantly different variables while discussing signatures of the clusters in the text following Table 3. Table 2, and especially Table 3, illustrate the distinctiveness of the identified five clusters of women with LUTS.

**Table 3.**
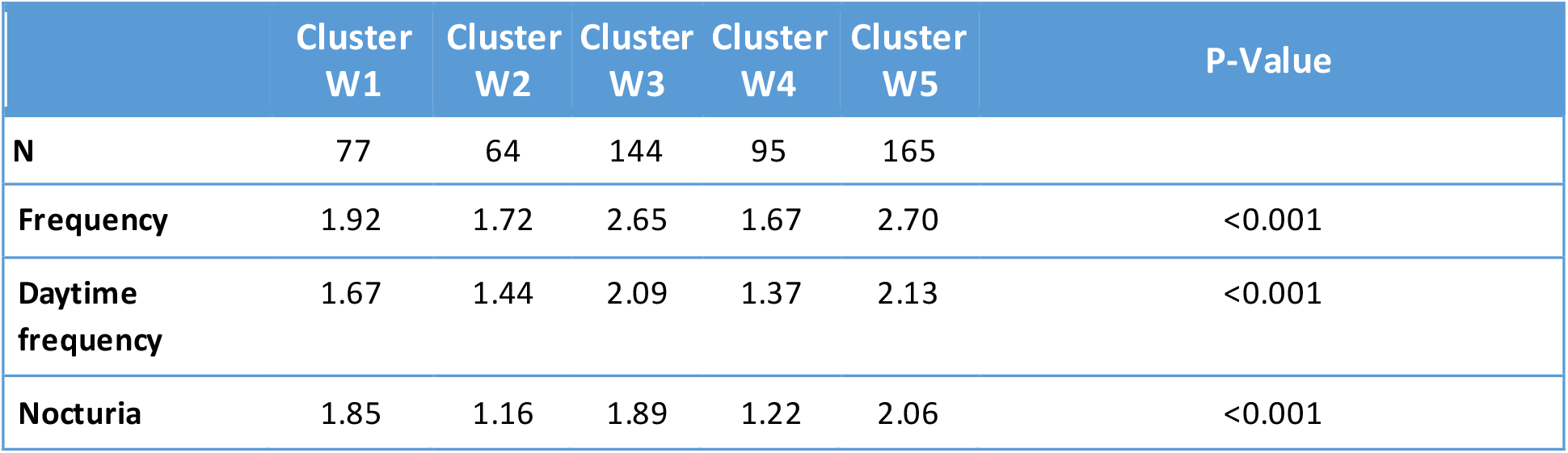
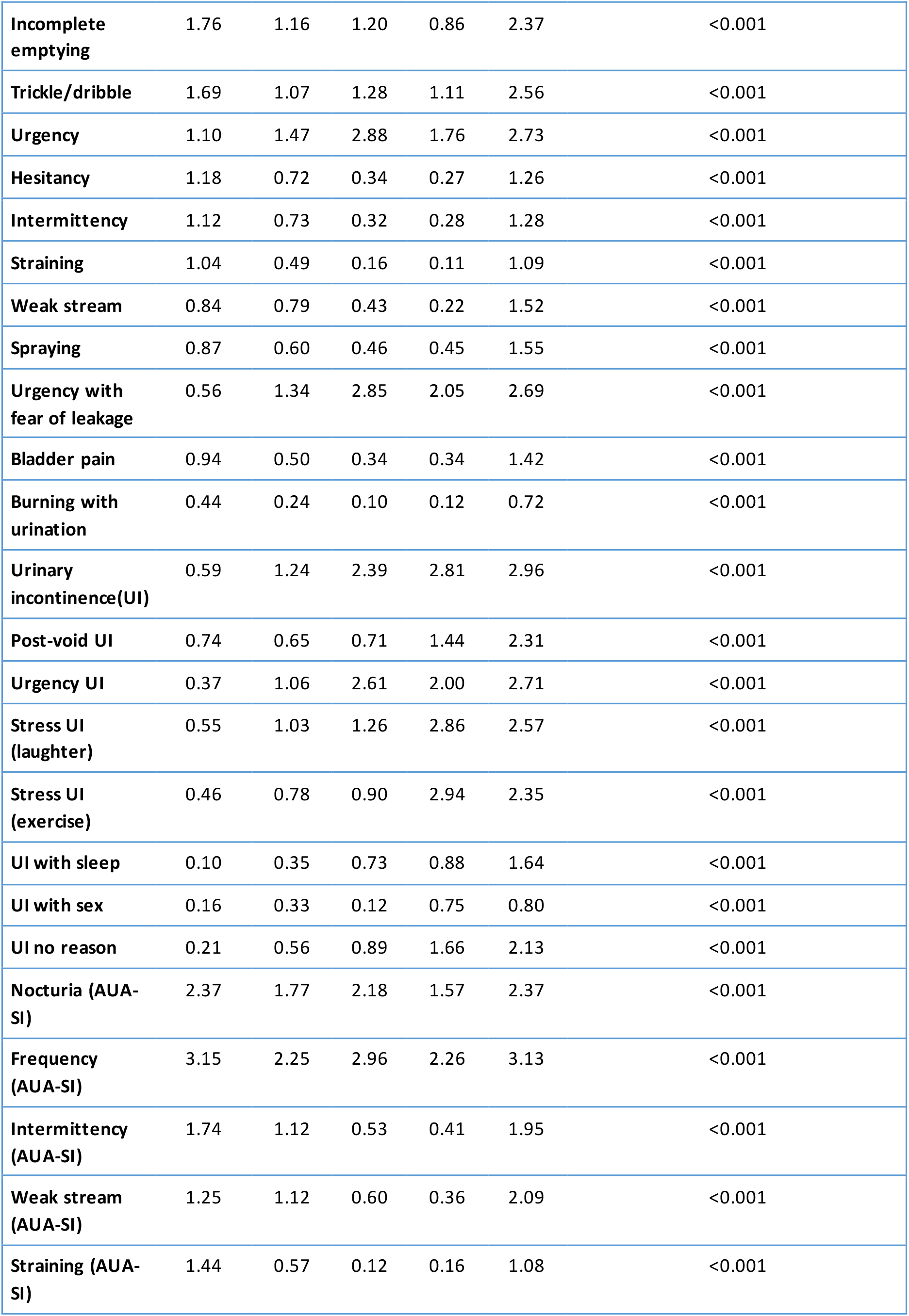
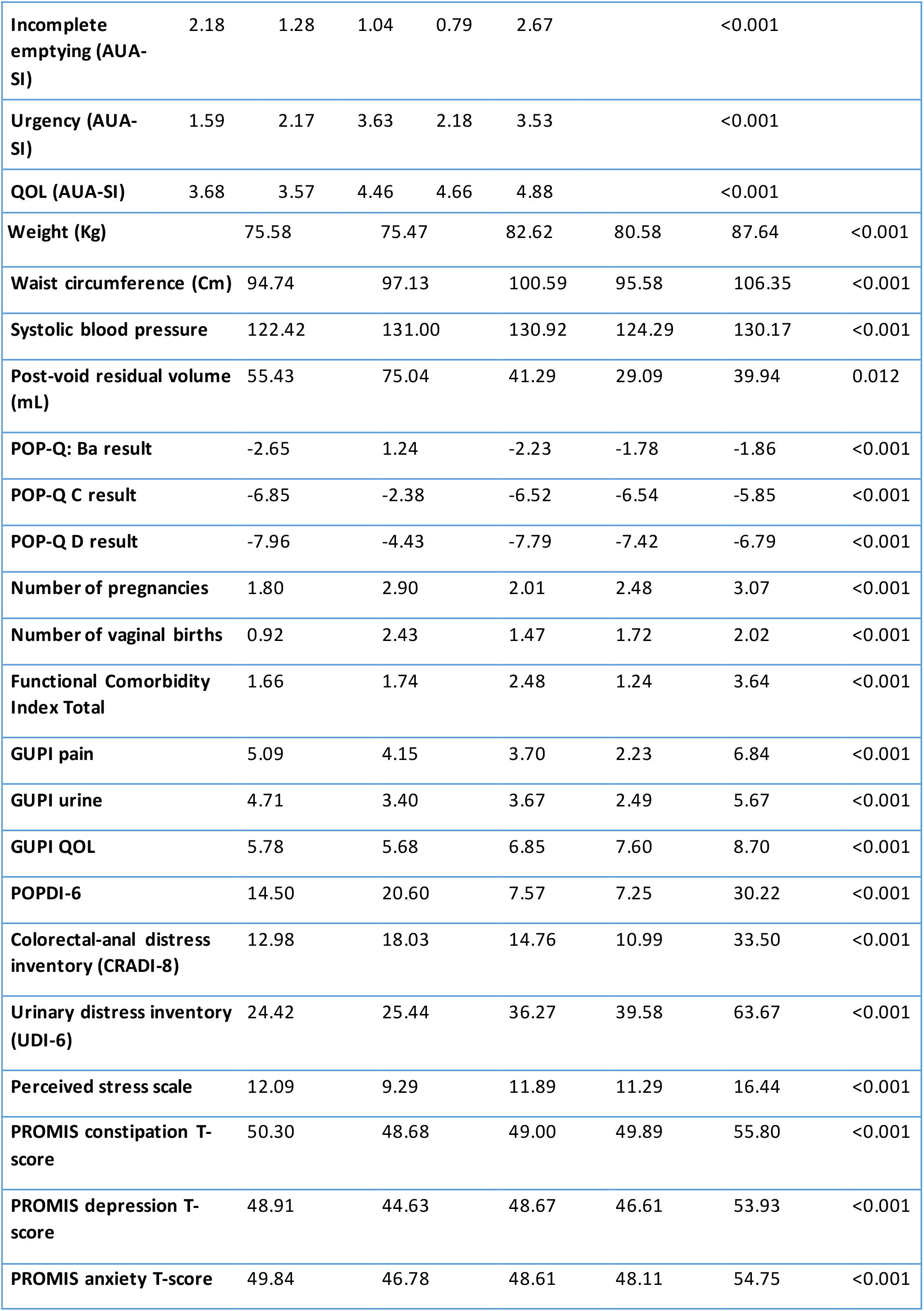
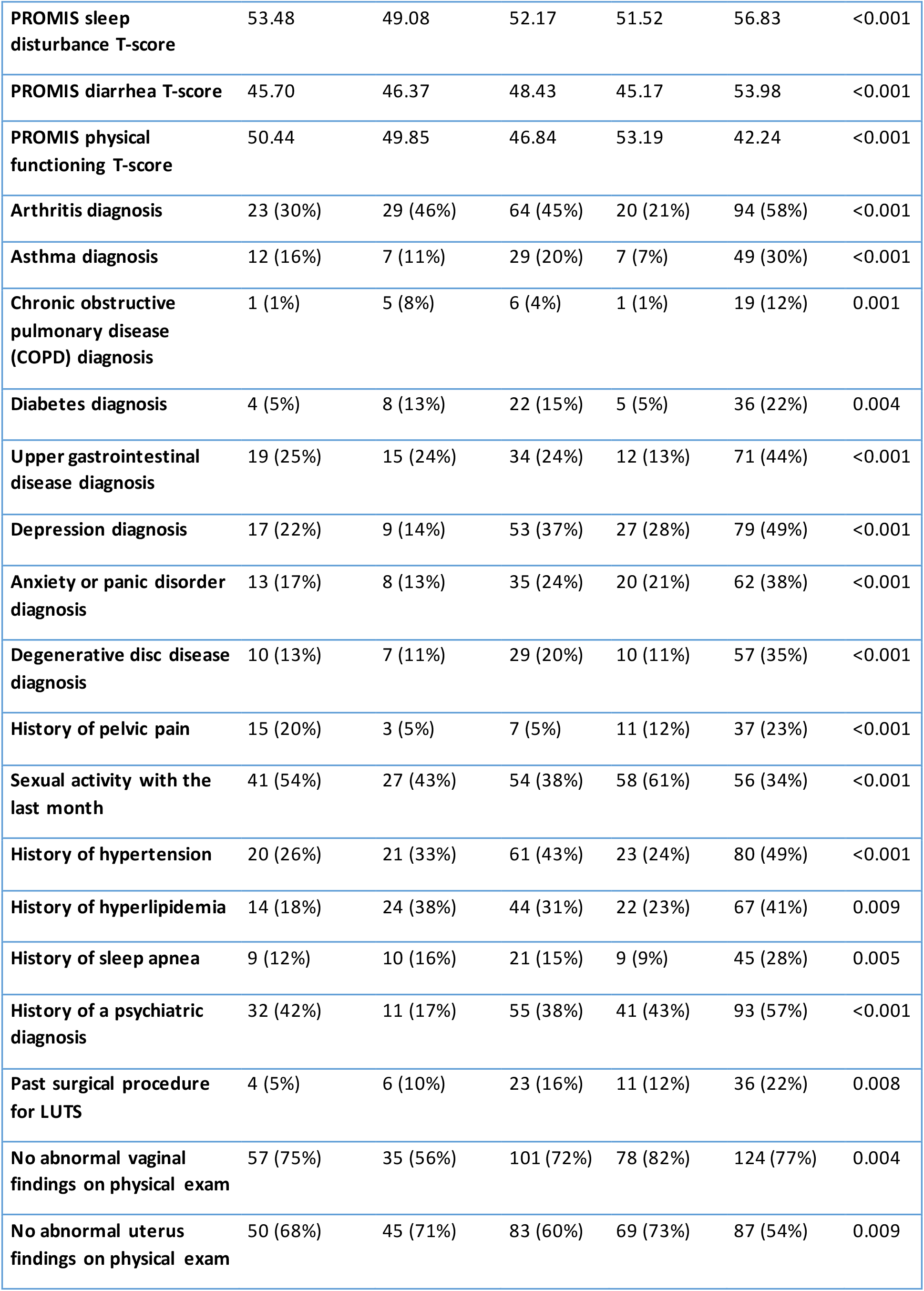
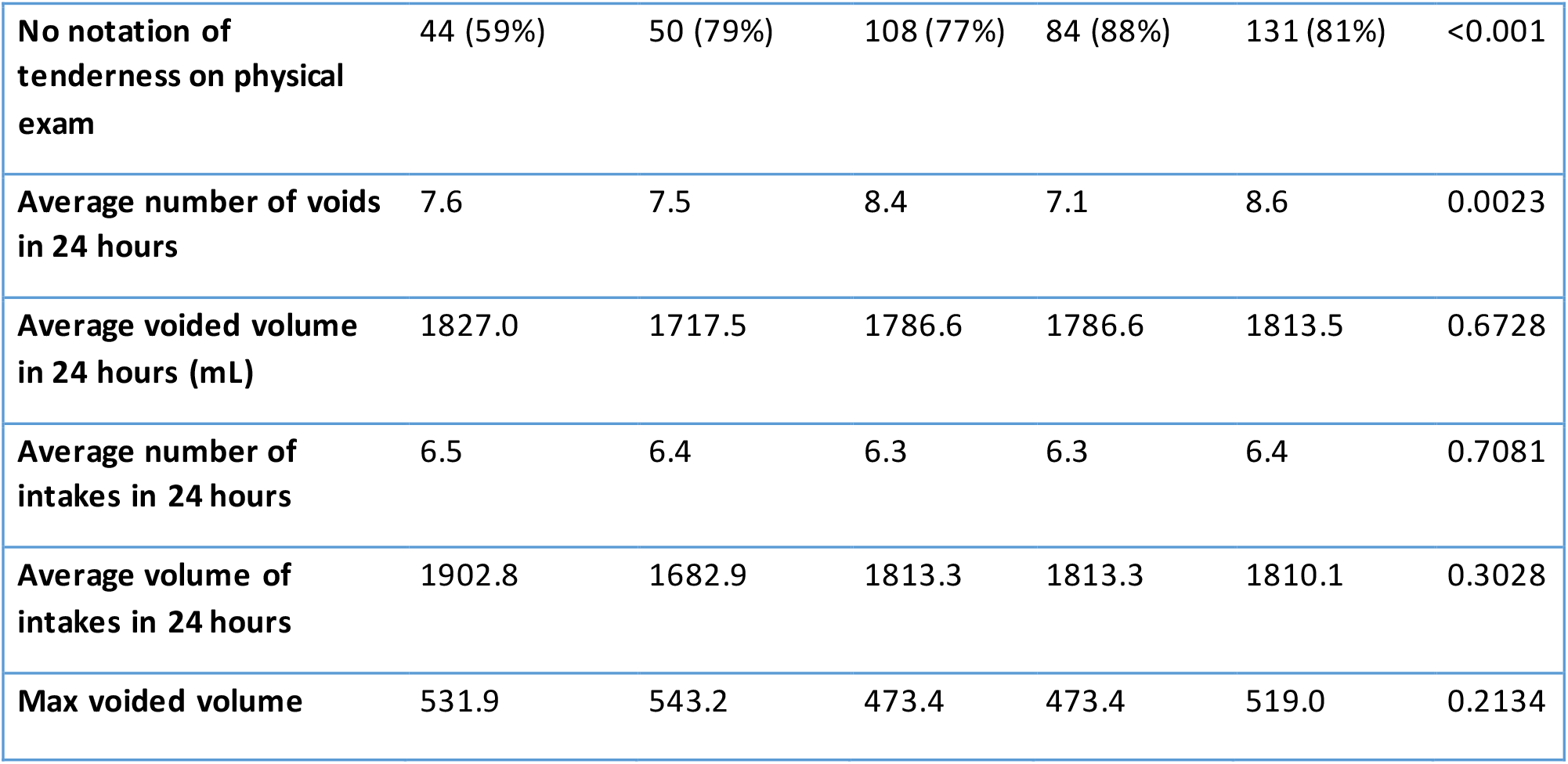
Urinary symptoms, bladder diary variables, non-urinary symptoms, and clinical variables across clusters W1-W5.

Properties of the five clusters are visualized in Figure 4. Each column represents one of five clusters. Radar plots in the first row illustrate urinary symptoms measured by LUTS Tool and AUA-SI; the second row illustrates demographics, clinical measurements, and non-urinary PROs; the third row shows categorical data on comorbidities and anomalies identified during the physical exam; the fourth row shows intake and voiding pattern variables collected in bladder diaries. Radar plots represent mean values of the raw variables across members of each of the clusters. None of the clusters could be characterized by a single symptom, but rather by a combination of symptoms with various levels of severity. Women in all five clusters reported higher than normal frequency of voiding (with the highest frequency in W3 and W5). Women in all clusters except W1 reported urinary urgency and some level of incontinence.

**Figure 4.**
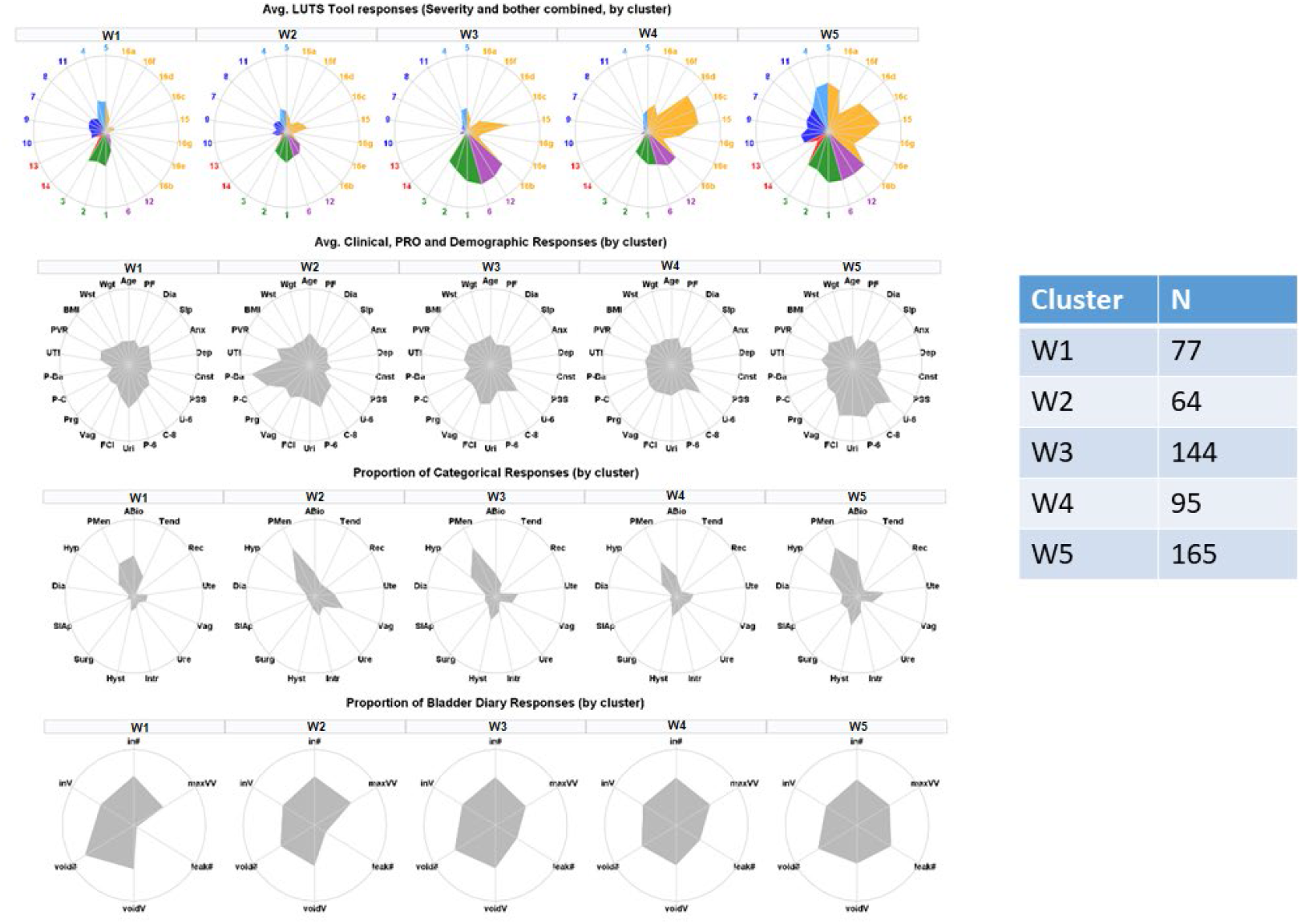
Radar plots illustrating mean values of urinary symptoms, demographics, clinical measurements, non-urinary PROs, comorbidities, and bladder diary variables for identified clusters of women with LUTS. Urinary symptoms are color-coded: green=frequency; blue=post-micturition; purple=urgency; dark blue=voiding; red=pain; orange=incontinence.

Women in cluster W1 (n=77) reported minimal urinary incontinence, but had mostly voiding and post-micturition symptoms (post-void dribbling, trickling, straining, hesitancy, and incomplete bladder emptying). They were younger than the average across the LURN female cohort, had a lower than average weight, number of pregnancies, and vaginal births. They had less comorbidities and abnormal findings in the physical exam. Women in cluster W2 (n=64) reported mild urinary symptoms, including mild urinary incontinence. They presented clinically significant anterior vaginal wall prolapse (mean POP-Q point B anterior [Ba]=1.24 cm, which is outside the introitus), apical prolapse (mean POP-Q point C=-2.38 cm), and the most severe pelvic organ prolapse symptoms (with the highest Pelvic Organ Prolapse Distress Inventory [POPDI-6] values of 20.60). They were on average older(66 vs. 53 years old), and had a higher number of pregnancies (2.9 vs. 1.8) and vaginal births (1.47 vs. 0.92) than women in cluster W1. They also had the highest post-void residual urine volume (75 mL) across the clusters. Women in cluster W3 (n=144) reported high urinary frequency, urinary urgency, and urgency urinary incontinence. They had increased weight, had larger waist circumference, and higher functional comorbidity index (FCI) than women in W1, W2, and W4. They most frequently reported “urgency with fear of leakage” (2.85), but did not report any substantial post-voiding symptoms. Women in cluster W4 (n=95) reported multiple symptoms associated with stress urinary incontinence, as well as urgency urinary incontinence, and some post-void urinary incontinence. They were younger (mean 51 years), had less medical comorbidities (FCI =1.24), and had a higher level of physical functioning (PROMIS T-score=53.2) than others in the cohort. Women in W5 (n=165) reported higher frequencies and severities of LUTS for all symptoms. For 27 out of 30 urinary symptoms, they reported the highest levels across all five clusters. These women were heavier (87.6 Kg), had the lowest level of physical functioning (PROMIS T-score=42.2), had more medical comorbidities (FCI=3.64), and more pregnancies (3.07) than the rest of the cohort. They also reported higher psychosocial difficulties in depression, anxiety, and perceived stress, as well as sleep disturbance.

The presence of multiple significantly different variables across the clusters demonstrates that clusters W1-W5 meet the concise definition of clustering given by Liao as: “The goal of clustering is to identify structure in an unlabeled data set by objectively organizing data into homogeneous groups where the within-group-object dissimilarity is minimized and the between-group-object dissimilarity is maximized” [8]. Figure 5 visually illustrates results of pairwise comparisons of the clusters in a matrix form. Figure 5A provides cluster comparisons by LUTS Tool variables. Elements on the diagonal of the matrix present the level of severity for each LUTS Tool question, i.e., the severity urinary symptom signature of the cluster. The triangle of boxes above the diagonal demonstrates variables significantly different in the pairwise comparison of the clusters; each colored bar indicates a significantly different variable. As seen, the majority of symptoms are significantly different in the pairwise comparison of the clusters. Elements in the lower triangle of the matrix present the difference in symptom severity levels; e.g., the first (upper) element in the triangle represents the difference between symptom severity levels in cluster W2 and cluster W1, indicating that urgency symptoms are more severe in cluster W2, while voiding and pain symptoms are more severe in cluster W1 than in cluster W2. Similarly, Figures 5B-5C provide the results of pairwise comparison of the clusters for other variables from Tables 2-3, demonstrating multiple significantly different non-urologic symptoms, physical examination, clinical and demographic variables. In summary, clusters are distinct and significantly different, not only by their urinary symptom signatures, but by multiple non-urologic variables as well. Importantly, these significant differences are demonstrated by omnibus test (Tables 2-3) and by pairwise comparison of the clusters (Figure 5).

**Figure 5.**
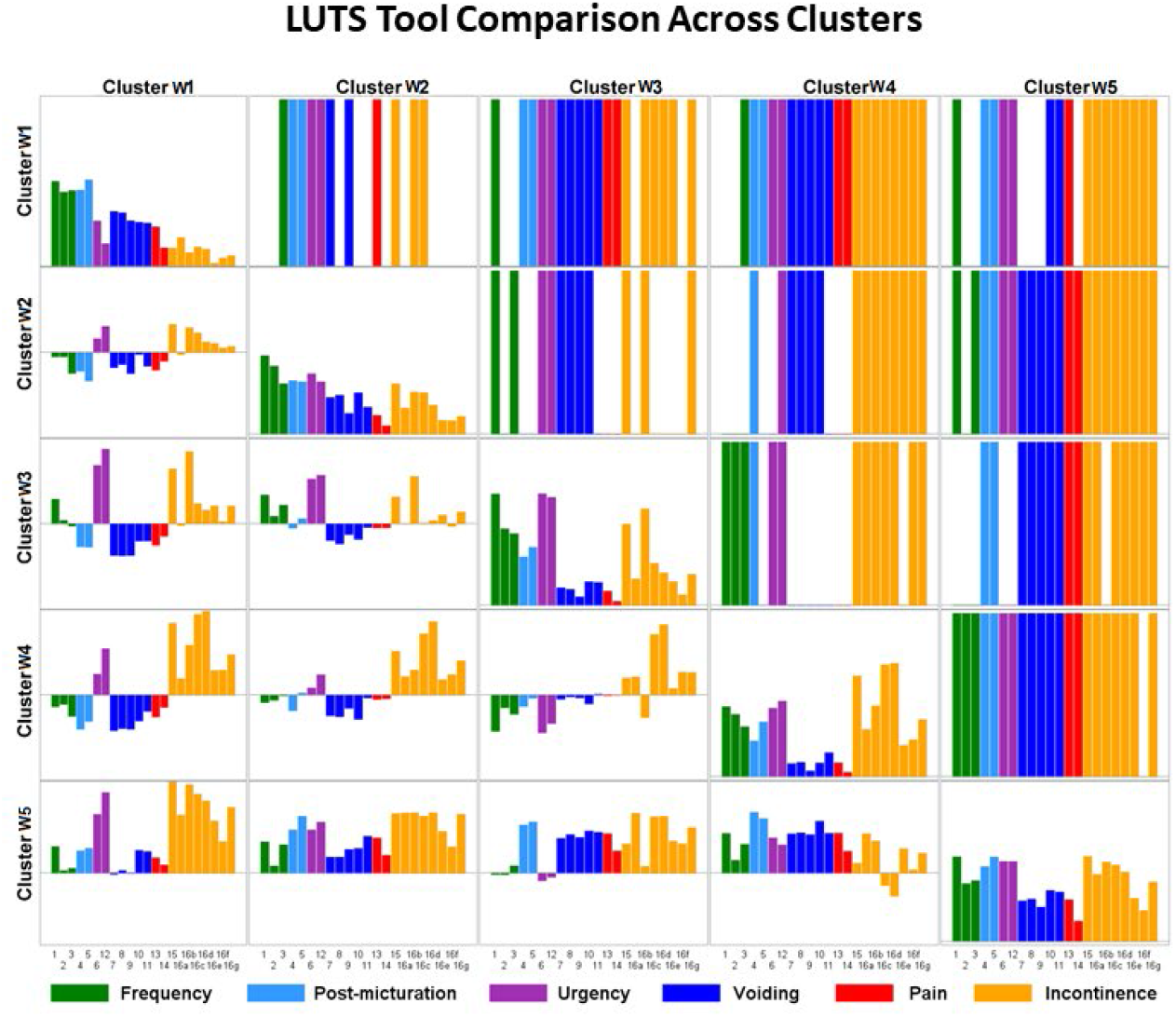
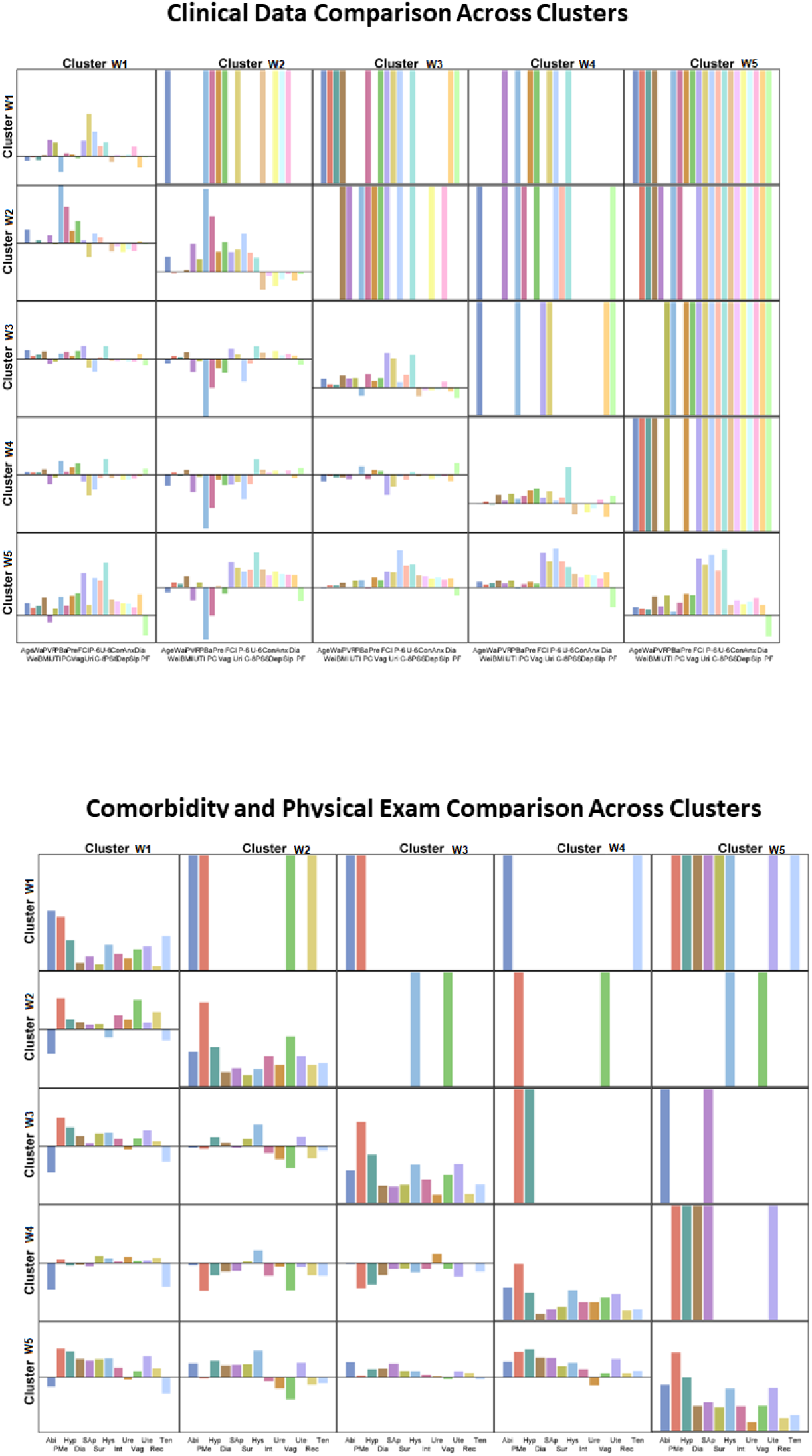
Results of the pairwise comparison of clusters W1-W5. (A) LUTS Tool variables. (B) Demographic and clinical variables. (C) Physical examination and comorbidities data.

### Differential protein abundance in serum of women with LUTS versus non-LUTS controls

Figure 6 presents the volcano plots comparing abundances of 276 proteins in baseline serum samples of women with LUTS versus non-LUTS controls. Figure 6A compares the abundances for all 230 women with LUTS to 30 controls, while Figures 6B-6F provide similar comparisons for members of the identified clusters W1-W5 for whom proteomics data was available (n1=37, n2=38, n3=53, n4=42, n5=60). Supplemental Table S3 provides the lists of significantly differentially abundant proteins for each of the comparisons. Multiple differentially abundant proteins are observed in the serum samples of women with LUTS versus non-LUTS controls, both overall and between each cluster and controls. While some of these have been shown [72] to be associated with LUTS (e.g., tumor necrosis factor [TNF], interleukin-10 [IL-10], monocyte chemotactic protein [MCP], and transforming growth factor [TGF]), the remainders are novel. The highest number of the differentially abundant proteins of 70 (29 after FDR correction for multiple testing) is observed for cluster W5, which demonstrated the highest level of all urinary symptoms and comorbidities. Note that overlap between the lists of differentially abundant proteins is quite low, meaning that clusters W1-W5 are “biochemically” different. The highest overlap of 18 differentially abundant proteins is observed for cluster W5 and cluster W3, defined mainly by high urinary frequency, urinary urgency, and urge urinary incontinence. Interestingly, the lowest number of differentially abundant proteins (n=10) are observed in clusters W2 (characterized by the presence of pelvic organ prolapse) and W4 (characterized by the presence of stress urinary incontinence), which are presumably driven by anatomic abnormalities, rather than biochemical changes. Without going into the detailed interpretation of these results, which are outside the scope of this paper, we think the observed differences in the differentially abundant proteins across W1-W5 clusters serve as important independent confirmation of the distinctiveness of the identified clusters.

**Figure 6.**
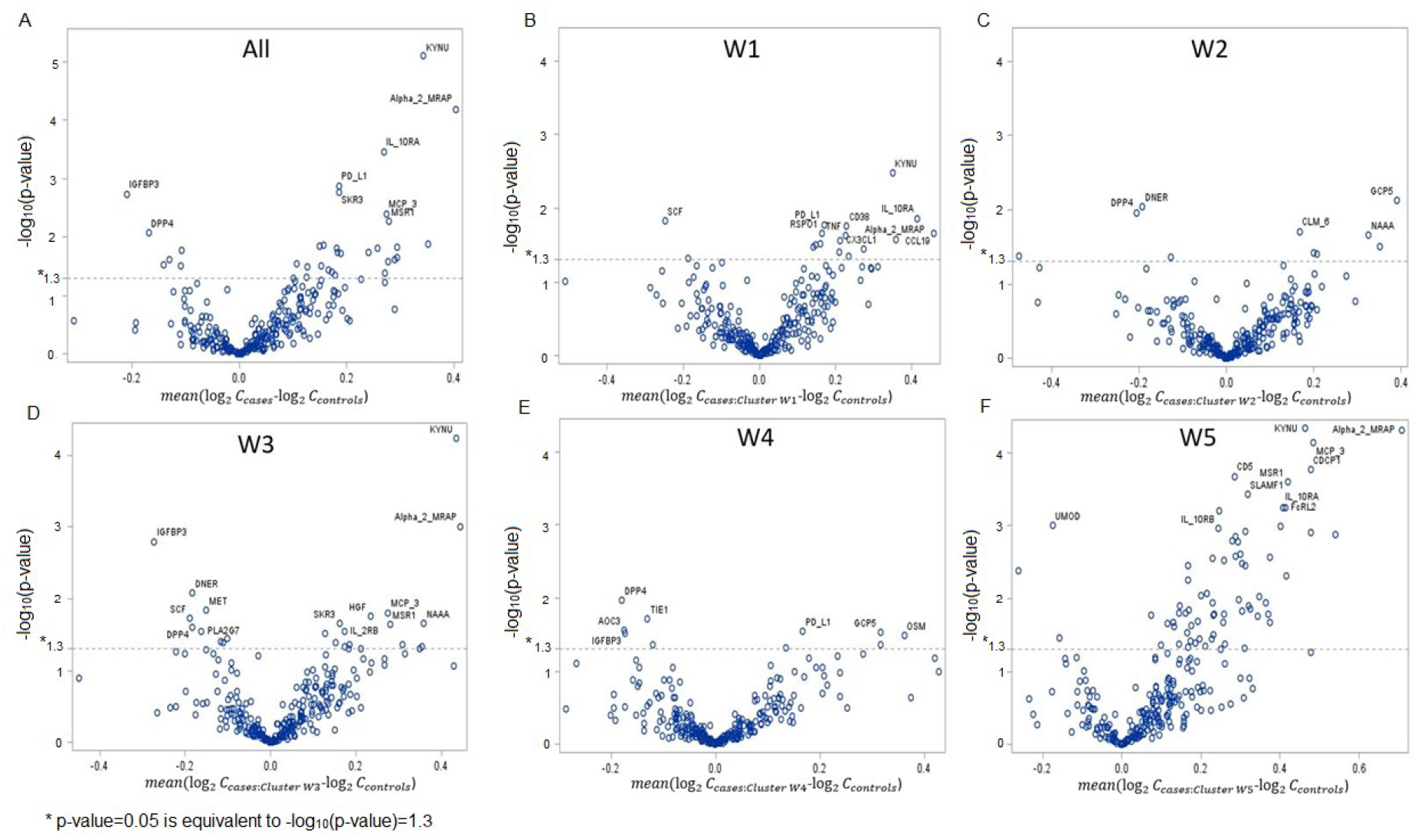
Volcano plots demonstrating differentially abundant proteins in women with LUTS vs. controls for 230 participants representing: (A) the whole cohort; and (B-F) for each of identified clusters W1-W5.

### Comparison of clusters W1-W5 with our previously published urinary symptom-based clusters F1-F4

#### Comparing quality of the clusters

Previously, we identified four clusters (F1-F4) by analyzing data on the same 545 women with LUTS using only urinary symptoms data collected via the LUTS Tool and AUA-SI (total of 52 variables) [41]. Since the same resampling procedure was performed when generating W1-W5 and F1-F4, both cluster structures are equally robust to the random variations of the cohort composition.

Distinctiveness, as determined by pairwise comparisons, was higher for the refined clusters compared with the previously published clusters (Tables 4 and 5). The proportion of significantly different variables in pairwise comparison of the clusters ranged from 27% to 83% (mean 52%) for the refined clusters, compared with a range of 27% to 58% (mean 43%) for the previous clusters. The proportion of core clusters was also higher for W1-W5, compared with F1-F4; it ranged from 73% to 93% (mean 87%) for the refined clusters, compared with a range of 40% to 83% (mean 62%) for the previous clusters. Summarizing, there are more significantly different variables across refined clusters (W1-W5) than across our previously published clusters (F1-F4), and the refined clusters contain a higher percentage of core members for whom the probability to be in the given cluster is higher than the probability to be in all other clusters combined. Therefore, the refined clusters identified in the current paper by using additional urinary and non-urinary variables (total of 185 variables) are substantially more distinct than our previously published clusters based only on urinary symptoms measured by the LUTS Tool and AUA-SI (52 variables).

**Table 4.** Proportion of significantly different variables in clusters W1-W5 and F1-F4.

|  | W1 | W2 | W3 | W4 | W5 |  | F1 | F2 | F3 | F4 |
| --- | --- | --- | --- | --- | --- | --- | --- | --- | --- | --- |
| W1 |  | 27% | 46% | 58% | 64% | F1 |  | 34% | 47% | 58% |
| W2 | 27% |  | 39% | 37% | 83% | F2 | 34% |  | 27% | 51% |
| W3 | 47% | 27% |  | 34% | 66% | F3 | 47% | 27% |  | 39% |
| W4 | 58% | 37% | 34% |  | 69% | F4 | 58% | 51% | 39% |  |
| W5 | 64% | 83% | 66% | 69% |  |  |  |  |  |  |
| Mean |  |  |  |  | 52.3% | Mean |  |  |  | 42.7% |

**Table 5.** Proportion of core clusters in W1-W5 and F1-F4.

|  | PCC |  | PCC |
| --- | --- | --- | --- |
| W1 | 88% | F1 | 40% |
| W2 | 73% | F2 | 74% |
| W3 | 89% | F3 | 51% |
| W4 | 93% | F4 | 83% |
| W5 | 92% |  |  |
| Mean | 87% | Mean | 62% |

#### Comparing cluster membership

The Sankey diagram in Figure 7 serves to compare cluster membership in W1-W5 and in F1-F4. Refined cluster W1 is mostly composed of the members of cluster F1 without prolapse. Cluster W2 is formed by the members of cluster F1 with pelvic organ prolapse and includes some members of other clusters having prolapse and moderate urinary symptoms. Cluster W3 is mainly composed of members of F2 and F3 with urgency urinary incontinence. Cluster W4 is predominantly formed by members of F3 with stress urinary incontinence. Cluster W5 includes nearly all members of F4 and approximately 30% of F3 who have both urgency urinary incontinence and stress urinary incontinence symptoms.

**Figure 7.**
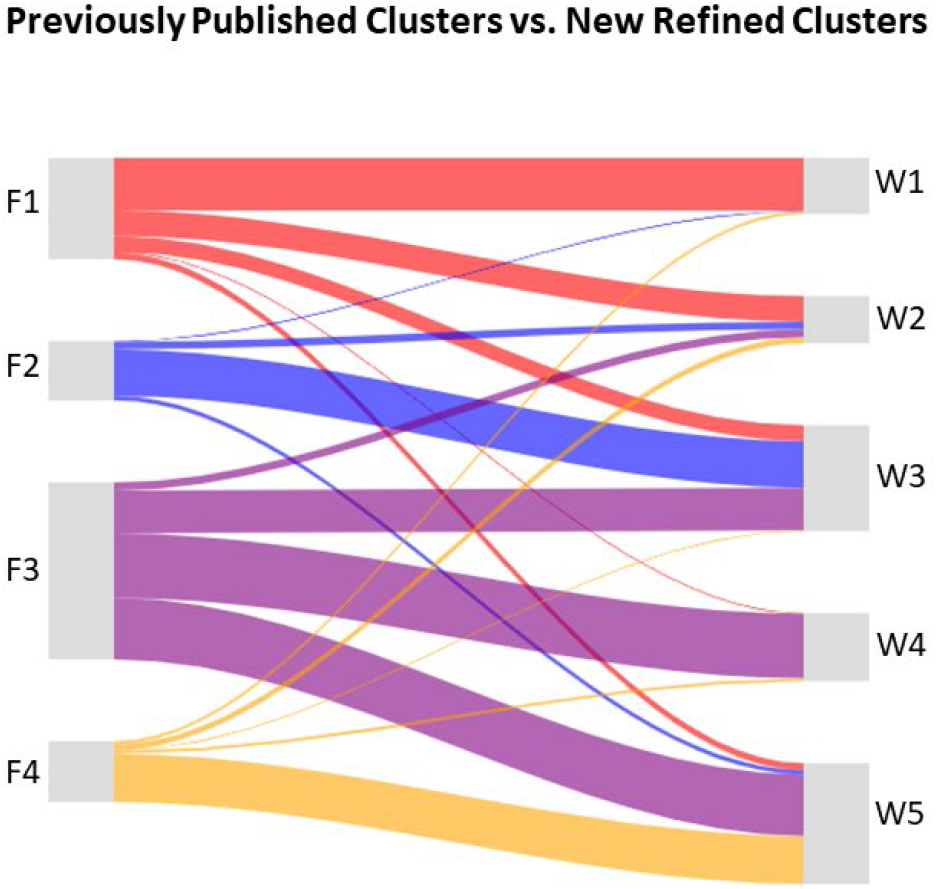
Sankey diagram comparing cluster memberships in W1-W5 and F1-F4.

#### Comparing radar plots

Figure 8 provides comparison of radar plots for the urinary symptom signatures of refined clusters W1-W5 and symptom-based clusters F1-F4. There are substantial similarities in the urinary symptom signatures of our previously published clusters, and of the refined clusters. Radar plots for W5 and F4 are similar in presenting all urinary symptoms at a uniformly high level. Signatures of W4 and F3 are similar in presenting the combination of stress urinary incontinence, urgency, and voiding dysfunction symptoms. W3 and F2 present urinary urgency, urgency urinary incontinence, and mild voiding problems. Symptom signatures of W1 and F1 are similar, presenting mostly voiding and post-micturition problems. The signature of cluster W2 presents mild LUTS and is mostly defined by clinically significant pelvic organ prolapse. The observed similarity of the clusters’ LUTS signatures confirms that additional variables did not result in radical changes, but rather in incremental changes that allowed for identification of the refined clusters, which are built upon, but are more distinct and uniform than, our previously published ones. We believe this is further evidence of the stability of the identified clusters. Urinary symptom data captured by the LUTS Tool and AUA-SI provided the foundation for data-driven subtyping of LUTS, while the remaining urinary and non-urinary variables allowed for identifying refined clusters that differ not only by urinary symptoms, but by other PRO, demographic, and clinical variables as well (Table 3, Figure 5, and Figures 6B-6C). Importantly, cluster refinement enhanced the distinctiveness and uniformity of the clusters, as well as the confidence in cluster membership, by increasing the overall proportion of core clusters from 62% to 87%.

**Figure 8.**
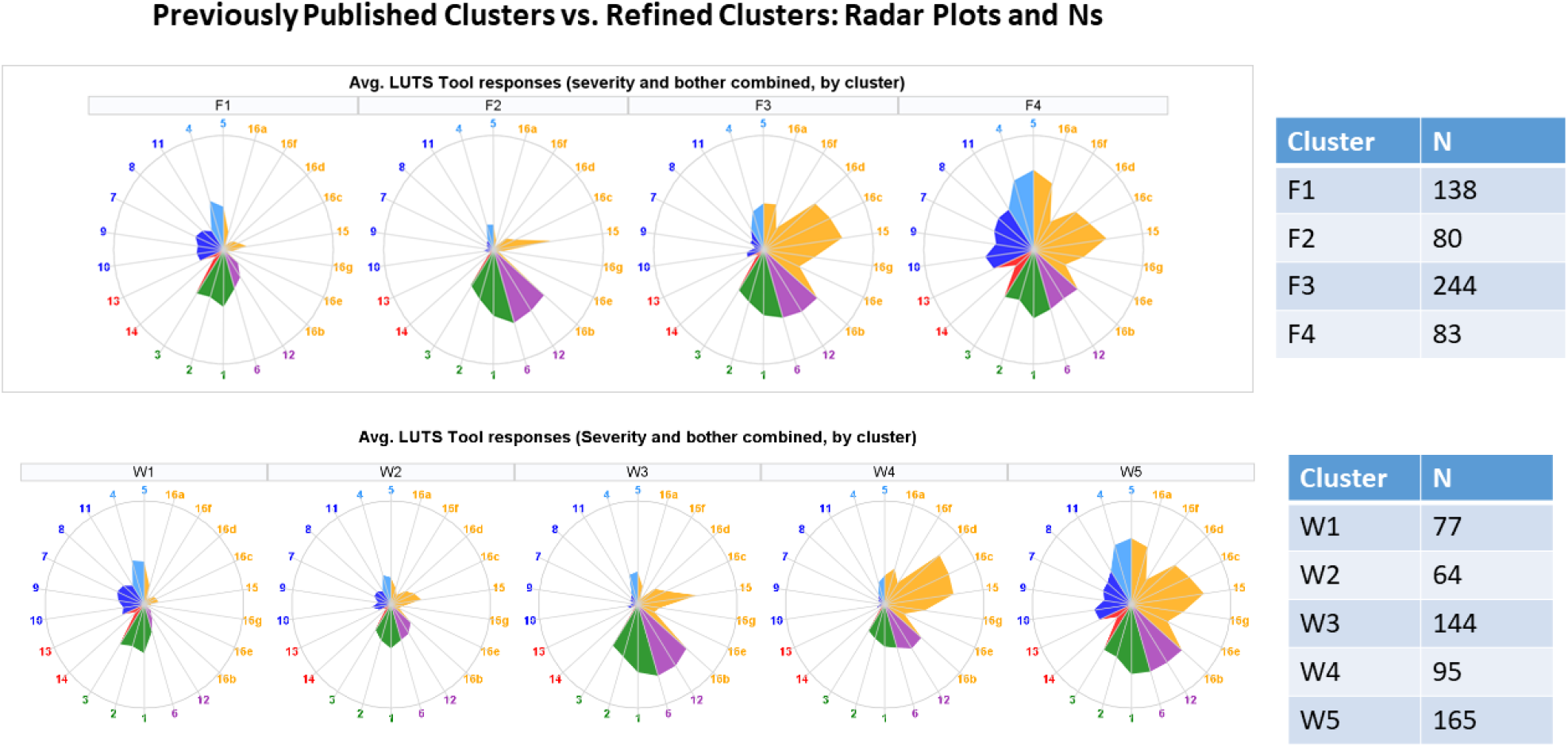
Comparison of radar plots of the urinary symptom signatures for clusters W1-W5 and F1-F4.

### Evolution of refined clusters W1-W5 in 3- and 12-month follow-up

Figure 9 presents radar plots of the urinary symptom signatures of refined clusters W1-W5 at 3- and 12-month follow-up. As seen, the shapes of the radar plots are conserved, while their areas representing overall severity of LUTS are decreased due to improvement in urinary symptoms for some participants. The percentage of improvers varied across the clusters (Table 6). Improvers were defined as having a ½ SD or greater improvement between baseline and 12 months on the calculated LUTS Tool Summary Score (including all 22 LUTS Tool severity variables). We view stability of the urinary symptom radar plot signatures’ shapes as additional evidence of robustness of the identified clusters.

**Table 6.** Percentage of improvers in cluster W1-W5 in 3- and 12-month follow-up.

| Cluster | N patients with 12-month follow-up | N (%) Improvers at 12 months |
| --- | --- | --- |
| Cluster W1 | 50 | 23 (46%) |
| Cluster W2 | 51 | 23 (45%) |
| Cluster W3 | 106 | 57 (53%) |
| Cluster W4 | 70 | 49 (70%) |
| Cluster W5 | 119 | 69 (58%) |

**Figure 9.**
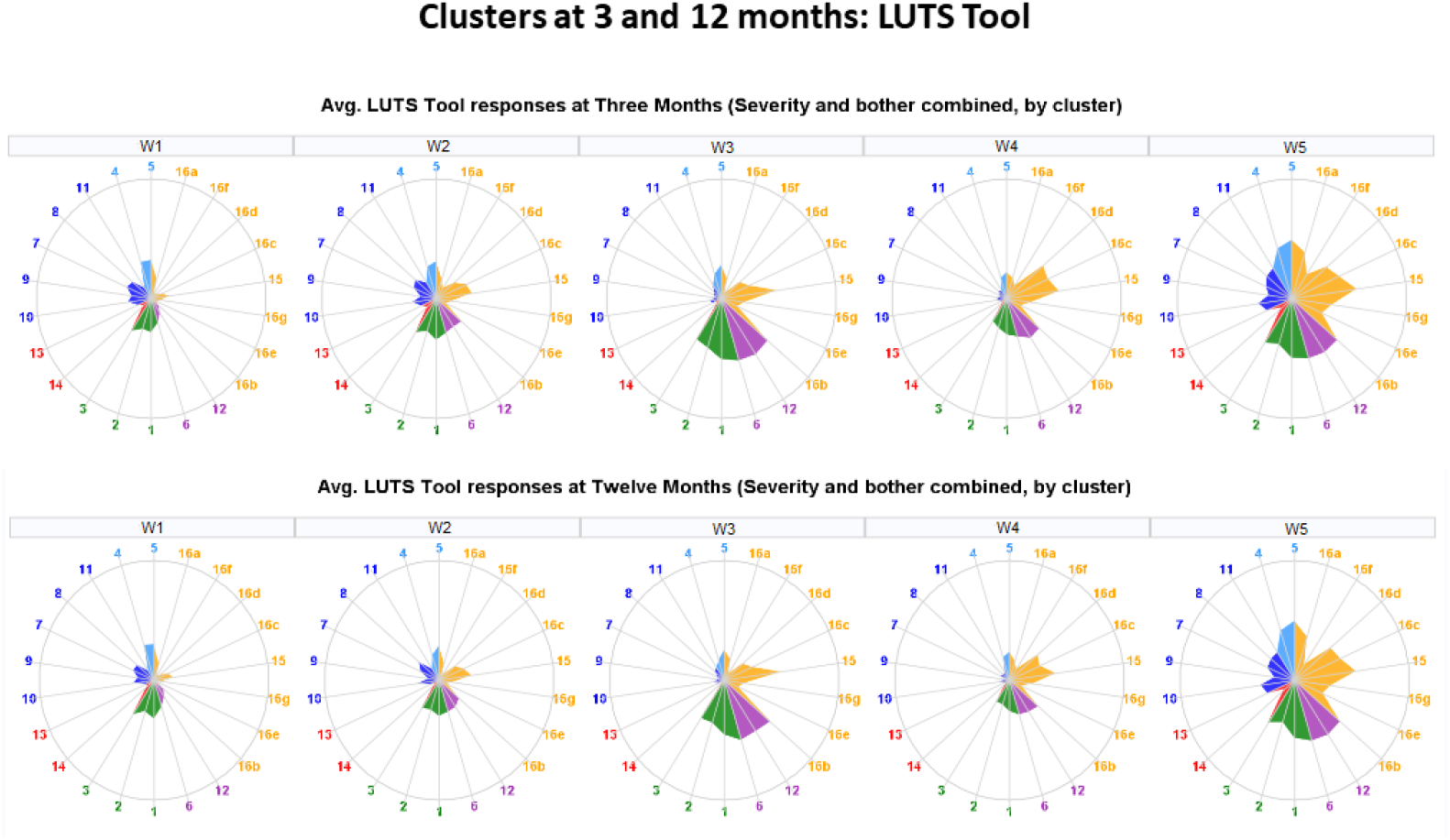
Evolution of the urinary symptom signatures in 3- and 12-month follow-up.

### Clinical significance of the identified clusters

The current paradigm for managing patients with LUTS is to assign a diagnosis based on a pre-defined symptom complex, such as overactive bladder (OAB), or based on a single predominant symptom, such as nocturia or stress urinary incontinence. Treatments are then administered based on these diagnoses [73-74]. Conventional classification of LUTS includes such partially overlapping groups as OAB wet, OAB dry, continent, stress urinary incontinence, urgency urinary incontinence, mixed urinary incontinence, underactive bladder, and bladder outlet obstruction. As stated in [39-41], there are limitations to this paradigm, as patients frequently present with multiple other urinary symptoms in addition to those being treated, and these combinations of symptoms may be relevant to treatment selection. Diagnosis and treatment based solely on patients’ chief complaints may be unsatisfactory, as they disregard other presenting symptoms. Mechanistic studies reveal that a functional impairment to a specific organ in the urinary tract may cause more than a single symptom. For example, a weak urethral sphincter is associated with both stress and urgency urinary incontinence [58, 75]. This may explain why mixed incontinence is so common. This raises the question of how current diagnostic paradigms correspond with biological changes of the continence system and how symptoms occur in women seeking treatment, which was the main rationale for the unbiased data-driven subtyping of women with LUTS in LURN. As seen, none of the clusters W1-W5 identified in our analysis could be characterized by a single symptom, but rather by a combination of symptoms with various levels of severity, which are in concert with the clinical observations mentioned above [58, 75]. The more detailed comparison of our refined clusters W1-W5 with the conventional LUTS groups and subtypes of LUTS identified by other researchers [36-38,76] is provided in Supplemental Material text.

### Clustering methodologies reported in studies on subtyping LUTS and other common diseases and disorders

Previous research on subtyping of diseases and disorders provides various levels of detail regarding the clustering methodologies that were employed. For instance, Coyne et al [36] provided detailed information on the 14 LUTS questions used for clustering and indicated that all the variables were scaled from 0 to 1. They took care of the robustness of clusters to the variation in composition of the cohort by performing clustering on the random 50% subset of participants first, and then by extending it to the whole cohort. They used k-means algorithm for clustering and scanned the number of clusters K from 3 to 7. They reported that the decision on the number of clusters was made by evaluating “each cluster model based on the clinical relevance and distinctiveness of each cluster, as well as the amount of variance accounted for by the cluster solution.” However, they did not provide the names or values of criteria used for cluster evaluation. They provided detailed and informative descriptive statistics, but for some reason, did not provide any tests on significantly different variables in pairwise comparison of the identified clusters.

Similarly, Hall et al [37] provided detailed information on the 14 LUTS questions and scaling. K-means clustering was performed with random 50% split of the cohort (split-half validation) similar to [36]. Detailed descriptive statistics and omnibus tests are provided across identified clusters and asymptomatic controls for significant difference in the variables not used for clustering, including demographics, comorbidities, risk factors, and lifestyle factors. However, no information on significant differences in the 14 LUTS variables used for clustering, and no information on pairwise comparison of the clusters is provided. Summarizing, these two LUTS clustering papers provide a reasonable level of details on scaling of the variables and on the clustering procedure, but unfortunately do not provide enough information on evaluation of the quality of the identified clusters.

In contrast, Miller et al [38] provided all necessary information on pairwise comparison of the identified clusters, both for six variables used for clustering, and for eight other variables collected but not used for clustering. Unfortunately, the authors did not perform any scaling of variables used for clustering. As stated in the paper, “the six clustering variables were (a) number of voids during daytime hours, (b) number of voids during nighttime hours, (c) daytime modal output volume in milliliters, (d) total 24-hour output volume in milliliters, (e) total 24-hour beverage intake in milliliters, and (f) BMI (a variable that was speculated to be related to intake).” If the volume of related variables *c, d, e* were not scaled but entered into clustering in milliliters (values can be as high as 500), then these variables will provide the domineering contribution (compared with the number of daytime voids, typically < 20) to the 6-dimensional Euclidean distance between clustered objects, and will serve as drivers determining cluster membership. The contribution of these variables to the Euclidean distance would be much lower if they were entered in liters instead of milliliters, which would change the cluster membership. This is the problem indicated by Hair et al [62], with unscaled, unstandardized data of the inconsistency between cluster solutions when the scale of some variables is changed, which is a strong argument in favor of standardization. To our mind, the best solution of the problem is the use of the variables scaled by comparison with controls, as we described in the Methods section. Proper scaling is especially important when using heterogeneous data combining dimensionless and dimensional variables measured in different units, as in [38], where frequencies, volumes (mL), and BMI units (kg/m^2^) are combined.

A broader look outside the LUTS domain shows that previous data on subtyping other common complex diseases provide different level of details on the clustering procedure and cluster evaluation as well. Table 7 below summarizes methodological information reported in the clustering papers [31-33] subtyping patients with asthma, diabetes, and sepsis, as well as in the LUTS papers [36-38] discussed above. More details on methodological information reported in [31-33] are provided in Supplemental Material text.

**Table 7.**
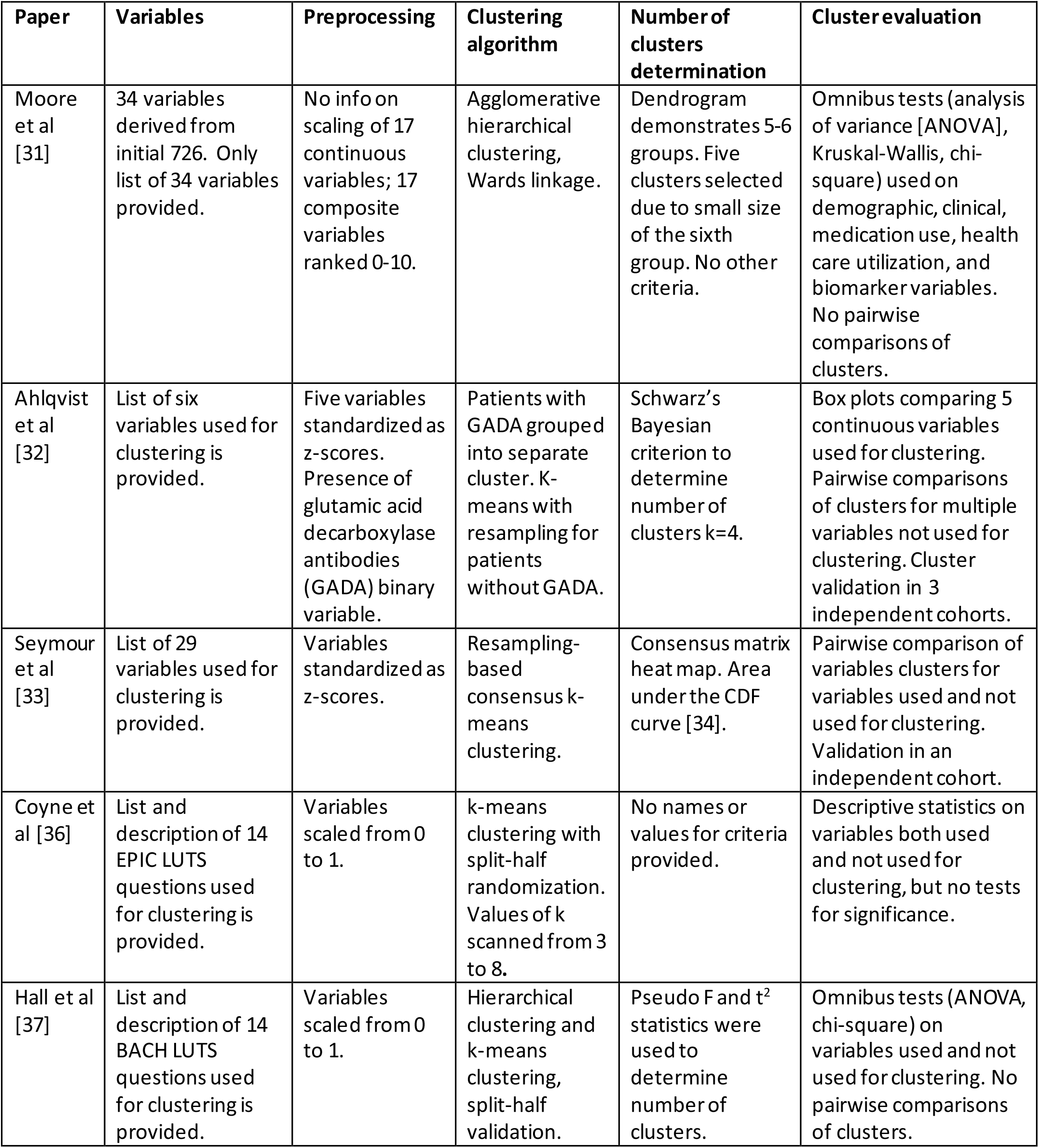
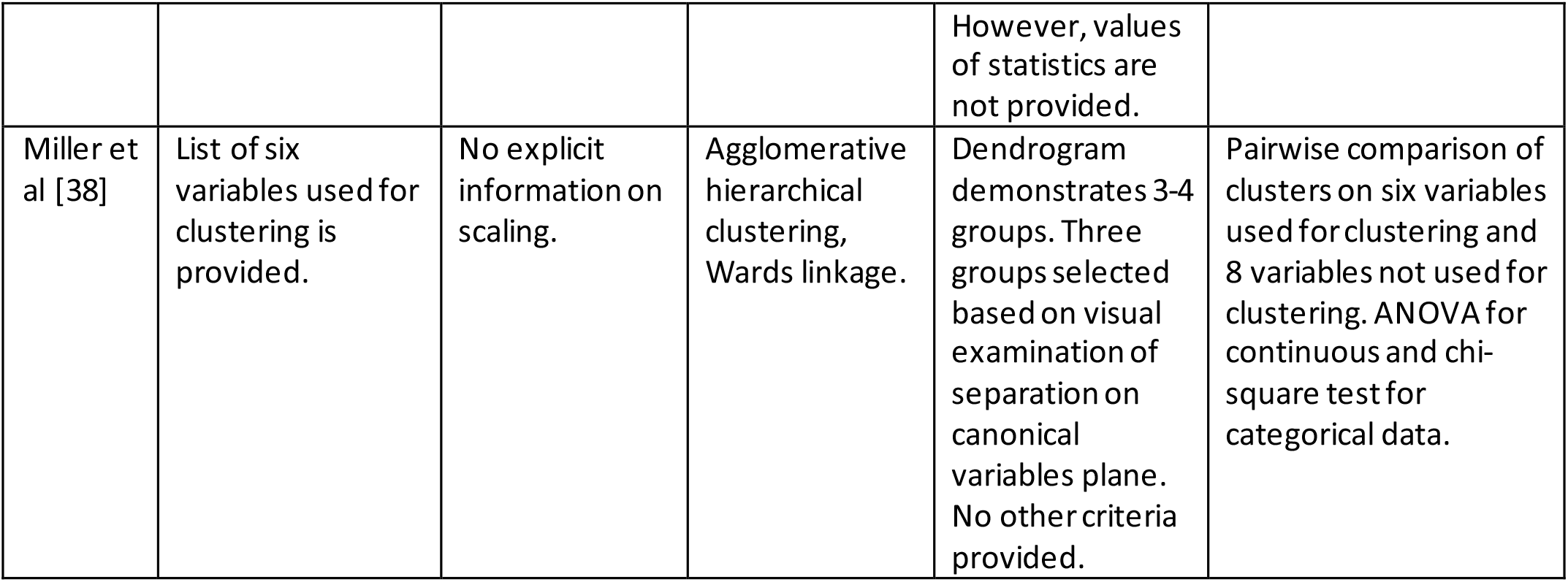
Methodological information provided in the clustering papers.

### Thoughts on minimal requirements for clustering publications

As shown above, publications using clustering for disease subtyping provide different levels of details on data preprocessing, clustering procedure, and cluster evaluation. In particular, information on scaling and weighting of the variables, values of criteria used for selection of the number of clusters, pairwise comparison of the clusters, and level of confidence in cluster membership are often not provided. This missing or hard-to-find information is important for better understanding of the papers’ results, for comparison of the proposed phenotypes with previous and future classifications, and for potential refinement. With this in mind, and guided by the principles of FAIR data (findability, accessibility, interoperability, and reusability) [77-78], we think it is time for the clustering community to develop minimum requirements for clustering reports (MICRo), similar to minimum information about a proteomics experiment (MIAPE), developed by the proteomics community [79], and minimum information about a microarray experiment (MIAME), developed by the transcriptomics community [80].

Exact guidelines for the minimal requirements for clustering publications should result from the clustering community discussion. Here, we would like to call for such discussion and propose the below items that we believe are important for future guidelines:

1. Complete list of variables used for clustering.
2. Explicit information on scaling and weighting of clustering variables.
3. Clustering algorithm used (name of the function with options and parameter values, or code).
4. Exact definition of criteria used to determine the number of clusters. Values of criteria for the selected and alternative number of clusters. Preferably, more than one criterion should be presented.
5. Results of pairwise comparison of the clusters, with the indication of clinically meaningful and significantly different variables in the pairwise comparison – for all variables used for clustering, and for selected important variables not used for clustering (e.g., demographics).
6. Information on the level of confidence in cluster membership.

### Limitations of the current study

Our paper carries some limitations. First, there are limitations in terms of the cohort, which included only treatment-seeking and predominantly white participants. Our analysis only contains women. Preliminary data analysis confirmed that sex is the major determinant of LUTS subtypes; therefore, sex-specific clustering was performed. We previously published the results of urinary symptom-based clustering of male participants [64]. Cluster refinement of male clusters, along the same lines as described in the current paper, is underway; the resulting refined subtypes will be compared with those found in the female cohort. Our control group used for scaling of the variables was relatively small (55 participants) but commensurate with the size of identified clusters. Not all data elements collected for the cases were available for the controls, so we used some literature data for general population and bladder diary data from a different study (EPI).

Second, there are limitations in terms of data elements used for clustering. Genomics data were not used so far. We do not expect that genomics data will produce dramatic effect on clustering results since LUTS is a highly prevalent common disease, especially in older age. Nevertheless, genotyping of the LURN participants is underway, which would allow for future cluster refinement by including the binary single nucleotide polymorphism (SNP) data using our novel weighted Tanimoto indices approach. Proteomics data are available for approximately 40% of the cohort and were not yet used for cluster refinement. However, they were used to demonstrate the presence of multiple significantly different proteins, indicating the refined symptom-based clusters are biochemically different.

Third, we developed methodology and a pipeline for integrating heterogeneous continuous and categorical data for clustering women with LUTS. We cannot claim that this is the preferable methodology for other data sets since data and research questions are different in different studies. However, we explicitly described our preprocessing and clustering procedures, as well as criteria used for determination of the number of clusters, cluster evaluation, and confidence level in cluster membership. We compared our methodology with alternative approaches and demonstrated that our methodology allows for combining heterogeneous continuous, categorical, and binary data, and that our refined clusters are more distinct than the previous urinary symptom-based clusters. Detailed description of the methodology, and comparison with the alternative approaches, allows interested readers to decide if it is suitable for their data and research questions. Availability of the pipeline source code allows for modifications, if needed.

Fourth, and most importantly, clinical significance of the identified clusters has yet to be determined. We already demonstrated the distinctness of our clusters, but now we need to establish their usefulness in clinical practice. This should be done through clinical trials, where treatments and outcomes of patients classified into the identified clusters would be compared with the standard treatment without the knowledge of cluster membership. Our preliminary analysis of multiple differentially abundant serum proteins in women with LUTS corroborated that the identified clusters are biochemically different. Further analysis of the affected biochemical pathways, and their longitudinal dynamics as related to symptom trajectories, will follow with the goal to enhance understanding of different etiologies of the identified subtypes and potentially establish more effective subtype-specific treatments. Further analysis will also include determination of the minimal set of variables sufficient for classification of patients with LUTS into identified subtypes in clinical practice.

## CONCLUSION

A novel clustering pipeline for subtyping of common complex diseases and syndromes using heterogeneous continuous and categorical data was developed. The advantages of scaling variables by comparison with the controls without the disease of interest were discussed and illustrated by the simulated example. The novel weighted Tanimoto indices approach to integrate multiple binary variables into the clustering procedure was developed. A cluster refinement procedure using data available only for the subset of participants through semi-supervised clustering was proposed. A novel contrast criterion (CC) for resampling-based consensus clustering was proposed and compared with existing criteria for consensus clustering, i.e., consensus score (SC) and proportion of ambiguously clustered pairs (PAC). A simulated example demonstrated the advantages of CC over SC and PAC.

Information provided in the literature on subtyping common complex diseases and disorders was reviewed and shown to be often incomplete, especially with regard to data preprocessing, clustering procedures, and cluster evaluation. Suggestions for the minimum requirements for clustering publications were formulated, and the community effort to work on creating such requirements following the principles of FAIR data was called for.

Five distinct clusters of women with LUTS were identified by using 185 variables, including demographics, physical exam, LUTS and non-LUTS questionnaires, and bladder diary variables. The quality of the clusters was evaluated using established criteria (Calinski-Harabasz, Davies-Bouldin, Dunn, Point-Biserial, and Silhouette [22-26]), as well as novel contrast criterion (CC) and percentage of core members of the clusters (PCC). Distinctiveness of the clusters was confirmed by multiple significantly different variables in pairwise comparison of the clusters. Refined clusters W1-W5 were compared with our previously published urinary symptom-based clusters F1-F4, and were shown to be more distinct by having a higher percentage of significantly different variables and a higher percentage of the core members of the clusters. Importantly, targeted proteomics data confirmed that our refined clusters based on clinical data are biochemically different. Identification of the clinically and biochemically distinct subtypes of LUTS has provided a foundation for studies of subtype-specific etiologies and treatments.

## Data Availability

The data that support the findings of this study are openly available in the NIDDK Central Repository at https://repository.niddk.nih.gov/; please reference the acronym LURN.

## ACKNOWLEDGMENTS

Heather Van Doren, Senior Medical Editor with Arbor Research Collaborative for Health, provided editorial assistance on this manuscript.

This is publication number 28 of the Symptoms of Lower Urinary Tract Dysfunction Research Network (LURN).

This study is supported by the National Institute of Diabetes & Digestive & Kidney Diseases through cooperative agreements (grants DK097780, DK097772, DK097779, DK099932, DK100011, DK100017, DK099879) and for Dr. Andreev’s Biomarker Ancillary LURN R01 (grant 5R01DK125251).

Research reported in this publication was supported at Northwestern University, in part, by the National Institutes of Health’s National Center for Advancing Translational Sciences, Grant Number UL1TR001422. The content is solely the responsibility of the authors and does not necessarily represent the official views of the National Institutes of Health.

The following individuals were instrumental in the planning and conduct of this study at each of the participating institutions:

Duke University, Durham, North Carolina (DK097780): PIs: Cindy Amundsen, MD, Eric Jelovsek, MD; Co-Is: Kathryn Flynn, PhD, Todd Harshbarger, PhD, Jim Hokanson, PhD, Aaron Lentz, MD, Michelle O’Shea, MD, David Page, PhD, Nazema Siddiqui, MD, Kevin Weinfurt, PhD Lisa Wruck, PhD; Study Coordinators: Yasmeen Bruton, Paige Green, Folayan Morehead

University of Iowa, Iowa City, IA (DK097772): PIs: Catherine S Bradley, MD, Karl Kreder, MD, MBA, MSCE; Co-Is: Bradley A. Erickson, MD, MS, Daniel Fick, MD, Vince Magnotta, PhD, Philip Polgreen, MD, MPH; Study Coordinators: Mary Eno, Sarah Heady, Chelsea Poesch

Northwestern University, Chicago, IL (DK097779): PIs: James W Griffith, PhD, Kimberly Kenton, MD, MS, Brian Helfand, MD, PhD; Co-Is: Carol Bretschneider, MD, David Cella, PhD, Sarah Collins, MD, Julia Geynisman-Tan, MD, Alex Glaser, MD, Christina Lewicky-Gaupp, MD, Margaret Mueller, MD; Study Coordinators: Sylwia Clarke, Melissa Marquez, Pooja Sharma, Michelle Taddeo, Pooja Talaty. Dr. Helfand and Ms. Talaty are at NorthShore University HealthSystem.

University of Michigan Health System, Ann Arbor, MI (DK099932): PI: J Quentin Clemens, MD, FACS, MSCI; Co-Is: John DeLancey, MD, Dee Fenner, MD, Rick Harris, MD, Steve Harte, PhD, Anne P. Cameron, MD, Aruna Sarma, PhD, Giulia Lane, MD; Study Coordinators: Ashly Chimner, Linda Drnek, Emma Keer, Marissa Moore, Greg Mowatt, Sarah Richardson

University of Washington, Seattle Washington (DK100011): PI: Claire Yang, MD; Co-I: Anna Kirby, MD; Study Coordinators: Lois Meryman, Brenda Vicars, RN

Washington University in St. Louis, St. Louis Missouri (DK100017): PI: H. Henry Lai, MD; Co-Is: Gerald L. Andriole, MD, Joshua Shimony, MD, PhD; Study Coordinators: Linda Black, Vivien Gardner, Patricia Hayden, Diana Wolff, Aleksandra Klim, RN, MHS, CCRC

Arbor Research Collaborative for Health, Data Coordinating Center (DK099879): PI: Robert Merion, MD, FACS; Co-Is: Victor Andreev, PhD, DSc, Brenda Gillespie, PhD, Abigail Smith, PhD; Project Manager: Melissa Fava, MPA, PMP; Clinical Monitor: Melissa Sexton, BA, CCRP; Research Analysts: Margaret Helmuth, MA, Jon Wiseman, MS, Jane Liu, MPH; Project Associate: Levi Hurley

National Institute of Diabetes and Digestive and Kidney Diseases, Division of Kidney, Urology, and Hematology, Bethesda, MD: Project Scientist: Ziya Kirkali MD; Project Officer: Christopher Mullins PhD; Project Advisor: Julie Barthold, MD.

## SUPPORTING INFORMATION

See separate Supplemental Materials document.

## SUPPLEMENTAL MATERIAL TEXT (Section Headings)

Simple example illustrating the benefits of scaling by controls.

Contrast criterion (CC): special cases.

Contrast criterion and proportion of ambiguous clustering: simulation example.

Details on the targeted proteomics study of serum samples of women with LUTS versus controls.

Comparison of the identified clusters W1-W5 with the conventional classification, subtypes, and clusters of women with LUTS identified in literature.

Details on the methodological information provided in the clustering papers [31-33].

## SUPPLEMENTAL TABLES (Captions)

**Supplemental Table S1**. Overview of the variables used for clustering of 545 women with lower urinary tract symptoms (LUTS) in comparison with the same variables for non-LUTS controls.

**Supplemental Table S2**. Means, standard deviations, pairwise distances, and misclassification errors resulting from the use of three scaling approaches (unscaled – U, scaled as cohort’s z-scores – C, and scaled by healthy controls – N).

**Supplemental Table S3**. Lists of differentially abundant proteins observed in the serum samples of 230 women with LUTS versus controls. Proteins with false discovery rate (FDR) adjusted p-value<0.05 are bolded.

**Supplemental Table S3A**. All 230 women with LUTS.

**Supplemental Table S3B**. W1 cluster (n=37).

**Supplemental Table S3C**. W2 cluster (n=38).

**Supplemental Table S3D**. W3 cluster (n=53).

**Supplemental Table S3E**. W4 cluster (n=42).

**Supplemental Table S3F**. W5 cluster (n=60).

## SUPPLEMENTAL FIGURES (Captions)

**Supplemental Figure S1**. Simulated example 1, illustrating distributions of two variables describing patients in two disease subtypes and healthy controls. Upper row – X_1_, lower row X_2_. Left column unscaled, center z-scored, right scaled by controls.

**Supplemental Figure S2**. Simulated example A. Non-overlapping transcript signatures. 40,000 transcripts. Five groups of people. Means, correlation matrix, and single instance of simulated distributions of transcript abundances.

**Supplemental Figure S3**. Simulated example A. Consensus matrices, K=2,3…8. CC vs. PAC. Effect size=2.

**Supplemental Figure S4**. Simulated example A. Consensus matrices, K=2,3…8. CC vs. PAC. Effect size=0.6.

**Supplemental Figure S5**. Simulated example A. Misclassification error vs. effect size. Comparison of consensus clustering using contrast criterion with k-means and hierarchical clustering using Calinski-Harabasz criterion. Three values of correlation coefficient for transcript abundances within transcript signatures.

**Supplemental Figure S6**. Simulated example A1. Same conditions as in simulated examples A, except transcript abundances being correlated following ***R***_***ij***_ = ***r***^|***i***−***j***|^.

**Supplemental Figure S7**. Simulated example B. Overlapping transcript signatures. Consensus matrices, K=2,3…8. CC vs. PAC. Effect size=2.

**Supplemental Figure S8**. Simulated example B. Overlapping transcript signatures. Consensus matrices, K=2,3…8. CC vs. PAC. Effect size=0.8.

**Supplemental Figure S9**. Simulated example B. Misclassification error vs. effect size. Comparison of consensus clustering using contrast criterion with k-means and hierarchical clustering using Calinski-Harabasz criterion. Three values of correlation coefficient for transcript abundances within transcript signatures.

**Supplemental Figure S10**. Simulated example B1. Same conditions as in simulated examples B, except transcript abundances being correlated following ***R***_***ij***_ = ***r***^|***i***−***j***|^.

## Notes

### Competing Interest Statement

The authors have declared no competing interest.

### Clinical Trial

NCT02485808

