## Supplemental Materials for "Subtyping of common complex diseases and disorders by integrating heterogeneous data. Identifying clusters among women with lower urinary tract symptoms in the LURN study"

**Supplemental Table S1. Overview of the variables used for clustering of 545 women with lower urinary tract symptoms (LUTS) in comparison with the same variables for non-LUTS controls.**

| | **Cases** | **Controls** |
|---|---|---|
| | Mean (Std) or N (%) | Mean (Std) or N (%), as available |
| LUTS tool | | |
| Urinate too frequently | 2.3 (1.1) | 0 (1) |
| Urinate too frequently: bother | 2.4 (1.1) | 0 (1) |
| Daytime frequency | 1.8 (0.9) | 0 (1) |
| Daytime frequency: bother | 2 (1.3) | 0 (1) |
| Nighttime frequency | 1.7 (1.1) | 0 (1) |
| Nighttime frequency: bother | 2.1 (1.3) | 0 (1) |
| Incomplete emptying | 1.6 (1.2) | 0 (1) |
| Incomplete emptying: bother | 2 (1.2) | 0 (1) |
| Trickle/dribble | 1.9 (1.3) | 0 (1) |
| Trickle/dribble: bother | 1.8 (1.3) | 0 (1) |
| Sudden rush to urinate | 2.1 (1.1) | 0 (1) |
| Sudden rush to urinate: bother | 2.6 (1.1) | 0 (1) |
| Delay start of urination | 0.8 (1) | 0 (1) |
| Delay start of urination: bother | 1.3 (1.1) | 0 (1) |
| Urine start/stop | 0.9 (1.1) | 0 (1) |
| Urine start/stop: bother | 1.3 (1.1) | 0 (1) |
| Strain to urinate | 0.6 (1) | 0 (1) |
| Strain to urinate: bother | 1.6 (1.2) | 0 (1) |
| Weak urine stream | 1 (1.1) | 0 (1) |
| Weak urine stream: bother | 1.2 (1.2) | 0 (1) |
| Splitting or spraying | 1 (1.2) | 0 (1) |
| Splitting or spraying: bother | 1.5 (1.3) | 0 (1) |
| Sudden rush to urinate with fear of leaking | 2 (1.2) | 0 (1) |
| Sudden rush to urinate with fear of leaking: bother | 2.7 (1.1) | 0 (1) |
| Pain or discomfort in bladder | 0.7 (1) | 0 (1) |
| Pain or discomfort in bladder: bother | 2 (1.1) | 0 (1) |
| Burning while urinating | 0.3 (0.7) | 0 (1) |
| Burning while urinating: bother | 1.9 (1.2) | 0 (1) |
| General leakage | 2 (1.2) | 0 (1) |
| General leakage: bother | 2.9 (1.1) | 0 (1) |

| | | |
|---|---|---|
| Dribble | 1.2 (1.2) | 0 (1) |
| Dribble: bother | 2.4 (1.2) | 0 (1) |
| Rushing with leaking | 1.9 (1.3) | 0 (1) |
| Rushing with leaking: bother | 2.9 (1.1) | 0 (1) |
| Leaking while sneezing | 1.7 (1.4) | 0 (1) |
| Leaking while sneezing: bother | 2.7 (1.1) | 0 (1) |
| Leaking with exercise | 1.5 (1.4) | 0 (1) |
| Leaking with exercise: bother | 2.7 (1.1) | 0 (1) |
| Leaking while sleeping | 0.7 (1.1) | 0 (1) |
| Leaking while sleeping: bother | 2.6 (1.2) | 0 (1) |
| Leaking with sex | 0.4 (0.9) | 0 (1) |
| Leaking with sex: bother | 2.7 (1.3) | 0 (1) |
| Leaking for no reason | 1.1 (1.2) | 0 (1) |
| Leaking for no reason: bother | 2.6 (1.2) | 0 (1) |
| AUA-SI | | |
| Times urinate overnight | 2.1 (1.4) | 0 (1) |
| Times bladder not completely empty | 1.7 (1.6) | 0 (1) |
| Times urinate less than 2 hours since last urination | 2.8 (1.4) | 0 (1) |
| Times urine stop/start | 1.2 (1.4) | 0 (1) |
| Times difficult to postpone urination | 2.9 (1.7) | 0 (1) |
| Times weak stream | 1.2 (1.4) | 0 (1) |
| Times strain to begin urination | 0.6 (1.2) | 0 (1) |
| How do you feel about your condition | 4.4 (1.3) | 0 (1) |
| Age (years) | 56.5 (14.5) | 52.9 (16) |
| Weight (Kg) | 81.9 (22.1) | 73.6 (19) |
| Waist circumference (Cm) | 100.3 (17.9) | 98.0 (35.6) |
| Post-void residual volume (ml) | 44.6 (59.5) | 29.1 (42.5) |
| Number of culture-proven UTIs in past 12 months | 0.5 (1.2) | 0 (1) |
| Body mass index (BMI) | 30.5 (7.9) | 27.7 (7.1) |
| POP-Q Ba measurement | -1.6 (1.9) | -2 (1) |
| POP-Q C measurement | -6 (4.7) | -7.3 (1.5) |
| Number of times pregnant | 2.5 (1.9) | 1.7 (1.7) |
| Number of vaginal deliveries | 1.7 (1.4) | 1 (1.3) |
| Functional comorbidity index total | 2.4 (2.2) | 1 (1.2) |
| GUPI urine [53] | 4.2 (2.7) | 1.7 (1.9) |
| POPDI-6 | 16.8 (19.3) | 5.6 (9.8) |
| CRADI-8 | 19.8 (20.1) | 9.3 (12.4) |
| UDI-6 [81] | 42.1 (24.4) | 12.3 (16.1) |
| Perceived stress scale [82] | 12.9 (7.6) | 13.7 (6.6) |
| PROMIS constipation T-score | 51.3 (8.8) | 50 (10) |
| PROMIS depression T-score | 49.4 (8.8) | 50 (10) |
| PROMIS anxiety T-score | 50.3 (9.1) | 50 (10) |
| PROMIS sleep disturbance T-score | 53.3 (8.7) | 50 (10) |

| | | |
|---|---|---|
| PROMIS diarrhea T-score | 48.9 (9.5) | 50 (10) |
| PROMIS physical functioning T-score [83] | 47.5 (10.3) | 50 (10) |
| Bladder diary composite variables [58] | | |
| Average number of voids in 24 hours | 8.3 (3.3) | 5.7 (2.4) |
| Average voided volume in 24 hours (ml) | 1800.8 (699.7) | 1355.9 (556.3) |
| Average number of Intakes in 24 hours | 6.3 (2.3) | 6.0 (3.5) |
| Average intake volume in 24 hours (ml) | 1739.4 (679.6) | 1541.9 (562.1) |
| Max voided volume (ml) | 526.1 (207.3) | 474.7 (214.2) |
| Hispanic | 21 (4%) | 3 (5%) |
| Race | | |
| American Indian/Alaskan Native | 7 (1%) | 0 (0%) |
| Asian/Asian American | 16 (3%) | 2 (3%) |
| Black/African American | 66 (12%) | 10 (16%) |
| Native Hawaiian or Pacific Islander | 1 (0%) | 0 (0%) |
| White | 452 (83%) | 53 (83%) |
| Other | 7 (1%) | 0 (0%) |
| Unknown | 5 (1%) | 0 (0%) |
| Education | | |
| Less than HS diploma/GED | 12 (2%) | 0 (0%) |
| HS diploma/GED | 47 (9%) | 7 (12%) |
| Some college or tech school, no degree | 126 (23%) | 5 (8%) |
| Associates degree | 63 (12%) | 8 (14%) |
| Bachelors degree | 158 (29%) | 23 (39%) |
| Graduate degree | 130 (24%) | 10 (17%) |
| Education: unknown | 9 (2%) | 6 (10%) |
| Employment status | | |
| Employed part-time | 76 (14%) | 10 (17%) |
| Employed full-time | 207 (38%) | 32 (53%) |
| Unemployed (looking for work) | 15 (3%) | 1 (2%) |
| Not employed (not looking for work, includes stay-at-home, retired) | 241 (44%) | 14 (23%) |
| Employment status: unknown | 6 (1%) | 3 (5%) |
| Marital status | | |
| Married/civil union | 306 (56%) | 27 (44%) |
| Living with a partner | 19 (3%) | 2 (3%) |
| Separated or divorced | 91 (17%) | 6 (10%) |
| Widowed | 42 (8%) | 0 (0%) |
| Single, never married | 84 (15%) | 17 (27%) |
| Marital Status: unknown | 3 (1%) | 10 (16%) |
| Number of alcoholic drinks per week | | |
| 0 to 3 drinks | 361 (66%) | 44 (70%) |
| 4 to 7 drinks | 66 (12%) | 7 (11%) |
| 8 to 14 drinks | 15 (3%) | 3 (5%) |

| | | |
|---|---|---|
| 14 or more drinks | 3 (1%) | 0 (0%) |
| Has not had alcohol in the past | 90 (17%) | 9 (14%) |
| Drinks per week: unknown | 10 (2%) | 0 (0%) |
| Smoking status | | |
| Current smoker | 36 (7%) | 0 (0%) |
| Former smoker | 155 (28%) | 21 (33%) |
| Never smoker | 349 (64%) | 42 (67%) |
| Smoking status: unknown | 5 (1%) | 0 (0%) |
| Recreational drug use status | | |
| Current recreational drug user | 24 (4%) | 1 (2%) |
| Former recreational drug user | 84 (15%) | 9 (14%) |
| Never recreational drug user | 427 (78%) | 53 (84%) |
| Recreational drug uses status: unknown | 10 (2%) | 0 (0%) |
| Comorbidities [56] | | |
| Arthritis | 231 (42%) | 22.9% |
| Osteoporosis | 69 (13%) | 5.6% |
| Asthma | 106 (19%) | 4.3% |
| COPD | 32 (6%) | 5.9% |
| Angina | 19 (3%) | 7.3% |
| Congestive heart failure | 28 (5%) | 2.3% |
| Heart attack | 11 (2%) | 6.2% |
| Neurological disease | 2 (0%) | 2.3% |
| Stroke | 22 (4%) | 3.8% |
| Peripheral vascular disease | 14 (3%) | 3.1% |
| Diabetes | 76 (14%) | 6.6% |
| Upper gastrointestinal disease | 152 (28%) | 12.1% |
| Depression | 188 (34%) | 18.5% |
| Anxiety or panic disorder | 139 (26%) | 5.2% |
| Visual impairment | 91 (17%) | 5.4% |
| Hearing impairment | 26 (5%) | 10.9% |
| Degenerative disc disease | 115 (21%) | 5 (8%) |
| Positive bacterial culture | 54 (10%) | 3 (5%) |
| Participant had more than two UTIs | 258 (47%) | 13 (21%) |
| Participant has a history of pelvic pain | 76 (14%) | 1 (2%) |
| Post-menopausal | 353 (65%) | 32 (51%) |
| Sexually active within the last month | 240 (44%) | 27 (44%) |
| Participant has had a UTI | 0 (0%) | 6 (10%) |
| Participant has had an STI | 0 (0%) | 2 (3%) |
| Participant has had a genital infection | 0 (0%) | 0 (0%) |
| Participant has a history of hormone treatment/use | 0 (0%) | 4 (6%) |
| History of hypertension | 209 (38%) | 11 (17%) |
| Participant has hyperlipidemia | 174 (32%) | 11 (17%) |
| Participant has diabetes | 78 (14%) | 3 (5%) |

| | | |
|---|---|---|
| Participant has sleep apnea | 95 (17%) | 5 (8%) |
| Participant has a psychiatric diagnosis | 235 (43%) | 10 (16%) |
| Participant has colorectal disease | 55 (10%) | 2 (3%) |
| Participant has other medical problems | 339 (62%) | 20 (32%) |
| History of bladder or urethral trauma | 9 (2%) | 0 (0%) |
| Participant has undergone a surgery for their LUTS | 81 (15%) | 0 (0%) |
| Participant has had a hysterectomy | 167 (31%) | 7 (11%) |
| Participant has had a C-section | 83 (15%) | 14 (22%) |
| Participant has had spinal or brain surgery | 43 (8%) | 2 (3%) |
| Participant has had rectal surgery | 30 (6%) | 1 (2%) |
| Participant has had other surgical procedures done | 463 (85%) | 43 (68%) |
| Participant is currently taking a medication | 496 (91%) | 44 (70%) |
| Participant has used antibiotics in the past 3 months | 200 (37%) | 9 (15%) |
| Participant has used antifungal medication in the past 3 months | 60 (11%) | 1 (2%) |
| Physical exam findings | | |
| Introitus findings: inflammation | 1 (0%) | 0% |
| Introitus findings: atrophic | 88 (16%) | 0% |
| Introitus findings: other | 26 (5%) | 0% |
| Introitus findings: none | 414 (76%) | 0% |
| Introitus findings: unknown | 17 (3%) | 100% |
| Urethra findings: mass/diverticulum | 0 (0%) | 0% |
| Urethra findings: caruncle | 12 (2%) | 0% |
| Urethra findings: other | 51 (9%) | 0% |
| Urethra findings: none | 461 (85%) | 100% |
| Urethra findings: unknown | 19 (3%) | 0% |
| Vagina findings: lesion/erosion | 0 (0%) | 0% |
| Vagina findings: other | 136 (25%) | 0% |
| Vagina findings: none | 395 (72%) | 100% |
| Vagina findings: not done | 15 (3%) | 0% |
| Uterus findings: absent | 127 (23%) | 0% |
| Uterus findings: mass | 0 (0%) | 0% |
| Uterus findings: other | 44 (8%) | 0% |
| Uterus findings: none | 334 (61%) | 100% |
| Uterus findings: unknown | 33 (6%) | 0% |
| Rectal exam findings: mass | 0 (0%) | 0% |
| Rectal exam findings: resting tone | 8 (1%) | 0% |
| Rectal exam findings: contraction strength | 7 (1%) | 0% |
| Rectal exam findings: other | 48 (9%) | 0% |
| Rectal exam findings: none | 290 (53%) | 100% |
| Rectal exam findings: unknown | 187 (34%) | 0% |
| Female tenderness: abdomen | 5 (1%) | 0% |
| Female tenderness: flank | 0 (0%) | 0% |
| Female tenderness: suprapubic | 3 (1%) | 0% |

| | | |
|---|---|---|
| Female tenderness: groin | 1 (0%) | 0% |
| Female tenderness: clitoris | 0 (0%) | 0% |
| Female tenderness: labia minora/majora | 0 (0%) | 0% |
| Female tenderness: introitus | 3 (1%) | 0% |
| Female tenderness: urethra | 3 (1%) | 0% |
| Female tenderness: cervix/uterus | 3 (1%) | 0% |
| Female tenderness: ovaries | 1 (0%) | 0% |
| Female tenderness: rectum | 1 (0%) | 0% |
| Female tenderness: pelvic floor | 61 (11%) | 0% |
| Female tenderness: other | 26 (5%) | 0% |
| Female tenderness: none | 417 (77%) | 100% |
| Female tenderness: unknown | 26 (5%) | 0% |

Abbreviations: Ba, point B anterior; BMI, body mass index; C, cervix/vaginal cuff; COPD, chronic obstructive pulmonary disease; CRADI, colorectal-anal distress inventory; C-section, cesarean delivery; GED, general educational development; GUPI, genitourinary pain index; HS, high school; LUTS, lower urinary tract symptoms; POPDI, pelvic organ prolapse distress inventory; POP-Q, pelvic organ prolapse quantification; PROMIS, patient-reported outcomes measurement information system; Std, standard deviation; STI, sexually transmitted infection; UDI, urinary distress inventory; UTI, urinary tract infection.

### Simple example illustrating the benefits of scaling by controls

Since the need for scaling is not universally understood, let us illustrate it by a simple example (simulated example 1). Imagine a situation of two disease subtypes A and B, each described by two variables $X_1$ and $X_2$. Let variable $X_1$ be normally distributed with mean value $m_{1A} = 2$ and standard deviation $\sigma_{1A} = 1$ in subtype A, and normally distributed with mean value $m_{1B} = 4$ and standard deviation $\sigma_{1B} = 1$ in subtype B. Let variable $X_1$ be normally distributed with mean $m_{1HC} = 1$ and standard deviation $\sigma_{1HC} = 0.5$ in healthy controls. Similarly, let variable $X_2$ be normally distributed in subtypes A, B, and healthy controls with mean values $m_{2A} = 200, m_{2B} = 202, m_{2HC} = 100$ and standard deviations $\sigma_{2A} = 100, \sigma_{2B} = 100, \sigma_{2HC} = 50$. Note that absolute difference in the mean values of variables $X_1$ and $X_2$ in subtypes A and B are equal $\Delta m_1 = \Delta m_2 = 2$, while the relative differences and differences scaled by standard deviations are quite different: $\frac{\Delta m_1}{m_{1A}} = 1, \frac{\Delta m_2}{m_{2A}} = 0.01$; $\frac{\Delta m_1}{\sigma_{1A}} = 2, \frac{\Delta m_2}{\sigma_{2A}} = 0.02$. These distributions are presented in Supplemental Figure S1. To illustrate how clustering works with different scaling approaches, we simulated 100 objects from cluster A and 100 objects from cluster B with uncorrelated variables $X_1$ and $X_2$, following the above distributions by using

MATLAB function multivariate normal random numbers distribution (mvnrnd). Pairwise distances between the objects were calculated using three approaches to scaling of variables: unscaled, scaled as cohort's z-scores, and scaled by healthy controls described by eq. (1). Then clustering of the simulated objects was performed by MATLAB function k-means using the above three scaling approaches. Pairwise distances together with misclassification errors are presented in Supplemental Table S2.

**Supplemental Figure S1. Simulated example 1, illustrating distributions of two variables describing patients in two disease subtypes and healthy controls. Upper row – $X_1$, lower row $X_2$. Left column unscaled, center z-scored, right scaled by controls.**

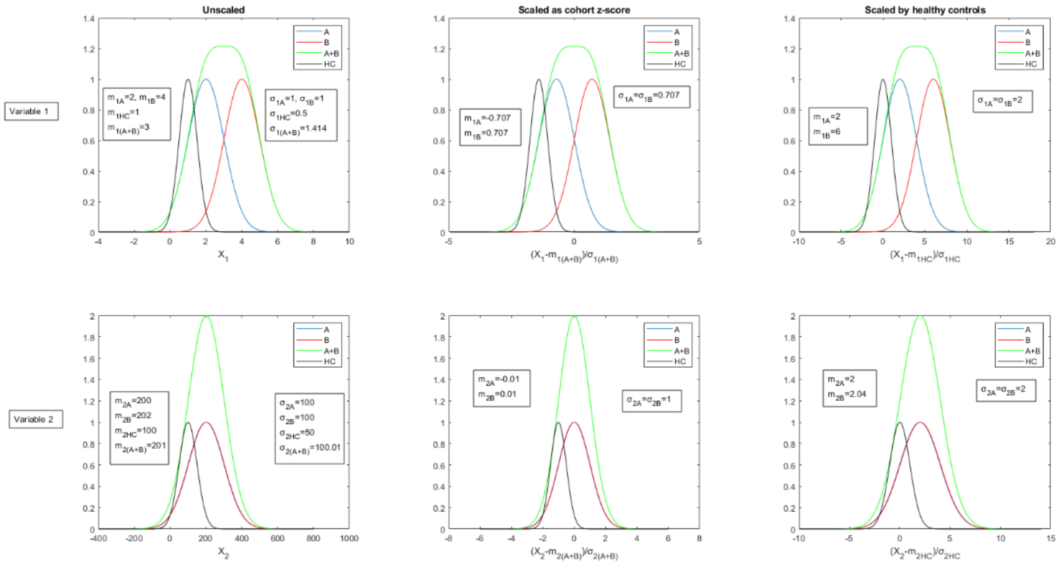


**Supplemental Table S2. Means, standard deviations, pairwise distances, and misclassification errors resulting from the use of three scaling approaches (unscaled – U, scaled as cohort's z-scores – C, and scaled by healthy controls – N).**

| | $m_{1A}(\sigma_{1A})$ | $m_{1B}(\sigma_{1B})$ | $m_{2A}(\sigma_{2A})$ | $m_{2B}(\sigma_{2B})$ | $PD_{1int}$ | $PD_{2int}$ | $PD_{int}$ | $PD_{1ext}$ | $PD_{2ext}$ | $PD_{ext}$ | $Err,\%$ |
|---|---|---|---|---|---|---|---|---|---|---|---|
| $U$ | 2(1) | 4(1) | 200(100) | 202(100) | 1.13 | 11.3 | 11.5 | 2.09 | 11.4 | 11.8 | 45.6 |
| $C$ | -0.71(0.71) | 0.71(0.71) | -0.01(0.01) | 0.01(0.01) | 0.95 | 1.13 | 1.63 | 1.55 | 1.13 | 2.10 | 22.4 |
| $N$ | 2(2) | 6(2) | 2(2) | 2.04(2) | 1.59 | 1.59 | 2.50 | 4.03 | 1.59 | 4.54 | 8.0 |

For the case of unscaled variables, the mean pairwise distance between the objects within a cluster $PD_{int}$ is dominated by the distance $PD_{2int}$ along the variable $X_2$, which is 10-fold larger than along $X_1$. Since distributions for subtype A and B are nearly completely overlapping along $X_2$, $PD_{2int} \approx PD_{2ext}$ ,

$PD_{int} \approx PD_{ext}$. Clustering algorithms seek to minimize the sum of within-cluster differences that are dominated by the second variable with a result of misclassification error of 45.6% that is only slightly better than 50% error based on classification by pure chance. Clearly, using unscaled variables does not result in proper clustering in this example. For the case of scaling as cohort's z-scores, the situation is better since the pairwise distances along both variables are commensurate; however, the increased standard deviation for the whole cohort along $X_1$ leads to the decreased internal and external differences across this variable relative to the second variable. The k-means clustering algorithm is trying to minimize distances along both directions but to a larger extent along $X_2$, resulting in misclassification error of 22.4%. When scaled by healthy controls, within-cluster distances are equal along both variables, while the between cluster distance is dominated by the first variable, where the relative distance (and scaled distance) between the clusters is higher. Clustering works much better with misclassification error of 8%.

### Contrast criterion (CC): special cases

The terms in eq. (15) can be rearranged into eq. (15b):

$$CC = \frac{1}{K} \cdot \sum_{k=1}^{K} \frac{1}{N_k} \sum_{n=1}^{N_k} \left\{ \frac{\sum_{q=1,q\neq n}^{N_k} M_{nq}}{N_k - 1} - \frac{\sum_{i=1,i\neq k}^{K} \sum_{q=1}^{N_i} M_{nq}}{N - N_k} \right\} \tag{15a}$$

where the first term in the brackets represents the averaged probability for $n^{th}$ object assigned to cluster $k$ to be together with any other of $N_k - 1$ objects assigned to cluster $k$; while the second term is the averaged probability for $n^{th}$ object to be together with $N - N_k$ objects not assigned to this cluster. Note that the sum of these terms is not necessary equal to one, since there is the possibility of the third outcome, i.e., that in some instances of k-means, object $n$ is not grouped together with any other objects but constitutes a cluster of its own.

Now let us generalize our definition of CC to the case where at least one of the alleged clusters might contain only one object.

$$N_j = 1, CC = \frac{1}{K}\left\{\left[\sum_{k=1,k\neq j}^{K}\frac{1}{N_k}\sum_{n=1}^{N_k}\left(\frac{\sum_{q=1,q\neq n}^{N_k}M_{nq}}{N_k-1} - \frac{\sum_{i=1,i\neq k}^{K}\sum_{q=1}^{N_i}M_{nq}}{N-N_k}\right)\right] - \frac{\sum_{i=1,i\neq j}^{K}\sum_{q=1}^{N_i}M_{nq}}{N-1}\right\} \quad (15b)$$

Here, for the alleged cluster number $j$, which consists of only one object, the first term of eq. (15a) is equal to zero. It is not an unlikely situation, especially if the number of variables is high and commensurate with the number of objects $N$. Imagine the case of $N$ objects described by $N$ binary variables. Now let $i^{th}$ variable equal one for $i^{th}$ object and equal zero for all other objects. In this case, there will be no reason to group any objects together, and each of them will be a cluster of its own. Such a situation might occur when clustering patients by their Single Nucleotide Polymorphism (SNPs). In this case, eq. 15a will be reduced to:

$$CC = -\frac{\sum_{n=1}^{N}\sum_{q=1,q\neq n}^{N}M_{nq}}{N(N-1)} \quad (15c)$$

In the ideal case described above, all the non-diagonal elements of M are equal to zero, so $CC$=0. In the real-life situation, the presence of this negative term ensures the cases of multiple small clusters are not favored by the contrast criterion.

### Contrast criterion and proportion of ambiguous clustering: simulation example

To illustrate the properties of contrast criterion (CC) and compare it with proportion of ambiguous clustering (PAC), we simulated the following idealized simplified example A. Assume we are analyzing omics data, e.g., 40,000 transcript (or gene expression) levels. Assume we have these data for 100 people, consisting of five groups of equal size. Assume that, for each group of people, there is a group of 4,000 upregulated transcripts (signatures of the subtype of disease). Assume that these groups of transcripts are non-overlapping. Therefore, there are 20,000 differentially abundant transcripts across five groups of people, the remaining 20,000 transcripts being similarly distributed in all five groups.

Assume that distribution of transcript levels in the five groups of people is described by the multivariate normal distribution with matrix of mean values $\mu(5 \times 40000)$ and correlation matrix $\Sigma(40000 \times 40000)$. Supplemental Figure S2 presents $\mu$ and $\Sigma$ for the first example. Variables are already standardized so $\mu = 2$ correspond to mean equal two standard deviations or the effect size =2. Note that transcript levels are simulated to be highly correlated (R=0.9) within the 4,000-transcript signatures (yellow squares on the diagonal of the correlation matrix, Supplemental Figure S2) and uncorrelated outside of the signatures. The right side panel of the figure demonstrates an example of a single instance of the simulated distribution of transcripts abundances for members of each of five groups (color coded as blue, red, black, cyan, and purple) of people.

**Supplemental Figure S2. Simulated example A. Non-overlapping transcript signatures. 40,000 transcripts. Five groups of people. Means, correlation matrix, and single instance of simulated distributions of transcript abundances.**

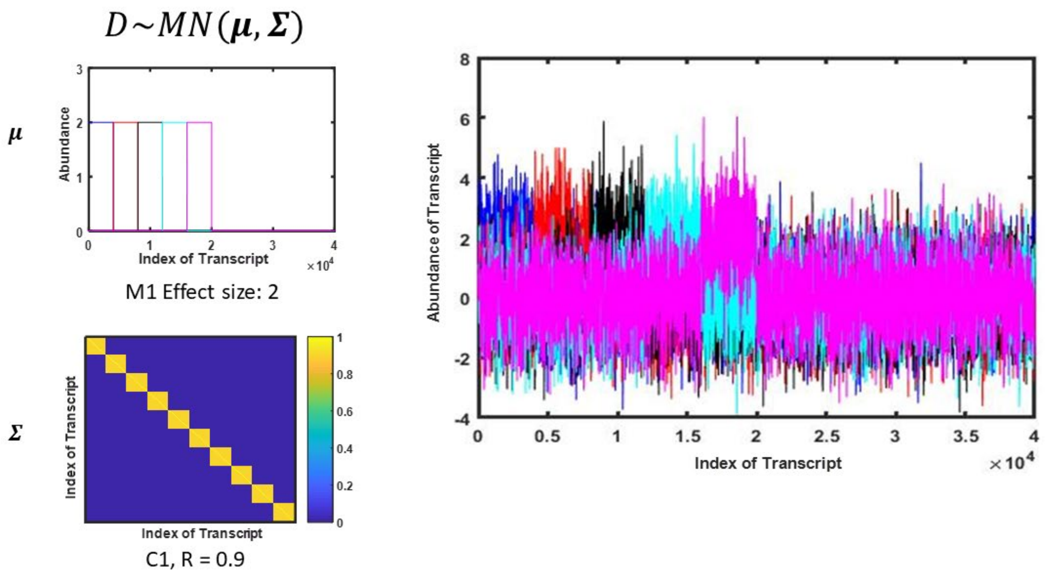


Supplemental Figure S3 illustrates consensus matrices generated when performing resampling-based consensus clustering [34] described in the “Consensus clustering using continuous variables” subsection of the manuscript. The number of clusters K is scanned from 2 to 8. For this example of high effect size=2 (standardized mean values for each of differentially abundant transcript=2) , the optimal number of clusters K=5 is obvious both from visual inspection of consensus matrices and from the values of CC (maximum at K=5) and values of PAC with ambiguity range (0.1, 0.9) and more liberal range (0.2,0.8), minima at K=5. Therefore, all these criteria work equally well in this case. However, if the effect size is reduced to 0.6 (remaining conditions are the same), the clustering decision becomes more complicated. Visual inspection of the consensus matrices presented in Supplemental Figure S4 do not reveal clear optimum, PAC reaches minimum at K=8, and only CC provides the right number of clusters K=5. Supplemental Figures S5 and S6 demonstrate the dependence of misclassification error on the effect size and the level of correlation between the transcripts' abundance levels. Three clustering methods are compared: resampling-based consensus clustering with contrast criterion, k-means with Calinski-Harabasz criterion, and hierarchical clustering with Calinski-Harabasz criterion. Three panels illustrate cases with different levels of correlation between transcript abundances R=0.1, 0.45, and 0.9. Supplemental Figure S6 differs from Supplemental Figure S5 by the pattern of correlation between the transcripts. In Supplemental Figure S5, members of the signatures are equally correlated (and the rest of the transcripts are not correlated); in Supplemental Figure S6, transcripts are arranged in a way that neighbors in the correlation matrix are correlated according to the following equation: $R_{ij} = r^{|i-j|}$ , where *i,j* are transcript indices, and r=0.1, 0.45, and 0.9 (simulated example A1). As seen through comparison of the panels in Supplemental Figures S5 and S6, misclassification error depends both on the level and on the pattern of correlation between the variables, which is in concert with simulations of [63]. Importantly, in all of the cases, consensus clustering with contrast criterion outperforms the other two methods, allowing for reliable clustering at much lower effect size and higher correlation levels.

**Supplemental Figure S3. Simulated example A. Consensus matrices, K=2,3…8. CC vs. PAC. Effect size=2.**

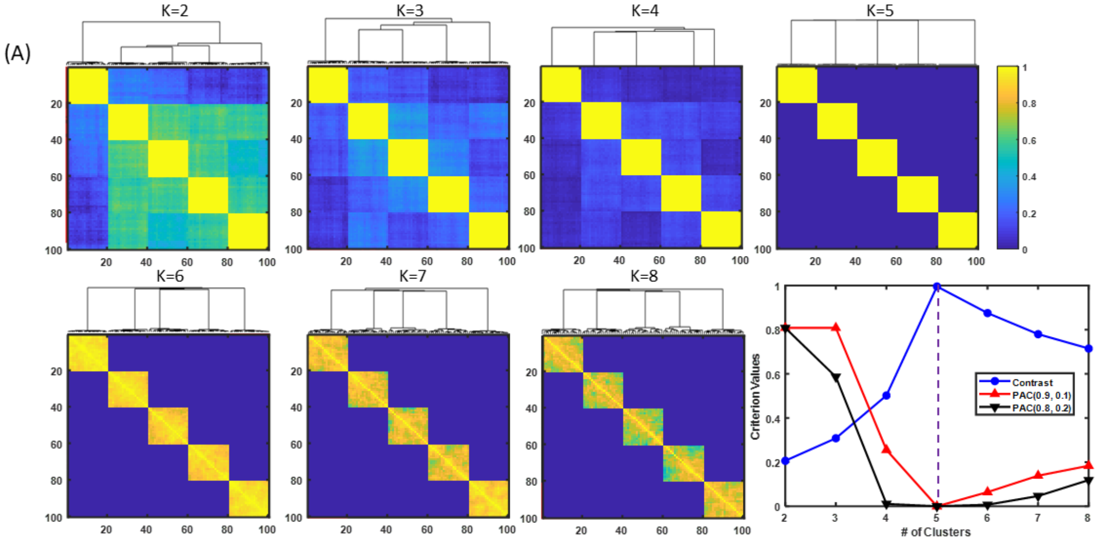


**Supplemental Figure S4. Simulated example A. Consensus matrices, K=2,3…8. CC vs. PAC. Effect size=0.6.**

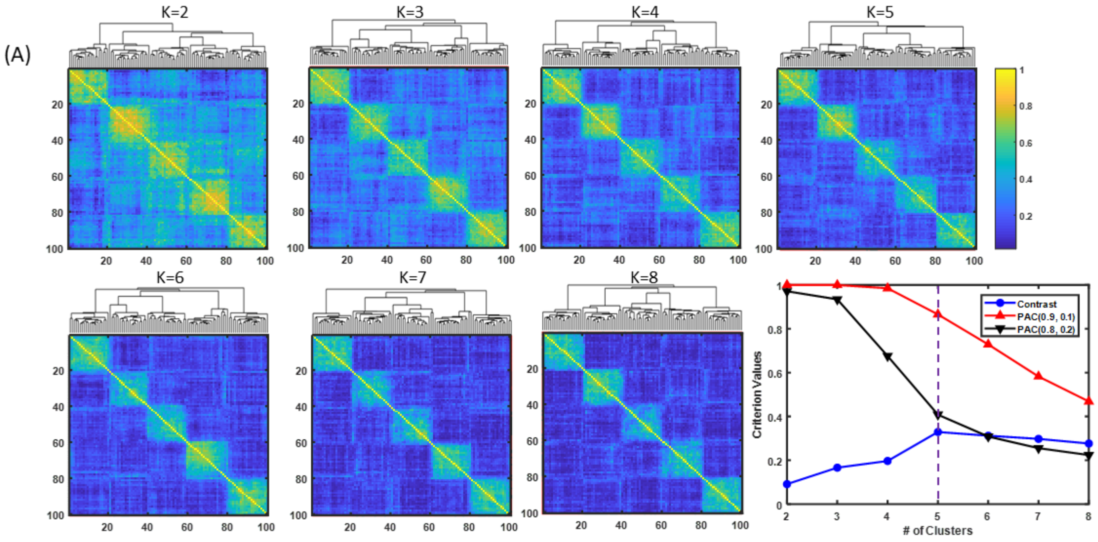

**Supplemental Figure S5. Simulated example A. Misclassification error vs. effect size. Comparison of consensus clustering using contrast criterion with k-means and hierarchical clustering using Calinski-Harabasz criterion. Three values of correlation coefficient for transcript abundances within transcript signatures.**

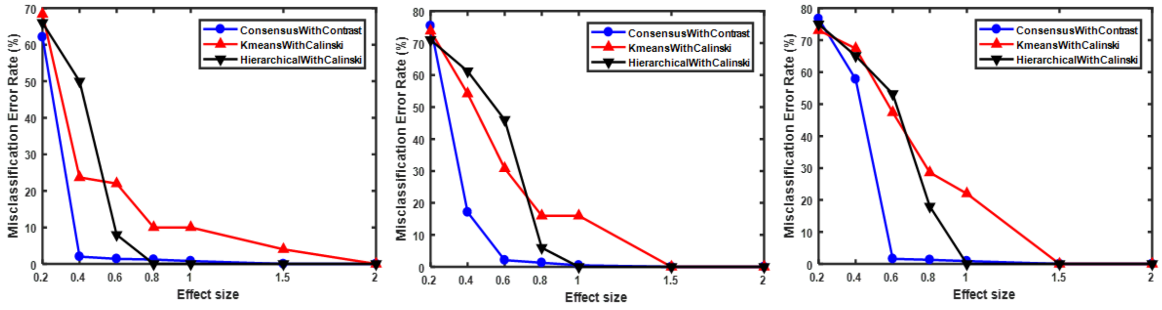


R=0.1 R=0.45 R=0.9

**Supplemental Figure S6. Simulated example A1. Same conditions as in simulated examples A, except transcript abundances being correlated following $R_{ij} = r^{|i-j|}$ .**

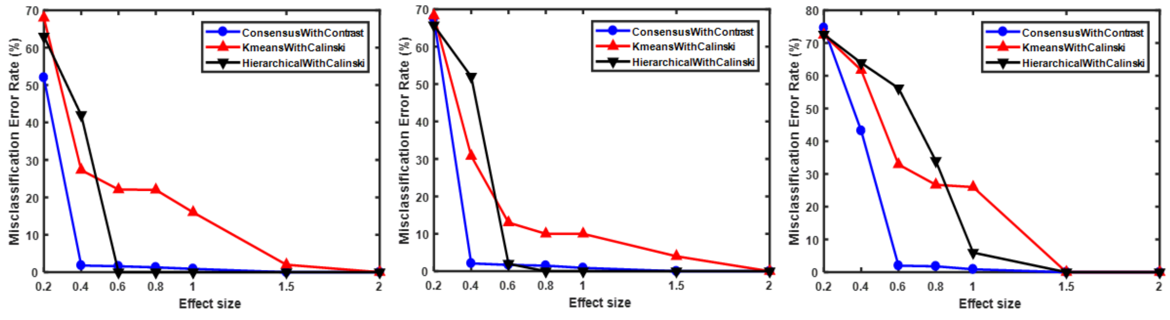


r=0.1 r=0.45 r=0.9

Another simulated example (B) illustrates that CC outperforms PAC, and that clustering with CC outperforms k-means and hierarchical clustering, with Calinski-Harabasz criterion for the case of overlapping signatures as well. The five clusters of patients were simulated using multivariate normal distributions with different mean vectors. Signatures for the clusters were different but completely overlapping, meaning that only for the first 4,000 of 40,000, the mean values were different across the clusters. We simulated different but overlapping signatures by assuming that, in cluster 1, all transcripts were equally upregulated; in cluster 2, all transcripts were equally down-regulated. In clusters 3 , 4, and 5, transcripts were intermittently up- and down-regulated in the following patterns: +-+-+-+-… in cluster 3, -+-+-+-+… in cluster 4, and ++--++--… in cluster 5. Supplemental Figure S7 illustrates the case of the above overlapping signatures with $\mu = 2$ (effect size =2 for all 4,000 transcripts in the signatures). In this case of high effect size, visual inspection of consensus matrices, CC, and PAC criterion work equally well in determining K=5 as a number of clusters. However, in case of lower effect size $\mu = 0.8$, visual inspection of consensus matrices (Supplemental Figure S8) does not provide an unambiguous answer, but indicates 4≤K≤7. PAC (0.1,0.9) indicates K=8, while CC and PAC (0.2, 0.8) indicate correct value K=5. In this case of overlapping signatures (in Supplemental Figure S9, members of the biomarker signatures are equally correlated; in Supplemental Figure S10, correlation is described by equation $R_{ij} = r^{|i-j|}$ [simulated example B1]), consensus clustering with CC works well, providing a misclassification error below 5% for $\mu \geq 0.8$, for all simulated patterns and levels of correlation between transcript abundances, while two other methods result in an unacceptably high level of 60%, even for a high effect size of 2.

**Supplemental Figure S7. Simulated example B. Overlapping transcript signatures. Consensus matrices, K=2,3…8. CC vs. PAC. Effect size=2.**

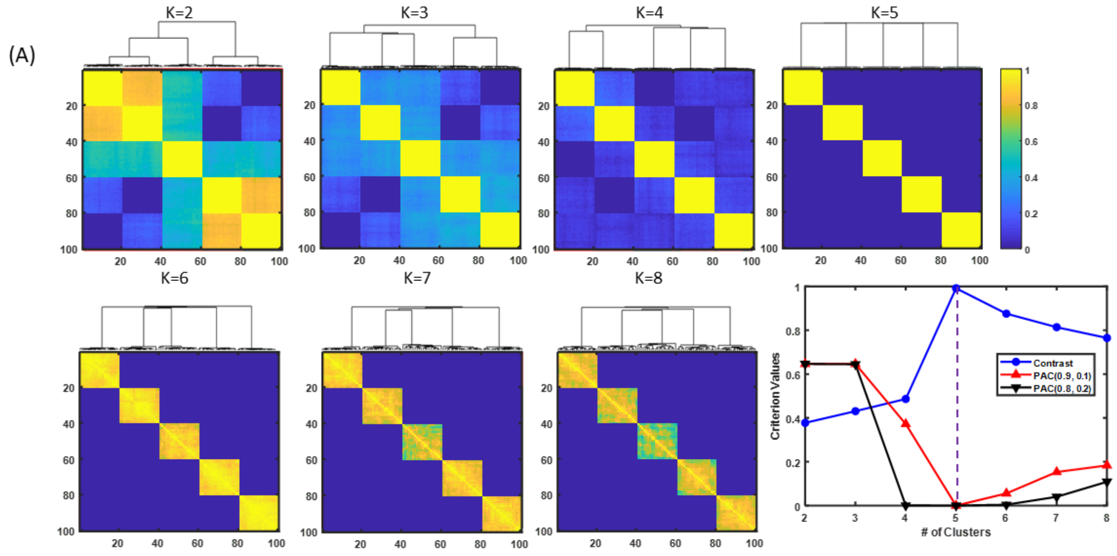


**Supplemental Figure S8. Simulated example B. Overlapping transcript signatures. Consensus matrices, K=2,3…8. CC vs. PAC. Effect size=0.8.**

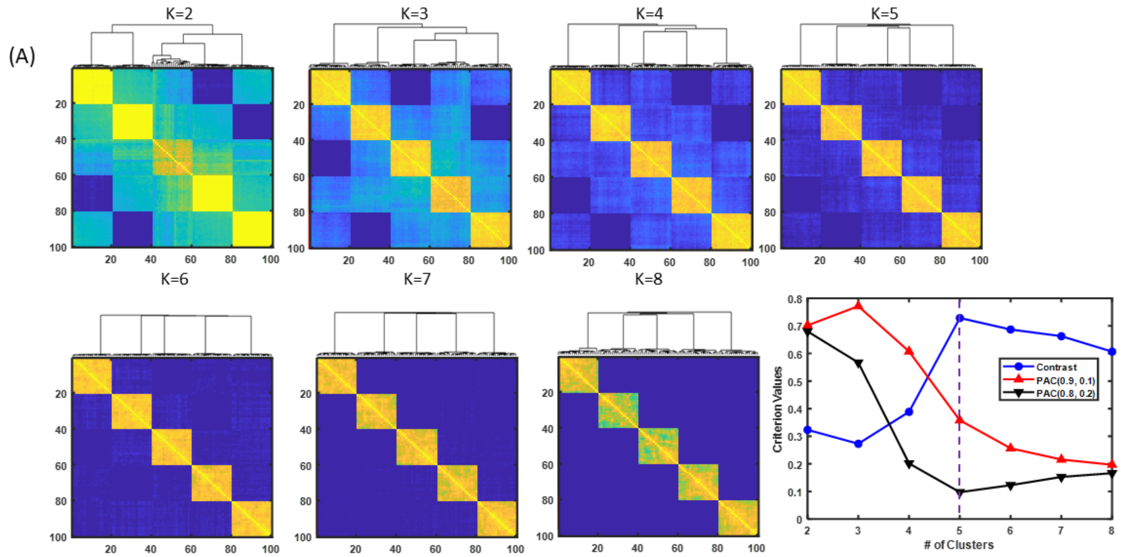

**Supplemental Figure S9. Simulated example B. Misclassification error vs. effect size. Comparison of consensus clustering using contrast criterion with k-means and hierarchical clustering using Calinski-Harabasz criterion. Three values of correlation coefficient for transcript abundances within transcript signatures.**

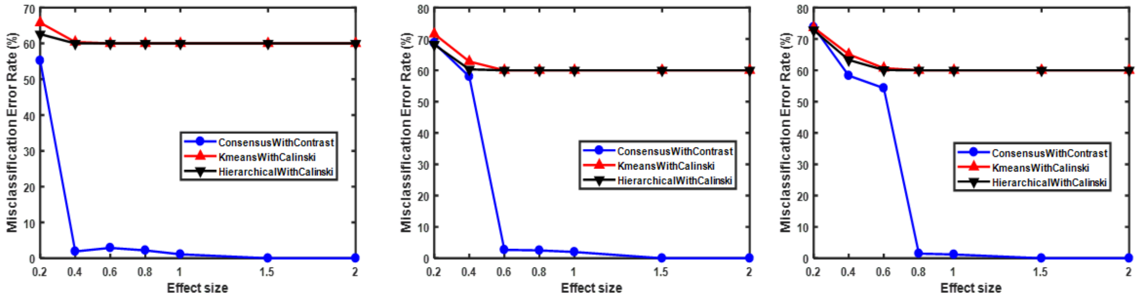


R=0.1 R=0.45 R=0.9

**Supplemental Figure S10. Simulated example B1. Same conditions as in simulated examples B, except transcript abundances being correlated following $R_{ij} = r^{|i-j|}$.**

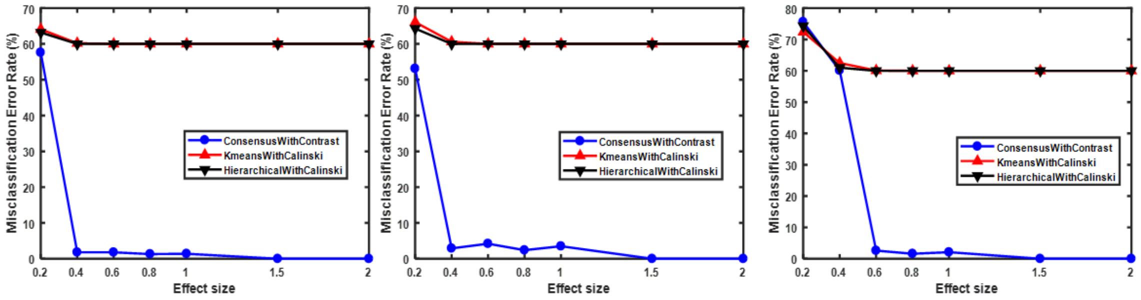


r=0.1 r=0.45 r=0.9

### Details on the targeted proteomics study of serum samples of women with LUTS versus controls

**Supplemental Table S3. Lists of differentially abundant proteins observed in the serum samples of 230 women with LUTS versus controls. Proteins with false discovery rate (FDR) adjusted p-value<0.05 are bolded.**

**Supplemental Table S3A. All 230 women with LUTS.**

| Protein | $mean(\log_2 \frac{C_{cases}}{C_{controls}})$ | P-Value |
|---|---|---|
| IGFBP3 | -0.2099 | 0.0018 |
| DPP4 | -0.1689 | 0.0083 |
| SCF | -0.1409 | 0.0297 |
| DNER | -0.1297 | 0.0244 |
| TIE1 | -0.1091 | 0.0309 |
| UMOD | -0.1080 | 0.0165 |
| RSPO1 | 0.1077 | 0.0282 |
| IL_10RB | 0.1246 | 0.0489 |
| EPHB6 | 0.1269 | 0.0326 |
| CLM_6 | 0.1501 | 0.0144 |
| TNF | 0.1527 | 0.0312 |
| CD5 | 0.1565 | 0.0136 |
| CLEC10A | 0.1655 | 0.0372 |
| HGF | 0.1718 | 0.0395 |
| DRAXIN | 0.1790 | 0.0456 |
| SLAMF1 | 0.1807 | 0.0152 |
| SCARB2 | 0.1824 | 0.0185 |
| PD_L1 | 0.1861 | 0.0013 |
| SKR3 | 0.1862 | 0.0017 |
| CD38 | 0.1896 | 0.0190 |
| CDCP1 | 0.2407 | 0.0183 |
| FcRL2 | 0.2574 | 0.0153 |
| IL_10RA | 0.2689 | 0.0004 |
| TGF_alpha | 0.2710 | 0.0406 |
| MCP_3 | 0.2738 | 0.0040 |
| GCP5 | 0.2764 | 0.0263 |
| MSR1 | 0.2780 | 0.0054 |
| NAAA | 0.2899 | 0.0238 |
| IL12 | 0.2931 | 0.0151 |
| TNFSF14 | 0.2939 | 0.0219 |
| **KYNU** | **0.3428** | **<.0001** |
| CCL19 | 0.3517 | 0.0133 |

| | | |
|---|---|---|
| **Alpha_2_MRAP** | **0.4033** | **<.0001** |

**Supplemental Table S3B. W1 cluster (n=37).**

| **Protein** | $mean(\log_2 \frac{C_{W1\,cases}}{C_{controls}})$ | **P-Value** |
|---|---|---|
| SCF | -0.2485 | 0.0147 |
| LTBP2 | -0.1877 | 0.0481 |
| SPOCK1 | 0.1409 | 0.0339 |
| EPHB6 | 0.1486 | 0.0314 |
| SKR3 | 0.1610 | 0.0302 |
| RSPO1 | 0.1645 | 0.0215 |
| PD_L1 | 0.1708 | 0.0170 |
| SCARB2 | 0.2104 | 0.0390 |
| CX3CL1 | 0.2129 | 0.0279 |
| TNF | 0.2269 | 0.0233 |
| CD38 | 0.2295 | 0.0173 |
| SMOC2 | 0.2348 | 0.0449 |
| MSR1 | 0.2750 | 0.0358 |
| KYNU | 0.3502 | 0.0033 |
| Alpha_2_MRAP | 0.3587 | 0.0268 |
| IL_10RA | 0.4153 | 0.0136 |
| CCL19 | 0.4583 | 0.0217 |

**Supplemental Table S3C. W2 cluster (n=38).**

| **Protein** | $mean(\log_2 \frac{C_{W2\,cases}}{C_{controls}})$ | **P-Value** |
|---|---|---|
| IFN_gamma | -0.4753 | 0.0426 |
| DPP4 | -0.2063 | 0.0108 |
| DNER | -0.1932 | 0.0092 |
| TIE1 | -0.1273 | 0.0434 |
| CLM_6 | 0.1674 | 0.0200 |
| KYNU | 0.1990 | 0.0378 |
| CLEC10A | 0.2075 | 0.0388 |
| NAAA | 0.3255 | 0.0218 |
| TNFSF14 | 0.3509 | 0.0316 |
| GCP5 | 0.3911 | 0.0074 |

**Supplemental Table S3D. W3 cluster (n=53).**

| Protein | $mean(\log_2 \frac{C_{W3\,cases}}{C_{controls}})$ | P-Value |
|---|---|---|
| IGFBP3 | -0.2729 | 0.0016 |
| SCF | -0.1897 | 0.0186 |
| DPP4 | -0.1842 | 0.0247 |
| DNER | -0.1834 | 0.0083 |
| PLA2G7 | -0.1633 | 0.0282 |
| MET | -0.1518 | 0.0144 |
| TIE1 | -0.1164 | 0.0398 |
| TNXB | -0.1101 | 0.0407 |
| ENG | -0.1015 | 0.0359 |
| THY_1 | 0.1284 | 0.0300 |
| EPHB6 | 0.1523 | 0.0408 |
| SKR3 | 0.1612 | 0.0217 |
| IL_2RB | 0.1736 | 0.0283 |
| PD_L1 | 0.1740 | 0.0458 |
| SCARB2 | 0.1823 | 0.0496 |
| CTSC | 0.1840 | 0.0438 |
| DRAXIN | 0.2114 | 0.0489 |
| HGF | 0.2336 | 0.0177 |
| MCP_3 | 0.2749 | 0.0160 |
| MSR1 | 0.2805 | 0.0228 |
| TGF_alpha | 0.3084 | 0.0438 |
| CCL19 | 0.3493 | 0.0488 |
| OSM | 0.3546 | 0.0459 |
| NAAA | 0.3593 | 0.0220 |
| **KYNU** | **0.4350** | **<.0001** |
| Alpha_2_MRAP | 0.4445 | 0.0010 |

**Supplemental Table S3E. W4 cluster (n=42).**

| Protein | $mean(\log_2 \frac{C_{W4\,cases}}{C_{controls}})$ | P-Value |
|---|---|---|
| DPP4 | -0.1796 | 0.0108 |
| AOC3 | -0.1749 | 0.0277 |
| IGFBP3 | -0.1736 | 0.0302 |

| | | |
|---|---|---|
| TIE1 | -0.1310 | 0.0195 |
| UMOD | -0.1206 | 0.0437 |
| IL_2RB | 0.1356 | 0.0473 |
| PD_L1 | 0.1676 | 0.0288 |
| GCP5 | 0.3160 | 0.0294 |
| TGF_alpha | 0.3171 | 0.0435 |
| OSM | 0.3616 | 0.0319 |

**Supplemental Table S3F. W5 cluster (n=60).**

| Protein | $mean(\log_2 \frac{C_{W5\,cases}}{C_{controls}})$ | P-Value |
|---|---|---|
| **IGFBP3** | **-0.2619** | **0.0041** |
| **UMOD** | **-0.1753** | **0.0010** |
| NCAN | -0.1580 | 0.0341 |
| NBL1 | 0.0742 | 0.0168 |
| DDR1 | 0.0981 | 0.0439 |
| CSF_1 | 0.1156 | 0.0218 |
| EZR | 0.1299 | 0.0211 |
| CDH6 | 0.1322 | 0.0241 |
| QPCT | 0.1442 | 0.0415 |
| TNFRSF21 | 0.1444 | 0.0158 |
| SCARA5 | 0.1462 | 0.0128 |
| PDGF_R_alpha | 0.1617 | 0.0156 |
| CD40 | 0.1643 | 0.0296 |
| **FLRT2** | **0.1671** | **0.0035** |
| ICAM1 | 0.1677 | 0.0460 |
| THY_1 | 0.1678 | 0.0056 |
| EPHB6 | 0.1694 | 0.0141 |
| GZMA | 0.1737 | 0.0387 |
| GDNF_1 | 0.1754 | 0.0232 |
| Beta_NGF_1 | 0.1850 | 0.0407 |
| N2DL_2 | 0.1929 | 0.0206 |
| CDH3 | 0.1960 | 0.0353 |
| TNF | 0.2010 | 0.0091 |
| TSLP | 0.2101 | 0.0455 |
| EFNA4 | 0.2139 | 0.0086 |
| LAYN | 0.2280 | 0.0353 |
| **CLM_6** | **0.2282** | **0.0028** |
| gal_8 | 0.2294 | 0.0112 |
| CCL11 | 0.2321 | 0.0168 |
| DRAXIN | 0.2321 | 0.0318 |
| IL_18R1 | 0.2342 | 0.0161 |
| **PD_L1** | **0.2431** | **0.0011** |
| IL_10RB | 0.2459 | 0.0006 |
| CRTAM | 0.2501 | 0.0499 |

| | | |
|---|---|---|
| TIMD4 | 0.2554 | 0.0212 |
| UNC5C | 0.2577 | 0.0161 |
| **JAM_B** | **0.2588** | **0.0030** |
| IL10 | 0.2600 | 0.0415 |
| **CD38** | **0.2799** | **0.0016** |
| **CD5** | **0.2844** | **0.0002** |
| **GFR_alpha_1** | **0.2870** | **0.0014** |
| **CLEC10A** | **0.2872** | **0.0026** |
| **HGF** | **0.2934** | **0.0016** |
| TNFRSF12A | 0.2946 | 0.0105 |
| IGLC2 | 0.2990 | 0.0025 |
| **SMOC2** | **0.3044** | **0.0033** |
| PLXNB1 | 0.3044 | 0.0149 |
| MCP_4 | 0.3100 | 0.0483 |
| **SCARB2** | **0.3120** | **0.0012** |
| **TNFRSF9** | **0.3123** | **0.0035** |
| **SLAMF1** | **0.3188** | **0.0004** |
| EDA2R | 0.3206 | 0.0126 |
| VEGFA | 0.3246 | 0.0140 |
| **SKR3** | **0.3356** | **<.0001** |
| TGF_alpha | 0.3440 | 0.0174 |
| SIGLEC1 | 0.3464 | 0.0093 |
| TNFSF14 | 0.3632 | 0.0112 |
| NAAA | 0.3702 | 0.0164 |
| **VWC2** | **0.3745** | **0.0027** |
| OSM | 0.3750 | 0.0214 |
| **IL18** | **0.4014** | **0.0010** |
| **IL_10RA** | **0.4076** | **0.0006** |
| **FcRL2** | **0.4139** | **0.0006** |
| **IL_12B** | **0.4148** | **0.0049** |

| | | |
|---|---|---|
| **MSR1** | **0.4190** | **0.0003** |
| **KYNU** | **0.4628** | **<.0001** |
| **CDCP1** | **0.4770** | **0.0002** |
| **IL12** | **0.4779** | **0.0013** |
| **MCP_3** | **0.4846** | **<.0001** |
| **CCL19** | **0.5397** | **0.0013** |
| **Alpha_2_MRAP** | **0.7075** | **<.0001** |

### Comparison of the identified clusters W1-W5 with the conventional classification, subtypes, and clusters of women with LUTS identified in literature

It is of interest to compare our clusters W1-W5 with conventional groups and with subtypes of LUTS identified by other researchers. Participants in W1 (predominantly urinary frequency, voiding, and post-micturition symptoms) would not fit into diagnostic categories like overactive bladder (OAB) or stress urinary incontinence (UI). Cluster W2 is defined by the presence of clinically significant prolapse combined with mild LUTS, which was not recognized as a separate subtype in conventional classification of LUTS. Participants in cluster W3 closely resemble the classical definition of ‘wet’ OAB. Unlike cluster W1, these women have urgency incontinence, as well as urinary urgency and frequency, suggesting that OAB wet and dry are different clinical entities. This finding has been demonstrated previously in population-based urodynamic testing studies, where women with urgency incontinence were found to have maximum urethral closure pressure more similar to women with stress incontinence than OAB dry [58, 75]. Participants in cluster W4 have several kinds of incontinence, but mostly stress UI, as their dominating symptom, along with urgency and frequency. This suggests these women might have poor urethral function, given that their storage symptoms are only modest, and voiding symptoms non-existent; therefore, we would hypothesize that these women have poor outlet resistance. Participants in cluster W5 have all LUTS, including voiding, storage, and incontinence reported at a severe degree, suggesting these women might have poor bladder function, as well as poor outlets.

Previous studies have used cluster analyses to characterize women with LUTS [36-37]. Coyne et al. [36] identified six clusters in an analysis of 8505 community-dwelling women from the EPIC study on 14 lower urinary tract symptoms, including seven American Urological Association Symptom Index (AUA-SI) questions. Because that study was population-based, 57% of females reported only minimal urinary symptoms. The remaining five clusters were defined as: (#2) nocturia of twice or more per night (12%); (#3) terminal dribble (10%); (#4) urgency (8%); (#5) stress UI (8%); and (#6) multiple symptoms (5%), including UI (95%), urinary urgency (85%), terminal dribble (43%), incomplete emptying (31%), and weak stream (18%). The latter cluster is similar to our cluster W5, which also includes participants with multiple symptoms at higher level of severity. Importantly, clusters # 2, 3, 4, and 5 are determined by predominant symptoms with low level of other symptoms, while our clusters are defined by combinations of several symptoms. For instance, we have not observed participants reporting nocturia without any other urinary symptoms. It is likely this difference, as well as the absence of minimal symptom cluster in our study, are reflections of the differences in the populations studied in the Symptoms of Lower Urinary Tract Dysfunction Research Network (LURN) and EPIC. LURN participants are patients presenting with bothersome LUTS and seeking treatments of their LUTS. Patients in the specialized urology and urogynecology clinics (LURN cohort) are not only likely to have higher level of severity, but also more complicated combinations of symptoms than people with LUTS in the general population. In addition, the inclusion of the LUTS Tool in the LURN study, together with non-urinary patient-reported outcomes (PROs) and other clinical and bladder diary data, provided higher granularity and allowed for the inclusion of symptoms that might have been missed in a shorter questionnaire.

Cluster analyses of 3167 females in the Boston Area Community Health (BACH) Survey [37] used 14 questions similar to those in the EPIC study described above. Among participants, 24.1% were asymptomatic, and the remainder were assigned to four clusters. Cluster 1 (40.9%) was defined by nocturia, frequency, and UI. Cluster 2 (18.2%) comprised frequency and nocturia. Cluster 3 (10.4%) was differentiated as stress and urgency UI and frequency. Cluster 4 (6.3%) reflected a general pattern of multiple symptoms of high prevalence, with no single predominant symptom (9 of 14 symptoms with prevalence >75%).

Although studies by Coyne et al. [36] and Hall et al. [37] clustered women from the general population and used similar questionnaires, their resultant clusters differed substantially (see detailed comparison in Rosen et al. [76]). According to Coyne et al., four of five symptomatic clusters were defined by single predominant symptoms, while the study by Hall et al. defined all four symptomatic clusters by the combinations of symptoms; the latter result being similar to our findings. Both population studies (EPIC and BACH) and our treatment-seeking patient study (LURN) defined the cluster in which women experienced multiple LUTS symptoms at high severity level. This cluster (W5) contained 30.3% of our cohort, 5.5% of symptomatic women in EPIC, and 8.3% of symptomatic women in BACH, which is reasonable, given the fact that LURN recruited only treatment-seeking patients. This cluster was found to be higher in obesity indices, both in BACH and LURN studies.

Another study [38] on clustering women with LUTS was limited to data from bladder diary variables and BMI. That paper analyzed community dwelling (unlike our study, which focused on treatment-seeking patients), and continent and incontinent women from the EPI study [76], including those without LUTS. Clustering was based on six bladder diary variables: number of voids during daytime and night, most frequent voiding volume, 24-hour output, 24-hour beverage intake, and BMI. Three clusters identified in this study were described as: “Conventional” (n=233, with low 1320 mL daily intake, 6 voids, and low daily output of 1069 mL); “Benchmark” (n=96, with higher average daily intake of 2,445 mL, 8 voids, and average daily output of 1,907 mL); and “Superplus” (n=23, with extreme daily volumes of intake 3,774 mL and void 3,281 mL, and 12 voids). All clusters included continent and incontinent women, with a significantly higher percentage of incontinent (over 90%) in the "Superplus" cluster, characterized by excessive fluid intake. Authors of [38] clearly demonstrated the disadvantage of excessive fluid consumption. However, they were not using detailed LUTS information and were not aiming to identify subtypes of LUTS. On the contrary, our clusters defined by the multitude of LUTS variables were affected but not driven by bladder diary variables, i.e., only one of five bladder diary variables was used for clustering, and the average number of voids in 24 hours proved to be significantly different across the identified clusters. This comparison of our results with results of [38] emphasizes an obvious but often overlooked fact that clustering results are largely determined by the choice of variables used for clustering. If the number of variables is low, then it is important to be sure that none of the important variables is missed. In a sense, using a limited number of variables predefines the results of clustering by selecting only the variables presumed to be important. Using a comprehensive list of variables creates an opportunity for more objective, unbiased clustering, especially if the redundancy of variables is addressed by proper weighting, and the importance of variables is assessed through scaling by controls, as described in the Methods section.

### Details on the methodological information provided in the clustering papers [31-33]

Moore et al [31] report on clustering of 726 patients with asthma into 5 subtypes using hierarchical clustering algorithm on 34 core variables derived from initial 628 variables. "Half of the 34 variables that were included in the cluster analysis were numeric variables", for whom scaling information is not provided; while another half were composite variables derived by consolidating binary responses to multiple questions into "ranked severity scale". "All composite variables were assigned a range of 0 to 10 so that they were equally weighted in the analysis"; it is not clear however, how this corresponds to the range/scale of the 17 numerical variables. Criteria for choosing the number of clusters is not provided; however, evaluation of the distinctiveness of the identified clusters is performed by analysis of variance. Kruskal-Wallis and chi-square tests were used for parametric continuous, nonparametric continuous, and categorical variables, respectively. The important merit of this paper is that the investigators performed discriminant analysis and identified "11 most important variables that determine assignment to individual clusters, six are pulmonary function tests, two are related to age (age of onset and duration of asthma), two are composite variables that reflect medication use (corticosteroids, b-agonists), and one is gender."

Ahlqvist et al [32] reported on subtyping patients with diabetes using both hierarchical and k-means clustering on six variables: "glutamate decarboxylase antibodies (GADA), age at diagnosis, BMI, HbA1c, and homoeostatic model assessment 2 estimates". They reported that "Cluster analysis was done on values centered to a mean value of 0 and an SD of 1", meaning that they performed clustering on the z-scores (option discussed in our Methods section and Supplemental Material). "Presence or absence of GADA was included as a binary variable." Robustness of the clusters was ensured by "resampling of the data set 2000 times and computing the Jaccard similarities to the original clusters". Clustering was performed with k-means algorithm using Schwarz's Bayesian criterion to determine number of clusters k=4. "Only individuals negative for GADA were included because the k-means method does not accommodate binary variables", "all individuals who were GADA-positive were clustered together" into the fifth subtype. Note that, by doing this, authors avoided integrating categorical and continuous variables discussed in our Methods section; however, this approach likely would not work in case of multiple binary variables.

The important merit of the paper by Ahlqvist et al [32] is that they provide both detailed evaluation of the identified clusters and its validation in three independent cohorts. Box plots are provided comparing five continuous variables used for clustering across five clusters. Associations of clusters with variables not used for clustering, including known genetic risk factors, are provided. Authors acknowledge the importance and possibility "to refine the stratification further through inclusion of additional cluster variables, such as biomarkers, genotypes, or genetic risk scores."

Seymour et al [33] reported identification of four phenotypes of sepsis by consensus k-means clustering of 16,552 patients on 29 variables. These variables included "demographic variables (e.g., age, sex, Elixhauser comorbidities), vital signs (e.g., heart rate, respiratory rate, Glasgow Coma Scale score, systolic blood pressure, temperature, and oxygen saturation), markers of inflammation (e.g., white blood cell count, premature neutrophil count erythrocyte sedimentation rate, and C-reactive protein), markers of organ dysfunction or injury (e.g., alanine aminotransferase, aspartate aminotransferase, total bilirubin, blood urea nitrogen, creatinine, international normalized ratio, partial pressure of oxygen, platelets, and troponin), and serum levels of glucose, sodium, hemoglobin, chloride, bicarbonate, lactate, and albumin"[33]. For each variable, the most abnormal value recorded within the first 6 hours of hospital presentation was used. Variables were z-scored. To determine the optimal number of clusters, authors "evaluated a combination of phenotype size, clear separation of the consensus matrix heatmaps, characteristics of the consensus cumulative distribution function (CDF) plots, and adequate pairwise consensus values between cluster members (>0.8)". Supplemental Material of [33] provides a heat map of the consensus matrix demonstrating four distinct clusters, confirmed by analysis of the area under the CDF curve versus number of clusters, following the criterion originally proposed in [34]. The important merit of this paper is the detailed evaluation of the identified phenotypes including "ranked plots of variables by the mean standardized difference between the phenotype pairs". Authors also report the "proportion of patients with a probability of phenotype assignment on the margin, which was defined as between 45% and 55%", which is likely conveying information similar to our PCC.
